# Investigating causal relations between sleep duration and risks of adverse pregnancy and perinatal outcomes: Linear and nonlinear Mendelian randomization analyses in up to 356,069 European women

**DOI:** 10.1101/2022.01.19.21267648

**Authors:** Qian Yang, Maria C Magnus, Fanny Kilpi, Gillian Santorelli, Ana Gonçalves Soares, Jane West, Per Magnus, John Wright, Siri Eldevik Håberg, Eleanor Sanderson, Deborah A Lawlor, Kate Tilling, Maria Carolina Borges

**Affiliations:** MRC Integrative Epidemiology Unit at the University of Bristol, Bristol, UK; Population Health Sciences, Bristol Medical School, University of Bristol, UK; Centre for Fertility and Health, Norwegian Institute of Public Health, Oslo, Norway; Bradford Institute for Health Research, Bradford Teaching Hospitals NHS Foundation Trust, Bradford, UK; National Institute for Health Research Bristol Biomedical Centre, University Hospitals Bristol NHS Foundation Trust and University of Bristol, Bristol, UK

**Keywords:** sleep duration, Mendelian randomization, pregnancy, ALSPAC, MoBa

## Abstract

**Background:** Observational studies have reported maternal short/long sleep duration to be associated with adverse pregnancy and perinatal outcomes. However, it remains unclear whether there are nonlinear causal effects. Our aim was to use multivariable regression (MVreg) and Mendelian randomization (MR) to examine nonlinear effects of sleep duration on stillbirth, miscarriage, gestational diabetes, hypertensive disorders of pregnancy, perinatal depression, preterm birth, low/high offspring birthweight (LBW/HBW).

**Methods:** We used data from European women in UK Biobank (UKB, N=208,140), FinnGen (N=∼123,579), Avon Longitudinal Study of Parents and Children (N=6826), Born in Bradford (N=2940) and Norwegian Mother, Father and Child Cohort Study (MoBa, N=14,584). We used 78 previously identified genetic variants as instruments for sleep duration, and investigated its effectsusing two-sample MR and one-sample nonlinear MR (in UKB only). We compared MR findings with MVreg in MoBa (N=76,669), where maternal sleep duration was measured at 30 weeks.

**Results:** In UKB, MR provided evidence of nonlinear effects of sleep duration on stillbirth, perinatal depression and LBW, but not for other outcomes. Shorter and longer duration increased stillbirth and LBW; shorter duration increased perinatal depression. For example, there was a lower risk of LBW with increasing duration (odds ratio 0.79 per one-hour/day (95% confidence interval (CI): 0.68, 0.93) in the shortest duration group and a higher risk (odds ratio 1.38 (95% CI: 1.06, 1.81) in the longest duration group, suggesting shorter and longer duration increased the risk. These were supported by the lack of evidence of a linear effect of sleep duration with any outcome using two-sample MR. In MVreg, risks of all outcomes (except for stillbirth showing opposite directions) were higher in the women reporting <5- and ≥10-hours/day sleep compared with the reference category of 8-9-hours/day, despite some wide CIs including the null. Nonlinear models fitted the data better than linear models for most outcomes(likelihood ratio P-value=0.02 to 3.2×10^−52^), except for stillbirth and gestational diabetes.

**Conclusions:** Our results supported possible nonlinear sleep duration effects on perinatal depression and LBW. Statistical support for nonlinear models across outcomes suggests potential nonlinear effects on other outcomes that larger studies could detect.

## Introduction

Sleep occupies up to one third of the human life span. For adults, the minimum and maximum sleep durations are recommended as 7 and 9 hours/day (h/d), respectively [1]. Habitual sleep duration is regulated by genetic factors [2], and can be influenced by a person’s daily routine and lifestyle factors [1]. Pregnancy is associated with alterations in sleep duration: both total and nocturnal sleep duration tend to be longer around the end of the first trimester declining by the third trimester due to pregnancy-induced changes, e.g. uterine contractions, heartburn, orthopnoea, leg cramps, pelvic girdle pain and uncomfortable sleeping position [3-5].

A systematic review of observational studies published up to 15 January 2018, which explored associations of prenatal sleep duration with psychological outcomes, reported a linear association of longer sleep duration with a lower risk of perinatal depression [6]. Other systematic reviews indicate that both short (<6 or <7 h/d) and/or long (>9 h/d) duration are associated with higher risks of adverse perinatal events (see Table S1 in File S1). In particular, short sleep duration is associated with higher risks of gestational diabetes (GD) [7, 8], preeclampsia [8] and preterm birth (PTB) [8-10], while long sleep duration is associated with higher risks of stillbirth [8, 11] and GD [8, 12]. These observational studies may be vulnerable to residual confounding, with demonstrated between-study heterogeneity likely influenced by variation in confounder control [13]. Few studies have examined several outcomes together, which is important for trying to identify a range of duration that minimises any adverse outcomes. Studies to date have mostly examined binary variables of short and long sleep duration rather than trying to explore different patterns across sleep duration.

In the absence of large, well-conducted randomized controlled trials of interventions targeting on sleep duration during pregnancy [14], Mendelian randomization (MR) provides an alternative means of probing the effect of sleep duration on adverse pregnancy and perinatal outcomes. MR uses single nucleotide polymorphisms (SNPs) that are robustly associated with potential risk factors, e.g. sleep duration, as instrumental variables (IVs) to explore causal effects of these factors on outcomes [15, 16]. MR is less prone to confounding than observational studies, as SNPs being randomly allocated at meiosis cannot be influenced by the wide range of socio-demographic or behavioural factors conventionally confounding observational studies, nor can they be influenced by health status [15, 16]. Under key assumptions (discussed in **Methods**), MR can be used to estimate a causal effect from SNPs-risk factor and SNPs-outcome associations. SNPs robustly associated with self-report sleep duration have recently been identified in the most updated genome-wide association study (GWAS) using UK Biobank (UKB) [2]. These SNPs have been used as IVs in MR studies to investigate linear and nonlinear effects of sleep duration on cancer [17], cardiometabolic health [18, 19] and mental health [20, 21]. To the best of our knowledge, MR has not been used to explore effects of sleep duration on pregnancy and perinatal outcomes.

Our aims are to use MR to explore and compare linear and nonlinear effects of lifelong sleep duration on stillbirth, miscarriage, GD, hypertensive disorders of pregnancy (HDP), perinatal depression, PTB, low offspring birthweight (LBW), high offspring birthweight (HBW) and variation in BW in up to 356,069 women. We also compared MR findings with confounder-adjusted linear and nonlinear multivariable regression (MVreg) of maternal sleep duration reported during pregnancy with the same outcomes in 76,669 women.

## Methods

### Participants

This study was undertaken using data from the MR-PREG consortium, which aims to explore causes and consequences of different pregnancy and perinatal events [22]. As shown in Figure S1 (File S2), we include women of European descent from 1) UKB (208,140 women recruited at age 40-60 years between 2006-2010 and providing retrospective reports of pregnancy and perinatal outcomes), 2) FinnGen (the nation-wide network of Finnish biobank with up to 123,579 women recruited at 54 years (interquartile range=25) with outcomes obtained via health record linkage), and three birth cohorts (women recruited during pregnancy with most outcomes collected prospectively) – 3) Avon Longitudinal Study of Parents and Children (ALSPAC, 6826 women recruited between 1991-1992), 4) Born in Bradford (BiB, 2940 women recruited between 2007-2010), and 5) the Norwegian Mother, Father and Child Cohort Study (MoBa, 76,669 women in MVreg, of whom 14,585 women had genome wide data and were included in two-sample MR; recruited between 1999-2009). All studies had ethical approval from relevant national or local bodies and participants provided written informed consent. Details of their recruitment procedures, information on genetic data, and measurements of baseline characteristics are described in Text S1 (File S2).

### Self-report sleep duration

Information on sleep duration was obtained from UKB and MoBa. In UKB, sleep duration was measured via a self-administered question – “About how many hours sleep do you get in every 24 hours? (please include naps)” at the initial assessment centre, which recruited mostly non-pregnant participants. Women reported their sleep duration in integer values ranging from 1 to 23. Following a previous MR study [20], only women whose sleep duration ranged from 2 to 12 hours were included in our analyses (N=206,500 (99.2%)). The UKB data were used to identify genetic variants associated with sleep duration by the GWAS [2], and thus contributed to our two-sample (linear) and one-sample (nonlinear) MR.

In MoBa, sleep duration was assessed via a self-administered question – “How many hours a day do you usually sleep now when you are pregnant?” at 30 weeks of gestation. Women reported their sleep duration in five categories, which were “over 10 hours”, “8-9 hours”, “6-7 hours”, “4-5 hours” and “less than 4 hours”. The questionnaire did not specify a time period so it is unclear whether the women would have reported duration only for night sleep or across 24 hours (as in UKB). Due to small numbers, we combined the last two categories into “≤5 hours”. The MoBa sleep duration data were used to explore the relevance of genetic IVs in pregnancy, and linear/nonlinear associations in MVreg.

### Selection of genetic IVs for self-report sleep duration

Currently, nine GWAS of self-report sleep duration are available, with details in Table S2 (File S1) [23]. All GWAS combined women and men (mainly of White European descent) with no sex specific analyses; all large GWAS included UKB. The largest and most updated GWAS identified 78 SNPs genome-wide significantly (P-value<5×10^−8^) associated with sleep duration in its discovery cohort – UKB men and women (N=446,118), with 55 of them being directionally consistent in the replication cohort – the Cohorts for Heart and Aging Research in Genomic Epidemiology (CHARGE,N=47,180) [2]. Table S3 (File S1) lists the characteristics of the 78 SNPs, as well as summary data for their association with sleep duration in our UKB women. The 78 SNPs explained 0.69% of the variance in sleep duration in UKB [2]. We used the ‘clumping’ function from the TwoSample MR R package [24], to check that the 78 SNPs were independent (i.e. linkage equilibrium) based on a threshold of R^2^≤0.01 and all European samples from the 1000 genome project as the reference population. For two-sample MR, we used the 78 SNPs as IVs. For one-sample MR, we combined the 78 SNPs into an unweighted genetic risk score (GRS) [25], given weights could not be obtained from genetic association estimates generated in the GWAS discovery stage including UKB (due to sample overlap with our analyses sample) or replication stage using CHARGE (due to the trait increasing allele being inconsistent between discovery and replication stages for some SNPs).

The same GWAS has also reported results for short (defined as ≤6 h/d) and long (defined as ≥9 h/d) sleep duration, identifying 27 and 9 genome-wide significant SNPs, respectively [2]. We decided *a priori* not to use these in the one-sample MR in order to explore other possible nonlinear association or different thresholds of ‘healthy’ sleep duration to these and for different outcomes. There were further technical considerations that are described in more detail in Text S2 (File S2).

### Pregnancy and perinatal outcomes

We examined associations with nine adverse pregnancy and perinatal outcomes, including stillbirth, miscarriage, GD, HDP, perinatal depression (occurring during pregnancy or within a year after delivery), PTB (gestational age <37 completed weeks), LBW (BW <2500 grams), HBW (BW >4500 grams), and BW (grams) as a continuous outcome. Definitions of these outcomes in UKB, ALSPAC, BiB, MoBa and FinnGen, and harmonization of their definitions across cohorts are provided in Table S4 (File S1). If multiple pregnancies were enrolled in the birth cohorts, we randomly selected one pregnancy per woman. In FinnGen, it was only possible to include [miscarriage](https://risteys.finngen.fi/phenocode/O15_ABORT_SPONTAN) (N=9113 cases/89,340 controls), [GD](https://risteys.finngen.fi/phenocode/GEST_DIABETES) (N=5687 cases/117,892 controls), [HDP](https://risteys.finngen.fi/phenocode/O15_GESTAT_HYPERT) (N=4255 cases/114,735 controls) and [PTB](https://risteys.finngen.fi/phenocode/O15_PRETERM) (N=5480 cases/98,626 controls), and those outcomes were defined based on ICD codes [26].

We combined pre-eclampsia and gestational hypertension as HDP since we would not have sufficient statistical power to consider pre-eclampsia separately. In UKB gestational age was only available for a subset of women (N=7280) who were young enough to have had a child born during or after 1989, the earliest date for which linked hospital labour and perinatal data are available. As a result, numbers with data on PTB are smaller than for any other outcome, and we decided *a priori* to examine associations with LBW/HBW rather than small-/large-for-gestational age. In the three birth cohorts, stillbirth and miscarriage were retrospectively reported at the time of the index pregnancy when women were asked if they had ever experienced a (previous) stillbirth or miscarriage. In ALSPAC and MoBa, we could not include women having stillbirth or miscarriage in the index pregnancy in the MR, as they were not genotyped.

### Confounders in MoBa for MVreg

We considered maternal age at time of delivery, parity, education, smoking status in pregnancy, alcohol intake in pregnancy, body mass index before pregnancy and average household income as potential confounders based on their known or plausible effects on maternal sleep duration and on pregnancy and perinatal outcomes. Details of how these variables were measured are provided in Text S1 (File S2).

### Statistical analyses

#### One-sample MR exploring whether data support nonlinear over linear effects

One-sample MR requires individual level data for estimation, and thus we used data from UKB women (Figure 1). This involves generating subgroups of different sleep duration length within the study sample and undertaking (linear) MR within each of those subgroups and then comparing effects across subgroups. We could split the women into subgroups based on their reported sleep duration, but doing that could introduce a type of selection bias known as collider bias in the subsequent MR, because of the role of the genetic instruments on sleep duration [27]. To avoid that we generated ‘residual’ sleep duration by regressing self-reported sleep duration on the sleep duration GRS, adjusting for genetic array, women’s age and top 40 PCs (to adjust for residual population stratification [28]). Residual sleep duration was then calculated as each woman’s observed sleep duration minus the mean centred genetic contribution to sleep duration from the GRS) [27, 29]. Therefore, the residual measure has a mean of 7.18 with a range from 1.77 to 12.33 h/d (Figure S2 in File S2). We then stratified UKB women into three and five groups based on the residual duration (details shown in Table S5 of File S1). We present results for three and five groups, and compared effects of increasing sleep duration (measured in 1-hour units) from MR analyses across the groups. Three groups would enable comparing results between analyses with greater power and IV strength, while five groups provide finer gradation for more detailed exploration of nonlinearity.

**Figure 1.**
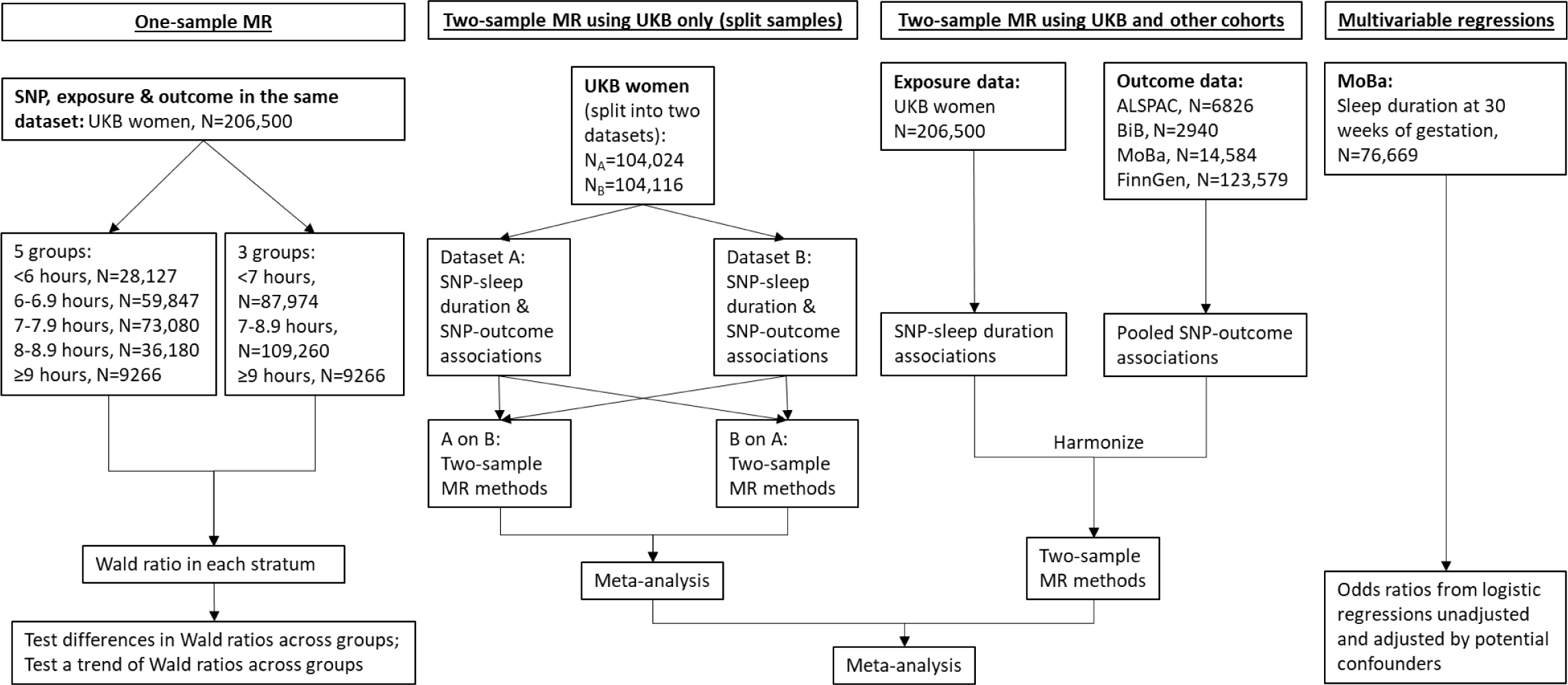
Summary of methods and data contributing to this study. In one-sample MR, the three and five groups of different duration lengths are based on thresholds from existing literature [1, 8]. We also split UKB women into thirds (N=68,833 or 68,834) as a sensitivity analysis to increase instrument strength and power in the longest duration group. Two-sample MR methods include: inverse variance weighted, MR-Egger, weighted median, and leave-one-out analysis. Multivariable regression analysis adjusted for maternal age at time of delivery, parity, education, smoking status in pregnancy, alcohol intake in pregnancy, body mass index before pregnancy and average household income. Abbreviations: ALSPAC, Avon Longitudinal Study of Parents and Children; BiB, Born in Bradford; MoBa, Norwegian Mother, Father and Child Cohort Study; MR, Mendelian randomization; SNP, single nucleotide polymorphism; UKB, UK Biobank.

Within each subgroup of residual sleep duration, we followed the approach used in a previous MR study [29] to calculate linear MR estimates for sleep duration on pregnancy and perinatal outcomes using the Wald ratio method [30]. Technical details of this method are described in Text 2 (File S2). Finally, we tested differences in MR estimates across groups using Cochran’s Q-statistic, with P-value <0.05 suggesting heterogeneity [27, 29]. We performed meta-regression of MR estimates against the mean of sleep duration in each group to test nonlinearity [27, 29]. A low P-value for the regression coefficient of sleep duration mean across groups provides evidence against the null hypothesis of a linear or null association, and we used the conventional P-value <0.05 as evidence to support nonlinearity. Similar non-null effects across the group (e.g. if there was evidence of a similar magnitude positive effect in all groups) would support a linear effect. Table 1 illustrates how the pattern of a nonlinear effect is identified by comparing the magnitudes and directions of linear associations in each group, and synthesizing results from one-sample and further two-sample MR.

**Table 1.**
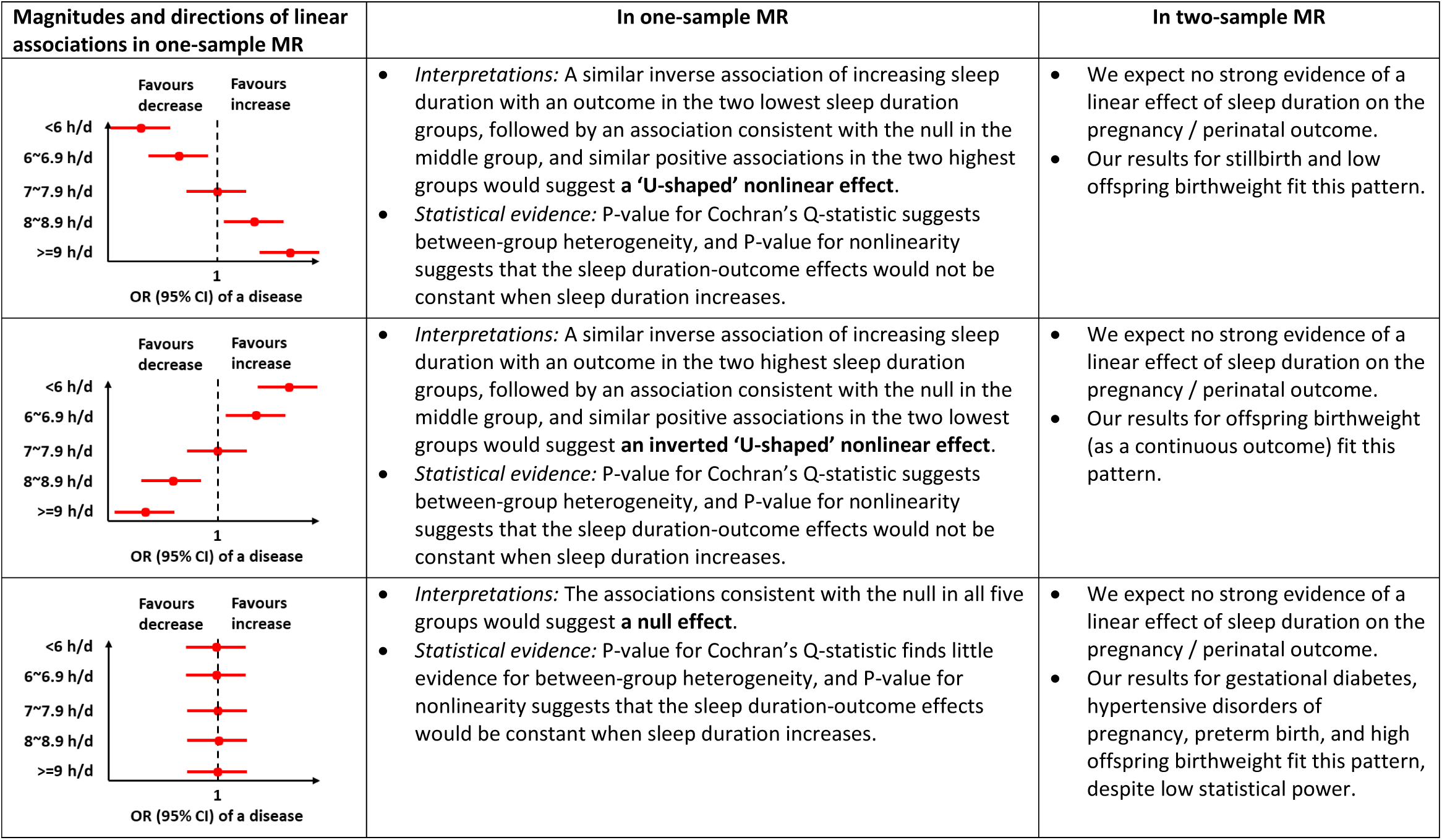
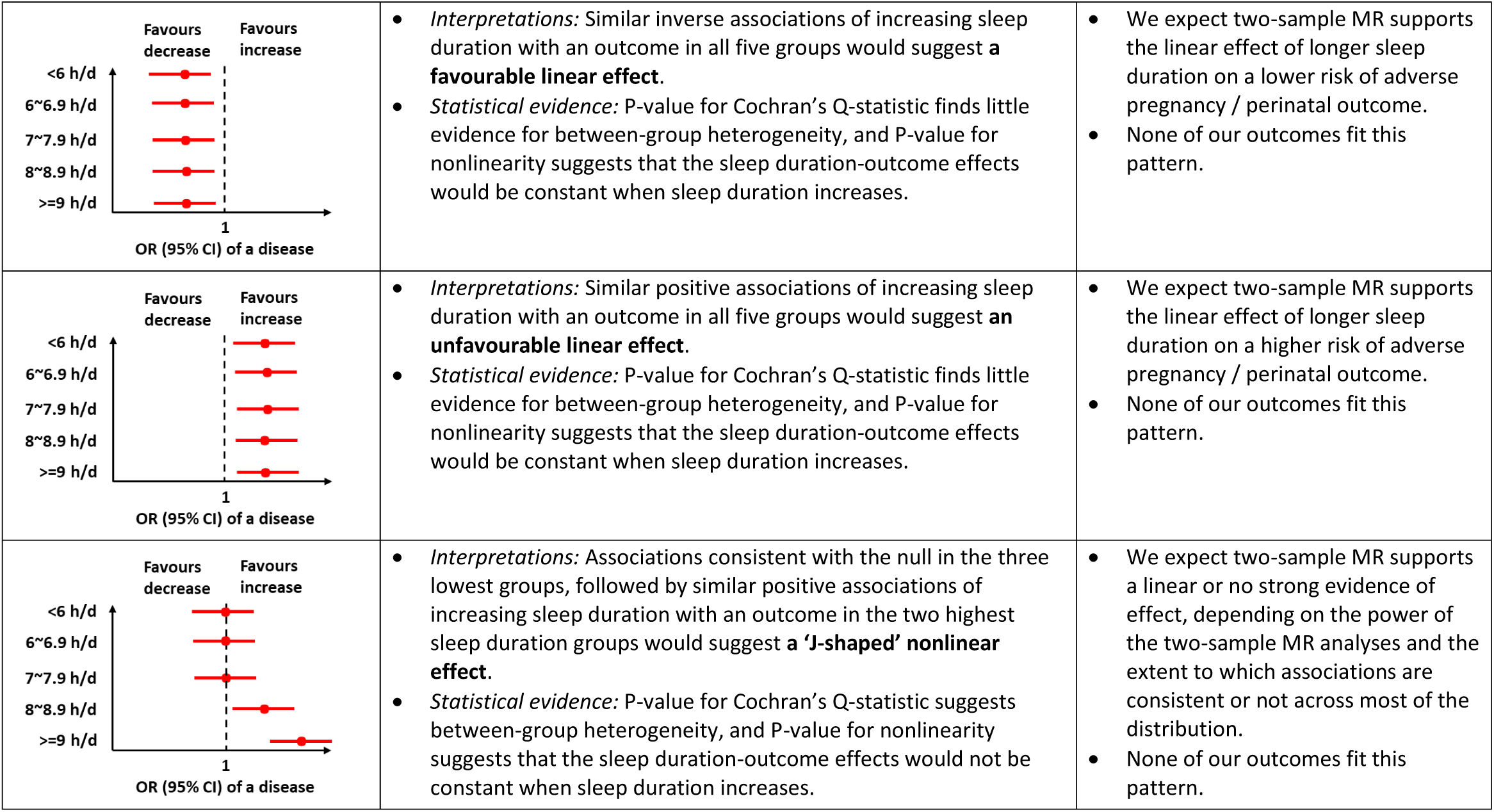
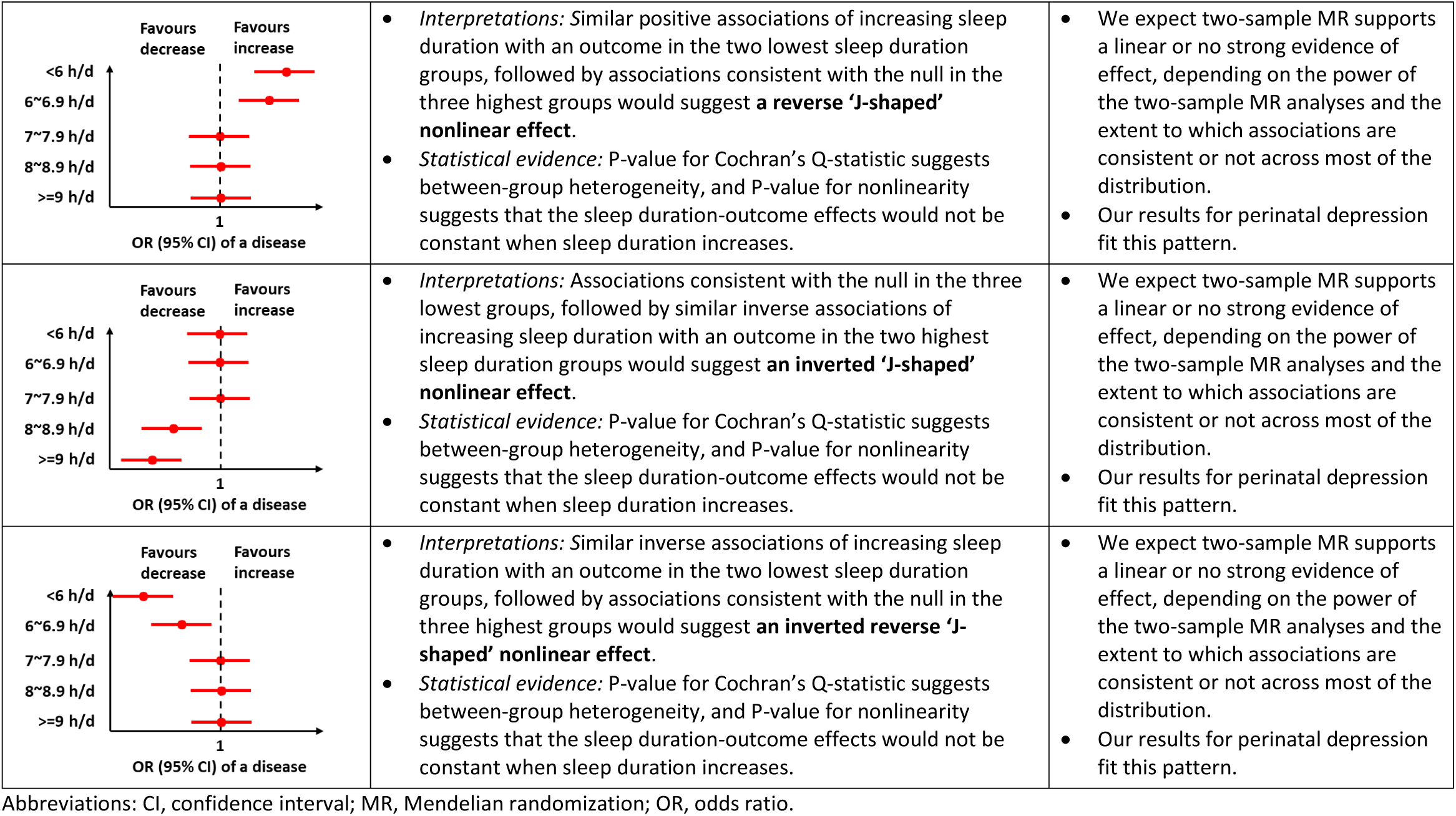
Possible patterns of effects identified by one-sample (using 5 groups as an illustration) and two-sample MR.

#### Two-sample MR exploring linear effects

We further undertook two-sample MR using both summary and individual participant data to explore potential linear effects (Figure 1). Details about obtaining SNP-sleep duration and SNP-outcome associations are described in Text S1 (File S2). In UKB women, we used individual participant data in a split cross-over samples design [20]. This involved randomly splitting the sample in half and generating summary data for SNP-sleep duration and SNP-outcome associations in both datasets and then conducting MR with SNP-sleep duration associations from dataset A and SNP-outcome associations from dataset B, and vice versa [20]. This was because the GWAS of sleep duration was conducted in UKB [2], and this split cross-over design enabled us to have the advantages (e.g. weak instrument bias towards the null and minimising over-prediction or winners curse) of MR using two independent samples [31, 32]. We then meta-analysed the MR estimates from the two together for each sleep duration-outcome pair using fixed-effects (with inverse variance weights). We also conducted two-sample MR using SNP-sleep duration associations from UKB women, and the pooled SNP-outcome associations combining ALSPAC, BiB, MoBa and FinnGen. For each outcome, we combined MR estimates from all five cohorts using fixed effects (with inverse variance weights). The degree of between-study heterogeneity was assessed using Cochran’s Q-statistic.

In the main analyses, we used the MR inverse variance weighted (IVW) method to explore the presence of linear effects of sleep duration on pregnancy and perinatal outcomes. IVW is a weighted regression of SNP-outcome associations on SNP-sleep duration associations with the intercept of the regression line forced through zero [33]. All two-sample MR analyses were conducted using “TwoSampleMR” package in R version 3.5.1 (R Foundation for Statistical Computing, Vienna, Austria).

#### Sensitivity and additional analyses

The strength of the IVs was evaluated by the F-statistic of IV-sleep duration associations [16]. We selected the 78 SNPs robustly related to sleep duration in general population rather than pregnant women [2]. Therefore, we used linear regression to test whether our IV was also related to sleep duration during pregnancy [15, 16]. The one-sample MR requires GRS-sleep duration association to be consistent across groups [27, 29]. We explored this by using Cochran’s Q-statistic and meta-regression of these associations against the mean of observed sleep duration in each stratum [27, 29].

Previous two-sample MR studies testing effects of sleep duration on different outcomes have used 78 genome-wide significant SNPs in the discovery sample, each having slightly different numbers after harmonization with outcome GWAS summary data [17-21]. Among the 78 GWAS significant SNPs, 55 SNPs gave estimates in the same direction in the replication cohort; 43 out of those 55 SNPs achieved genome-wide significance in the meta-analysis of the discovery and replication cohorts [2]. To maximise IV strength, we used the 78 GWAS significant discovery SNPs but repeated IVW analyses with the 55 SNPs and 43 SNPs to test whether our results would be robust to IV selection. To explore potential unbalanced horizontal pleiotropy, our sensitivity analyses for two-sample MR included (I) assessing between-SNP heterogeneity (which if present may be due to one or more SNPs having horizontal pleiotropic effects on the outcome) using Cochran’s Q-statistic and leave-one-out analysis [33], and (II) conducting weighted median [34] and MR-Egger [35], which are more likely to be more robust in the presence of horizontal pleiotropy [36]. Technical details of these MR methods were summarized in our previous study [37]. A consistent finding across multiple MR methods would strengthen causal inference. When using MR to assess maternal exposures in pregnancy on perinatal outcomes, results might be biased via a horizontal pleiotropic path from maternal genotype to the outcome via fetal genotype [38]. To explore this, we compared maternal SNP-outcome associations with versus without adjustments for fetal SNPs in the birth cohorts.

#### MVreg in MoBa exploring linear/nonlinear associations and whether data supported nonlinear over linear associations

We explored the observational association of maternal sleep duration at 30 weeks of gestation with each pregnancy and perinatal outcome using logistic regression (linear regression for BW). To explore a possible nonlinear association, we entered the categories as indicator variables and obtained estimates comparing each of ≤5 h/d, 6-7 h/d and ≥10 h/d to our chosen reference category of 8-9 h/d. We also explored possible linear associations by recoding ≤5, 6-7, 8-9 and ≥10 h/d categories using their mid-points (i.e. 3.5, 6.5, 8.5, 11 h/d, respectively), assuming MoBa had the same minimum and maximum sleep duration as UKB. Statistical evidence for a nonlinear association across categories was obtained from a likelihood ratio test comparing the two models above.

Following the previous MoBa study [39], women who had returned maternal questionnaires at both 15 and 30 weeks of gestation were eligible to our MVreg (N=76,669), and there were varying amounts of missing data for sleep duration, outcomes, and covariates. This was lowest for parity (0.2% missing), and highest for PTB (8.4% missing). Table S6 (File S1) provides full details of the proportion missing for each variable. Therefore, we undertook both complete records analyses and multiple imputation (MI). Complete records analyses only included women with sleep duration, an outcome, and six confounders (N=42,001 (for stillbirth) to 62,929 (for BW)), assuming that missingness is not associated with the outcome. MI was conducted on all 76,669 eligible women, and assumes data are missing at random (i.e. conditional on variables included in MI, the outcome would not differ between those with missing data and those without) [40, 41].

MI used chained equations [40], and was conducted for each outcome separately. As shown in Table S6 (File S1), each imputation model included one outcome, the exposure (sleep duration), the seven confounders (same as those used in complete records analyses), and three auxiliary variables (paternal education, paternal smoking status in pregnancy and maternal usage of other kinds of nicotine in pregnancy). These auxiliary variables were selected on the basis that they were likely to be important predictors of missing data, and of values for confounders that were missing, in particular maternal education and maternal smoking status in pregnancy. For each outcome, 100 imputed datasets were generated and results were pooled across these datasets using Robin’s Rules [42]. All MI analyses were conducted in Stata 16 (StataCorp LLC, College Station, Texas).

## Results

Table 2 summarizes the characteristics of included women for MR analyses, and the proportion of cases for pregnancy and perinatal disorders across the four cohorts, which differ substantially for some outcomes. Table S6 (File S1) summarizes the characteristics of MoBa women for MVreg analyses.

**Table 2.**
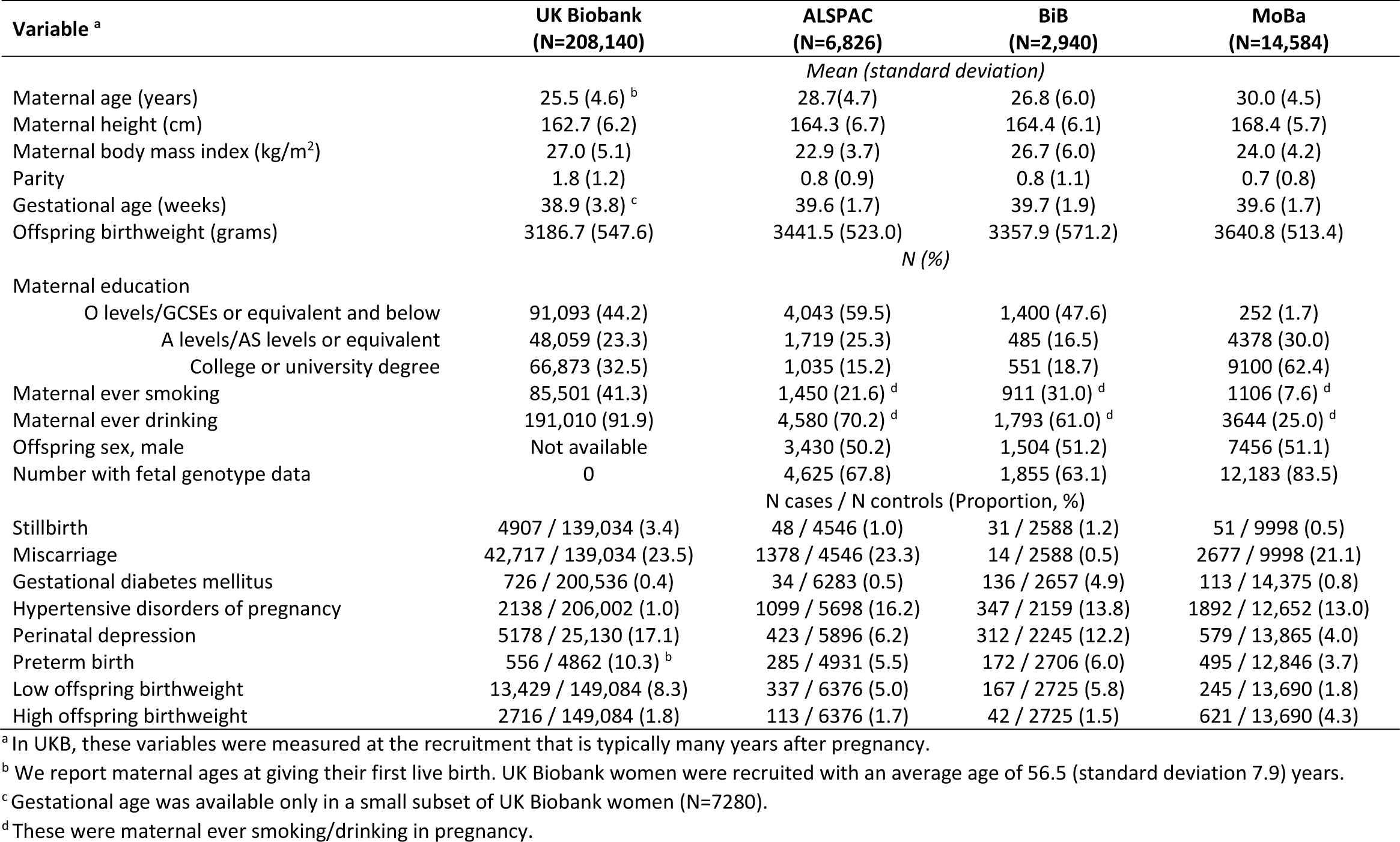
Characteristics of the women in UK Biobank, Avon Longitudinal Study of Parents and Children (ALSPAC), Born in Bradford, and Norwegian Mother and Child Cohort Study (MoBa) included in Mendelian randomization.

### One sample MR in UKB exploring whether data supported nonlinear over linear effects

The F-statistic for our GRS in all 206,500 UKB women was 1435, and within each group across all three ways in which they were stratified were between 64 and 759 (Table S5 in File S1). For each stratification approach, we also identified statistical evidence for between-group differences in associations of GRS with sleep duration. (Table S5 in File S1).

Figure 2 shows the MR linear effects within five sleep duration groups; the differences in directions and magnitudes of the results between groups can be used to describe the pattern of nonlinear effects of sleep duration. There was evidence of nonlinear effects of sleep duration on stillbirth, perinatal depression and LBW, with a pattern suggesting that both shorter and longer sleep duration increased risks of stillbirth and LBW, with shorter sleep duration increasing perinatal depression (Figure 2). The nonlinear effect with mean BW was broadly consistent with LBW as expected (Figure 2). For other outcomes there was no strong evidence of nonlinearity, though we acknowledge that several of the within-group results are imprecise with wide, and hence overlapping CIs. Patterns were generally similar for the analyses with three groups (Figure S3 in File S2).

**Figure 2.**
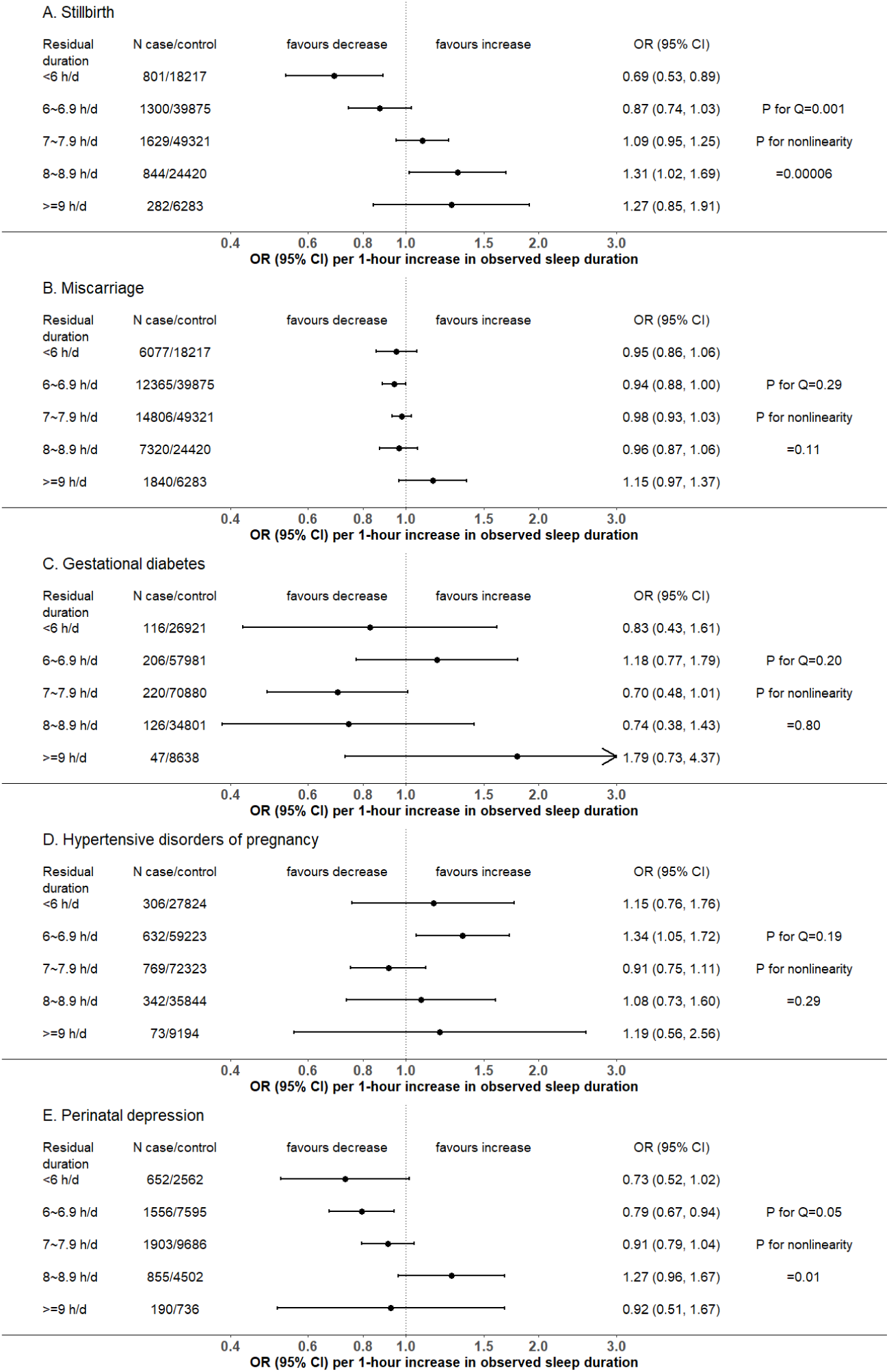
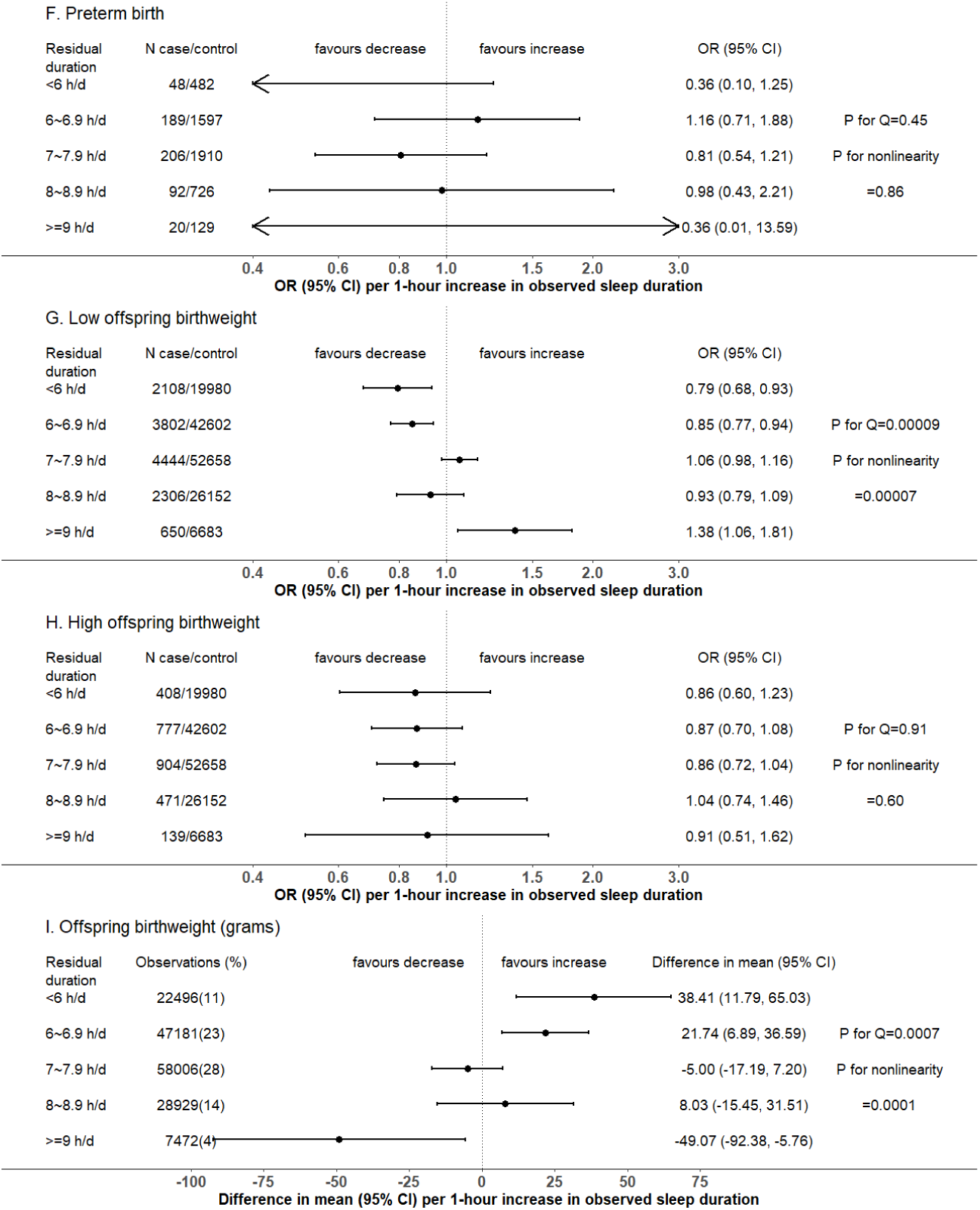
One-sample MR estimates of effects of sleep duration on pregnancy and perinatal outcomes in five groups (Total N=206,500) We present MR estimates of a linear effect of increasing duration on the outcome across the length of duration covered in each group. Further details about identifying the pattern of nonlinear effects are illustrated in Table 1. P-value for Cochran’s Q-statistic testing statistical evidence for between-group heterogeneity. P-value for nonlinearity testing statistical evidence whether the MR estimates are changed as the self-reported sleep duration mean increases. Abbreviation: CI, confidence interval; h/d, hours/day; MR, Mendelian randomization; OR, odds ratio.

### Two-sample MR exploring linear effects

In two-sample MR, the mean F-statistic for 78 sleep duration-IVs in 206,500 women was 18. In MoBa, one effect allele increase in the GRS for sleep duration was associated with 0.008 h/d longer sleep duration (95% CI: 0.003, 0.013, P-value =0.0009). Tables S3 & 7 (File S1) show summary results for SNP-sleep duration and SNP-outcome associations used in two-sample MR. After adjusting for fetal genotype (only possible in the birth cohorts), SNP-outcome associations with stillbirth, HDP, perinatal depression, PTB, and LBW were slightly attenuated; the associations with miscarriage, GD, HBW and BW moved slightly away from the null (Figure S4 in File S2).

The main IVW analyses combining UKB with other cohorts find no strong evidence to support a linear effect across the whole distribution of reported sleep duration of lifetime predisposition to longer average duration on the pregnancy and perinatal outcomes (Figure 3). However, 95% CIs were wide for most outcomes (Figure 3). Sensitivity analyses using weighted median and MR-Egger for these outcomes were largely directionally consistent (Figure 3). Between-SNP heterogeneity for MR analyses was observed for GD, HDP, perinatal depression, HBW and BW (Table S8 in File S1), but leave-one-out analyses suggested little evidence for a single SNP driving the MR IVW results (Figures S5-7 in File S2). The MR-Egger intercept P-value indicated unbalanced horizontal pleiotropy for stillbirth, but not for other outcomes (Table S8 in File S1). Across all outcomes there was little evidence of between-study heterogeneity (all Cochran’s P-values ≥0.1, Figure 3). IVW analyses using 55 SNPs and 43 SNPs (i.e. those with some evidence of replication) were broadly consistent with those from our main analyses with 78 SNPs (Figure S8 in File S2).

**Figure 3.**
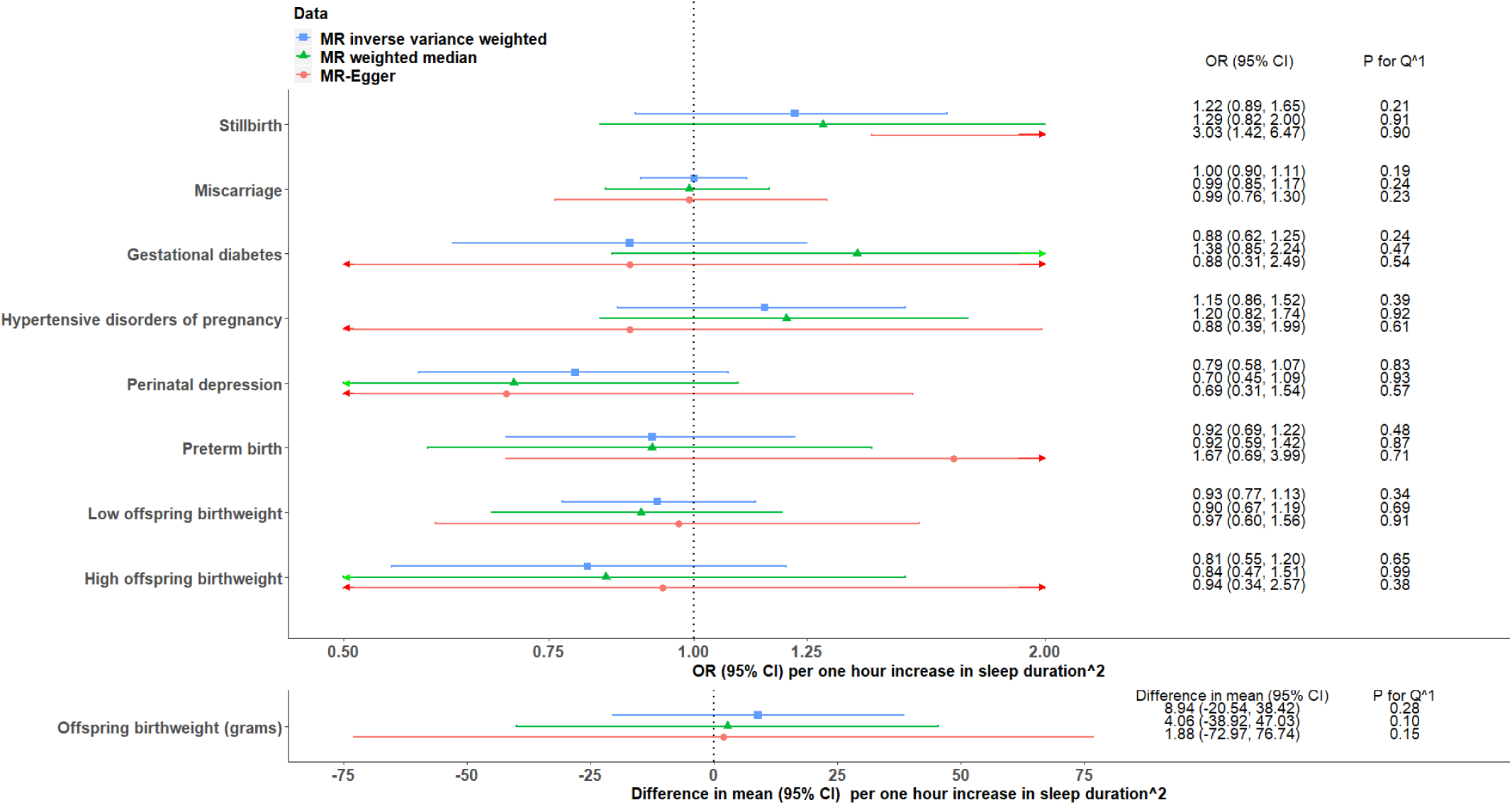
Two-sample Mendelian randomization (MR) estimates for linear effects of sleep duration on pregnancy and perinatal outcomes, meta-analysing UK Biobank, birth cohorts and FinnGen. ^1^P-value for Cochran’s Q-statistic testing statistical evidence for between-cohort heterogeneity. ^2^Results are ORs for binary outcomes (mean difference in birthweight) per lifetime genetic tendency to 1 hour longer during the 24-hour duration. Abbreviations: CI, confidence interval; OR, odds ratio.

### MVreg in MoBa exploring and comparing linear and nonlinear associations

Nonlinear models fitted data better than linear models across most outcomes (likelihood ratio P-values comparing the linear versus nonlinear models = 0.02 to 3.2×10^−52^). The exceptions, based on the conventional threshold of <0.05, were stillbirth (0.13) and GD (0.06). Odds of miscarriage, GD, HDP, perinatal depression, PTB, LBW and HBW were higher in women reporting ≤5 and ≥10 hours sleep compared with the reference category of 8-9 hours, despite some wide CIs including the null (Figure 4). Among them, perinatal depression showed considerably stronger magnitudes of associations than other outcomes, and evidence of increased odds in women reporting 6-7 hours (Figure 4). Odds of stillbirth and differences in mean BW were lower for those reporting ≤5, 6-7 and ≥10 hours compared to 8-9, but with very wide CIs including the null for stillbirth (Figure 4). Broadly consistent results were observed for LBW and HBW when all eligible participants were included in MVreg (Figure S9 in File S2), and for complete records analyses (Table S9 in File S1).

**Figure 4.**
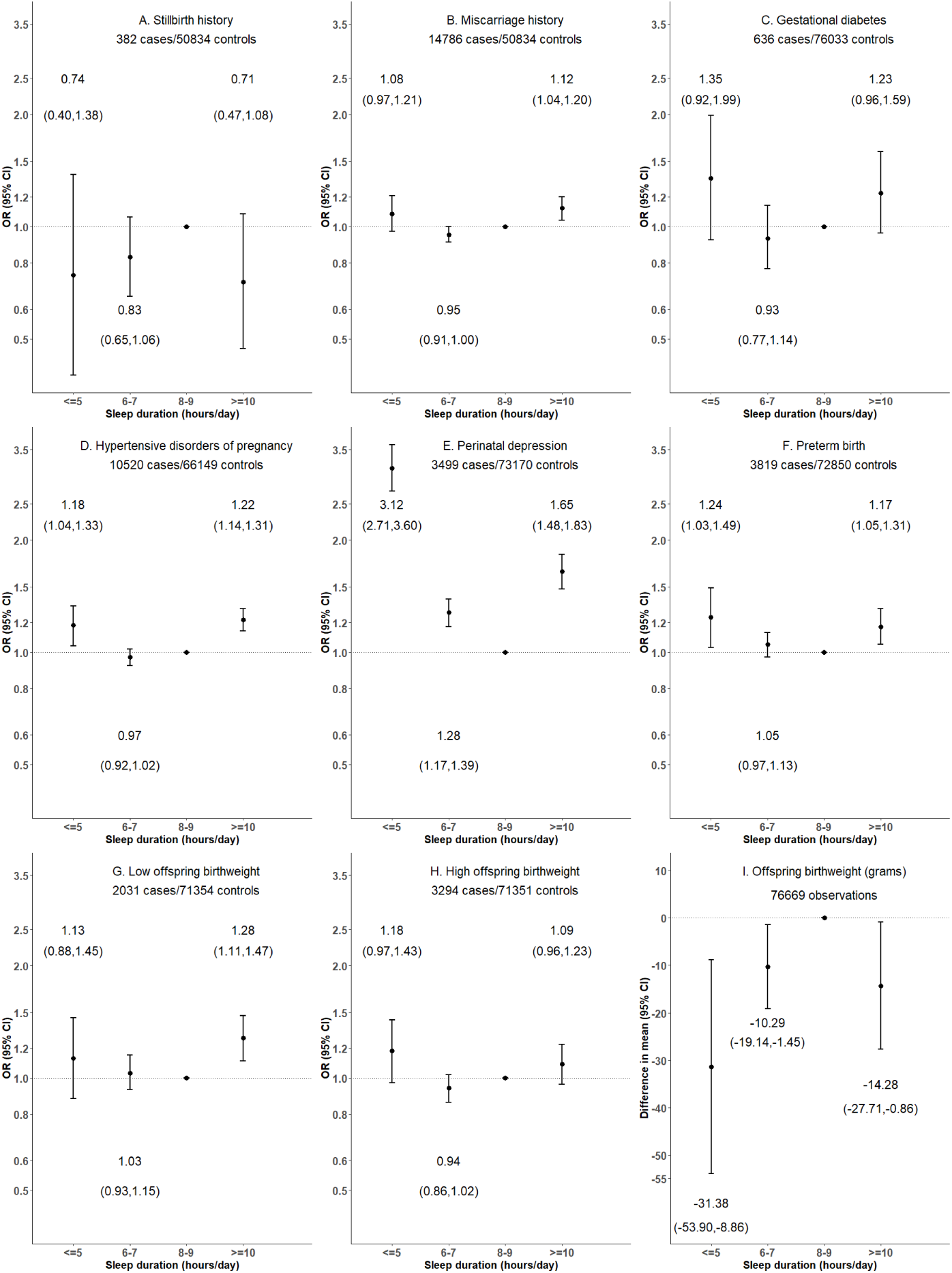
Multivariable regression associations of self-reported sleep duration categories with pregnancy and perinatal outcomes in Norwegian Mother, Father and Child Cohort Study (N=76,669) Estimates are from multiple imputation, which are also listed in Table S9 (File S1). Abbreviations: CI, confidence interval; OR, odds ratio.

## Discussion

To our knowledge, this is the first MR study exploring the effect of maternal sleep duration on adverse pregnancy and perinatal outcomes. We interpreted the MR results as reflecting a lifetime tendency to differences in mean 24-hour sleep duration on the basis that SNPs are determined at conception [43]. We interpreted MVreg results as reflecting associations of sleep duration in pregnancy, though this exposure is likely to correlate strongly with sleep duration before and after pregnancy. The association between our GRS and sleep duration at 30 weeks of gestation in MoBa indicated that genetic susceptibility to lifetime longer sleep duration is correlated with sleep duration during pregnancy. Overall, our MR analyses suggested that shorter and longer sleep duration potentially increase the risks of stillbirth and LBW, and, consistent with the latter decrease mean BW. They also suggest that shorter sleep duration increases the risk of perinatal depression. MVreg suggested an increased risk of perinatal depression and lower mean BW (with both short and long duration), and an increased risk of LBW (only for long duration), but little evidence for nonlinear associations for stillbirth. Notably there was statistical evidence that nonlinear models fitted the MVreg data better than linear models for most outcomes, highlighting the importance of further investigation of this in larger samples.

Previous observational studies have tended to assume nonlinearity and explored associations of short and long sleep duration with most pregnancy and perinatal outcomes, reporting unfavourable associations of both short and long duration with GD, short duration with pre-eclampsia and PTB, and long duration with stillbirth [7-12]. With the exception of stillbirth, our MVreg analyses in one of the largest studies to date, were broadly consistent with these findings. In MVreg, we found evidence of increased risks of perinatal depression with short and long duration, whereas a previous systematic review reported an association with shorter sleep duration only, which is consistent with our one-sample MR analyses [6]. MR analyses also supported increased risks of stillbirth and LBW with shorter and longer duration. MR evidence for other outcomes was too imprecise to make reliable conclusions. Furthermore, differences between what have been estimated (linear effects within groups of residual duration in one-sample MR versus odds of outcomes comparing for self-reported duration categories to a reference category in MVreg) make direct comparisons between our MR results versus our, and other published, MVreg associations difficult. The broad consistencies of nonlinear associations of sleep duration with a range of pregnancy and perinatal outcomes between our MVreg and existing literature highlight the need for further MR analyses of potential nonlinear effects in much larger sample sizes.

### Strengths and limitations

The main strengths of our study include that 1) as far as we are aware it is the first time that MR has been used to explore potential linear and nonlinear effects of sleep duration on pregnancy and perinatal outcomes; 2) we triangulated our MR findings with our MVreg results of sleep duration in pregnancy, adjusting for key confounders defined *a prior* [44]; 3) we explored a range of pregnancy and perinatal outcomes. To our knowledge, this is the first study to explore associations with miscarriage, and the largest observational study to explore associations of maternal sleep duration in pregnancy with all other outcomes.

Our MR analyses may be vulnerable to weak instrument bias, especially within some sleep duration groups in one-sample MR. Therefore, we used GRS to minimise the contribution of weak IVs in one-sample MR (with the lowest F-statistic=64), but our two-sample MR results may be biased towards the null [31]. Our MR analyses may be biased by horizontal pleiotropy, particularly because IVs for sleep duration have also been strongly linked to other sleep traits [2], as well as lifestyle factors such as obesity and alcohol consumption [45]. We explored the potential presence of bias by horizontal pleiotropy with sensitivity analyses for two-sample MR, which are more robust to such bias than IVW [36], but one-sample MR estimates in each stratum could still be prone to horizontal pleiotropy. We relied on three birth cohorts to further validate that fetal genotype could not be a substantial source of unbalanced horizontal pleiotropy for our outcomes. The MR monotonicity assumption requires that the proposed genetic IVs for longer sleep duration cannot increase sleep duration in some women while decreasing it in others – i.e. women are ‘compliers’ [46]. In our one-sample MR, constant GRS-duration effects across all groups is a stronger version of the monotonicity assumption [27]. As we found slightly different GRS-duration associations across groups, our one-sample MR results should be interpreted cautiously as tests for causal directions rather than as precise estimations of causal effects [46].

Both our MR and MVreg estimates could be vulnerable to selection bias as discussed in detail in other papers [47-49]. UKB participants are better educated and healthier than the general UK adult population [50], and perinatal depression and PTB may not be missing at random [51, 52]. Furthermore, by definition our MR and observational analyses only includes women who have ever experienced at least one pregnancy (i.e. fertility). Such selection could result in index event bias [53, 54], as a recent systematic review demonstrated weak evidence for a link between short sleep duration and infertility [55], and the lead SNP in *FADS1* gene for number of children was also associated with sleep duration in UKB [56]. Further studies are warranted to explore whether index event bias due to conditioning on pregnancy status would have a meaningful impact on MR estimates.

Sleep duration was measured via one self-administrated question in UKB and MoBa. Its measurement error in UKB may not bias our MR estimates, as sleep duration is a continuous variable in 1-hour units [57]. However, non-differential misclassification of sleep duration categories in MoBa would be expected to bias our MVreg towards the null [58]. Further studies using large, actigraphy-based sleep duration data would be required to explore optimal cut-off points for our suggestive nonlinear associations [27]. Moreover, it was ambiguous whether the variable in MoBa represented nocturnal sleep duration or included daytime naps. For our outcomes, there may be misclassification because of the absence of universal testing, assessment via self-report questionnaires, and differences between studies in definitions. Our MVreg analyses for pregnancy outcomes (including stillbirth, miscarriage, GD, HDP and perinatal depression) in MoBa could be vulnerable to reverse causality, as they were defined based on some information earlier than 30 weeks of the index pregnancy. Furthermore, our MVreg analyses in MoBa could be biased by residual and unmeasured confounding.

To conclude, our study raised the possibility of nonlinear effects of sleep duration on perinatal depression, LBW and BW, which might have public health implications for suggesting an optimal range of sleep duration for pregnancy health. The statistical support for nonlinear (compared to linear) models across outcomes suggested potential nonlinear effects on other outcomes that we did not have sufficient power to assess. We did not find conclusive evidence supporting linear effects of sleep duration on pregnancy and perinatal outcomes. However, even with large sample sizes some MR estimates are imprecise, and their point estimates suggested a potentially important clinical effect (e.g. 21% decreased odds of perinatal depression per 1-h/d in two-sample MR). We acknowledged the need for further MR studies based on larger number of cases for pregnancy and perinatal outcomes, and studies in women from non-White European ethnic background.

## Data Availability

We used both individual participant cohort data and publicly available summary statistics. We present summary statistics that we generated from those individual participant cohort data in Tables S3 & S7 in File S1. Full information on how to access UKB data can be found at its website (https://www.ukbiobank.ac.uk/researchers/). All ALSPAC data are available to scientists on request to the ALSPAC Executive via this website (http://www.bristol.ac.uk/alspac/researchers/), which also provides full details and distributions of the ALSPAC study variables. Similarly, data from BiB are available on request to the BiB Executive (https://borninbradford.nhs.uk/research/how-to-access-data/). Data from MoBa are available from the Norwegian Institute of Public Health after application to the MoBa Scientific Management Group (see its website https://www.fhi.no/en/op/data-access-from-health-registries-health-studies-and-biobanks/data-access/applying-for-access-to-data/ for details). Summary statistics from FinnGen are publicly available on its website (https://finngen.gitbook.io/documentation/data-download).

## Acknowledgments

This research has been conducted using the UKB Resources under application number 23938. The authors would like to thank the participants and researchers from UKB who contributed or collected data. We are extremely grateful to all the families who took part in this study, the midwives for their help in recruiting them, and the whole ALSPAC team, which includes interviewers, computer and laboratory technicians, clerical workers, research scientists, volunteers, managers, receptionists and nurses. BiB is only possible because of the enthusiasm and commitment of the Children and Parents in BiB. We are grateful to all the participants, teachers, school staff, health professionals and researchers who have made BiB happen. This research has been conducted using MoBa data using application number 2552. MoBa is supported by the Norwegian Ministry of Health and Care services and the Ministry of Education and Research. We are grateful to all the participating families in Norway who take part in this on-going cohort study. We thank the Norwegian Institute of Public Health (NIPH) for generating high-quality genomic data. This research is part of the HARVEST collaboration, supported by the Research Council of Norway (#229624). We also thank the NORMENT Centre for providing genotype data, funded by the Research Council of Norway (#223273), South East Norway Health Authority and KG Jebsen Stiftelsen. We further thank the Center for Diabetes Research, the University of Bergen for providing genotype data and performing quality control and imputation of the data funded by the ERC AdG project SELECTionPREDISPOSED, Stiftelsen Kristian Gerhard Jebsen, Trond Mohn Foundation, the Research Council of Norway, the Novo Nordisk Foundation, the University of Bergen, and the Western Norway health Authorities (Helse Vest). The authors thank FinnGen investigators for sharing their summary-level data.

## References

1. Hirshkowitz M, Whiton K, Albert SM, et al. National Sleep Foundation’s updated sleep duration recommendations: final report. Sleep Health. 2015; 1(4):233–43.

2. Dashti HS, Jones SE, Wood AR, et al. Genome-wide association study identifies genetic loci for self-reported habitual sleep duration supported by accelerometer-derived estimates. Nat Commun. 2019; 10(1):1100.

3. Pengo MF, Won CH, Bourjeily G. Sleep in Women Across the Life Span. Chest. 2018; 154(1):196–206.

4. Lamberg L. Sleeping poorly while pregnant may not be “normal”. JAMA. 2006; 295(12):1357–61.

5. Mindell JA, Cook RA, Nikolovski J. Sleep patterns and sleep disturbances across pregnancy. Sleep Med. 2015; 16(4):483–8.

6. Pauley AM, Moore GA, Mama SK, et al. Associations between prenatal sleep and psychological health: a systematic review. J Clin Sleep Med. 2020; 16(4):619–30.

7. Yang Z, Zhu Z, Wang C, et al. Association between adverse perinatal outcomes and sleep disturbances during pregnancy: a systematic review and meta-analysis. J Matern Fetal Neonatal Med. 2020:1–9.

8. Lu Q, Zhang X, Wang Y, et al. Sleep disturbances during pregnancy and adverse maternal and fetal outcomes: a systematic review and meta-analysis. Sleep Med Rev. 2020; 58:101436.

9. Warland J, Dorrian J, Morrison JL, et al. Maternal sleep during pregnancy and poor fetal outcomes: A scoping review of the literature with meta-analysis. Sleep Med Rev. 2018; 41:197–219.

10. Wang L, Jin F. Association between maternal sleep duration and quality, and the risk of preterm birth: a systematic review and meta-analysis of observational studies. BMC Pregnancy Childbirth. 2020; 20(1):125.

11. Cronin RS, Wilson J, Gordon A, et al. Associations between symptoms of sleep-disordered breathing and maternal sleep patterns with late stillbirth: Findings from an individual participant data meta-analysis. PLoS One. 2020; 15(3):e0230861.

12. Zhang X, Zhang R, Cheng L, et al. The effect of sleep impairment on gestational diabetes mellitus: a systematic review and meta-analysis of cohort studies. Sleep Med. 2020; 74:267–77.

13. Metelli S, Chaimani A. Challenges in meta-analyses with observational studies. Evid Based Ment Health. 2020; 23(2):83–7.

14. Bacaro V, Benz F, Pappaccogli A, et al. Interventions for sleep problems during pregnancy: a systematic review. Sleep Med Rev. 2019; 50:101234.

15. Davey Smith G, Ebrahim S. ‘Mendelian randomization’: can genetic epidemiology contribute to understanding environmental determinants of disease? Int J Epidemiol. 2003; 32(1):1–22.

16. Lawlor DA, Harbord RM, Sterne JA, et al. Mendelian randomization: using genes as instruments for making causal inferences in epidemiology. Stat Med. 2008; 27(8):1133–63.

17. Richmond RC, Anderson EL, Dashti HS, et al. Investigating causal relations between sleep traits and risk of breast cancer in women: mendelian randomisation study. BMJ. 2019; 365:l2327.

18. Daghlas I, Dashti HS, Lane J, et al. Sleep Duration and Myocardial Infarction. J Am Coll Cardiol. 2019; 74(10):1304–14.

19. Wang J, Kwok MK, Au Yeung SL, et al. Sleep duration and risk of diabetes: Observational and Mendelian randomization studies. Prev Med. 2019; 119:24–30.

20. Henry A, Katsoulis M, Masi S, et al. The relationship between sleep duration, cognition and dementia: a Mendelian randomization study. Int J Epidemiol. 2019; 48(3):849–60.

21. Anderson EL, Richmond RC, Jones SE, et al. Is disrupted sleep a risk factor for Alzheimer’s disease? Evidence from a two-sample Mendelian randomization analysis. Int J Epidemiol. 2020; 50(3):817–28.

22. Brand JS, Gaillard R, West J, et al. Associations of maternal quitting, reducing, and continuing smoking during pregnancy with longitudinal fetal growth: Findings from Mendelian randomization and parental negative control studies. PLoS Med. 2019; 16(11):e1002972.

23. Garfield V. Sleep duration: A review of genome-wide association studies (GWAS) in adults from 2007 to 2020. Sleep Med Rev. 2020; 56:101413.

24. Hemani G, Zheng J, Elsworth B, et al. The MR-Base platform supports systematic causal inference across the human phenome. Elife. 2018; 7.

25. Hartwig FP, Davies NM. Why internal weights should be avoided (not only) in MR-Egger regression. Int J Epidemiol. 2016; 45(5):1676–8.

26. FinnGen. FinnGen Documentation of R5 release. 2021. Available from: https://finngen.gitbook.io/documentation/.

27. Staley JR, Burgess S. Semiparametric methods for estimation of a nonlinear exposure-outcome relationship using instrumental variables with application to Mendelian randomization. Genet Epidemiol. 2017; 41(4):341–52.

28. Lawson DJ, Davies NM, Haworth S, et al. Is population structure in the genetic biobank era irrelevant, a challenge, or an opportunity. Hum Genet. 2020; 139:23–41.

29. Sun YQ, Burgess S, Staley JR, et al. Body mass index and all cause mortality in HUNT and UK Biobank studies: linear and non-linear mendelian randomisation analyses. BMJ. 2019; 364:l1042.

30. Palmer TM, Sterne JA, Harbord RM, et al. Instrumental variable estimation of causal risk ratios and causal odds ratios in Mendelian randomization analyses. Am J Epidemiol. 2011; 173(12):1392–403.

31. Lawlor DA. Commentary: Two-sample Mendelian randomization: opportunities and challenges. Int J Epidemiol. 2016; 45(3):908–15.

32. Burgess S, Davies NM, Thompson SG. Bias due to participant overlap in two-sample Mendelian randomization. Genet Epidemiol. 2016; 40(7):597–608.

33. Burgess S, Dudbridge F, Thompson SG. Combining information on multiple instrumental variables in Mendelian randomization: comparison of allele score and summarized data methods. Stat Med. 2016; 35(11):1880–906.

34. Bowden J, Davey Smith G, Haycock PC, et al. Consistent Estimation in Mendelian Randomization with Some Invalid Instruments Using a Weighted Median Estimator. Genet Epidemiol. 2016; 40(4):304–14.

35. Bowden J, Davey Smith G, Burgess S. Mendelian randomization with invalid instruments: effect estimation and bias detection through Egger regression. Int J Epidemiol. 2015; 44(2):512–25.

36. Hemani G, Bowden J, Davey Smith G. Evaluating the potential role of pleiotropy in Mendelian randomization studies. Hum Mol Genet. 2018; 27(R2):R195–r208.

37. Yang Q, Sanderson E, Tilling K, et al. Exploring and mitigating potential bias when genetic instrumental variables are associated with multiple non-exposure traits in Mendelian randomization. medRxiv. 2019.

38. Lawlor D, Richmond R, Warrington N, et al. Using Mendelian randomization to determine causal effects of maternal pregnancy (intrauterine) exposures on offspring outcomes: Sources of bias and methods for assessing them. Wellcome Open Res. 2017; 2:11.

39. Lupattelli A, Wood ME, Nordeng H. Analyzing Missing Data in Perinatal Pharmacoepidemiology Research: Methodological Considerations to Limit the Risk of Bias. Clin Ther. 2019; 41(12):2477–87.

40. Lee KJ, Tilling KM, Cornish RP, et al. Framework for the treatment and reporting of missing data in observational studies: The Treatment And Reporting of Missing data in Observational Studies framework. J Clin Epidemiol. 2021; 134:79–88.

41. Hughes RA, Heron J, Sterne JAC, et al. Accounting for missing data in statistical analyses: multiple imputation is not always the answer. Int J Epidemiol. 2019; 48(4):1294–304.

42. Rubin DB. Multiple Imputation for Nonresponse in Surveys. New York: John Wiley and Sons; 2004.

43. Burgess S, Davey Smith G, Davies NM, et al. Guidelines for performing Mendelian randomization investigations. Wellcome Open Res. 2019; 4:186.

44. Lawlor DA, Tilling K, Davey Smith G. Triangulation in aetiological epidemiology. Int J Epidemiol. 2016; 45(6):1866–86.

45. Kamat MA, Blackshaw JA, Young R, et al. PhenoScanner V2: an expanded tool for searching human genotype-phenotype associations. Bioinformatics. 2019; 35(22):4851–3.

46. Labrecque J, Swanson SA. Understanding the Assumptions Underlying Instrumental Variable Analyses: a Brief Review of Falsification Strategies and Related Tools. Curr Epidemiol Rep. 2018; 5(3):214–20.

47. Hughes RA, Davies NM, Davey Smith G, et al. Selection Bias When Estimating Average Treatment Effects Using One-sample Instrumental Variable Analysis. Epidemiology. 2019; 30(3):350–7.

48. Munafò MR, Tilling K, Taylor AE, et al. Collider scope: when selection bias can substantially influence observed associations. Int J Epidemiol. 2018; 47(1):226–35.

49. Paternoster L, Tilling K, Davey Smith G. Genetic epidemiology and Mendelian randomization for informing disease therapeutics: Conceptual and methodological challenges. PLoS Genet. 2017; 13(10):e1006944.

50. Fry A, Littlejohns TJ, Sudlow C, et al. Comparison of Sociodemographic and Health-Related Characteristics of UK Biobank Participants With Those of the General Population. Am J Epidemiol. 2017; 186(9):1026–34.

51. Adams MJ, Hill WD, Howard DM, et al. Factors associated with sharing e-mail information and mental health survey participation in large population cohorts. Int J Epidemiol. 2020; 49(2):410–21.

52. Davis KAS, Coleman JRI, Adams M, et al. Mental health in UK Biobank - development, implementation and results from an online questionnaire completed by 157 366 participants: a reanalysis. BJPsych Open. 2020; 6(2):e18.

53. Mahmoud O, Dudbridge F, Davey Smith G, et al. Slope-Hunter: A robust method for index-event bias correction in genome-wide association studies of subsequent traits. bioRxiv. 2020.

54. Diemer EW, Labrecque JA, Neumann A, et al. Mendelian randomisation approaches to the study of prenatal exposures: A systematic review. Paediatr Perinat Epidemiol. 2020; 35(1):130–42.

55. Caetano G, Bozinovic I, Dupont C, et al. Impact of sleep on female and male reproductive functions: a systematic review. Fertil Steril. 2020; 115(3):715–31.

56. Mathieson I, Day FR, Barban N, et al. Genome-wide analysis identifies genetic effects on reproductive success and ongoing natural selection at the FADS locus. bioRxiv. 2020.

57. Pierce BL, VanderWeele TJ. The effect of non-differential measurement error on bias, precision and power in Mendelian randomization studies. Int J Epidemiol. 2012; 41(5):1383–93.

58. Hutcheon JA, Chiolero A, Hanley JA. Random measurement error and regression dilution bias. BMJ. 2010; 340:c2289.

