## Supplementary material for "Investigating causal relations between sleep duration and risks of adverse pregnancy and perinatal outcomes: Linear and nonlinear Mendelian randomization analyses in up to 356,069 European women": File S2

**Text S1. Descriptions of each cohort**

***UK Biobank (UKB)***

All people in the UK National Health Service registry aged between 40-69 years and living within a 25 mile radius from one of 22 study centres were invited to participate between 2006-2010 [1]. In total 503,325 adults (5.5% of the ~9.2 million invited) were recruited into UKB [1]. Participants who had a valid email address (N=339,229) were invited to fill in a detailed online questionnaire assessing their mental health after January 2015, and 158,835 participants fully completed it by October 2017 [2]. Ethical approval for UKB was obtained from the North West Multi-centre Research Ethics Committee, and our study was performed under UKB application number 23938.

Genotyping, pre-imputation quality control, and imputation procedures were described in detail elsewhere [3], and are briefly summarized here. UKB men and women were genotyped on two arrays. The first ~50,000 samples were genotyped on the UK BiLEVE array and the remaining ~450000 samples were genotyped on the UK Biobank Axiom array. Genotype data were imputed against two references panels: Haplotype Reference Consortium (HRC) panel and UK10K + 1000 Genomes panel. We used the imputed data released by UKB in March 2018, and applied in-house post-imputation quality controls (QC, i.e. genetic sex same as reported sex, XX or XY in sex chromosome and no outliers in heterozygosity and missing rates) [4]. Women of European descent with qualified genotype data were eligible for inclusion in our Mendelian randomization (MR) analyses (N=208,140, see Figure S1A).

Information on observed confounders were collected at the initial assessment. Age (years) was derived based on date of birth and date of attending an initial assessment centre. Women were asked to report their age when they had their only one child, or the first child if they had given birth to more than one child. Anthropometric measures included standing height (cm) and body mass index (BMI, kg/m$^2$, constructed from standing height and weight). Participants were also asked to report (1) the numbers of their live-born babies and stillborn babies (if they had any pregnancy loss), and we derived parity by adding these numbers together; (2) the qualifications they achieved, and we grouped their highest qualifications into three levels: O levels/GCSEs or equivalent and below, A levels/AS levels or equivalent, and College or university degree; (3) their smoking status (current/former/never), and we derived a binary ever versus never measure of smoking status by combining current and former smokers; (4) frequency of drinking alcohol at six levels from ‘never’ to ‘daily or almost daily’, and we derived a binary variable for the comparison between cohorts.

To calculate the associations of maternal single nucleotide polymorphisms (SNPs) with our exposures and outcomes, we used a split-sample strategy to mitigate winner’s curse bias [5, 6]. We calculated SNP-sleep duration associations in both the whole sample and each split sample using linear regression, adjusting for genotyping batch, women’s age and top 40 genetic principal components (PCs). We calculated SNP-outcomes associations in each split sample using logistic regression (linear regression for offspring birthweight (BW)), adjusting for the same covariates.

***Avon Longitudinal Study of Parents and Children (ALSPAC)***

Pregnant women resident in Avon, UK with expected dates of delivery 1st April 1991 to 31st December 1992 were invited to take part in the study [7, 8]. The initial number of pregnancies enrolled is 14,541 (for these at least one questionnaire has been returned or a 'Children in Focus' clinic data had been attended by 19/07/99) [7, 8]. Of these initial pregnancies, there was a total of 14,676 foetuses, resulting in 14,062 live births and 13,988 children who were alive at 1 year of age [7, 8]. Our study relied on 13,867 pregnancies from a total of 13,761 women, and most pregnancies were recruited at the first antenatal clinic visit in the first trimester of pregnancy [7]. Questionnaires were sent at regular intervals during pregnancy, and biological samples were taken from parents and children including blood samples from which DNA was extracted. The study website (http://www.bristol.ac.uk/alspac/) provides details of all available phenotypes via a searchable data dictionary. Ethical approval for the study was obtained from the ALSPAC Ethics and Law Committee and the Local Research Ethics Committees. Consent for biological samples has been collected in accordance with the Human Tissue Act (2004). Informed consent for the use of data collected via questionnaires and clinics was obtained from participants following the recommendation of the ALSPAC Ethics and Law Committee at the time.

Genotyping, pre-imputation quality control, and imputation procedures were described in detail elsewhere [9], and are briefly summarized here. ALSPAC mothers were genotyped using Illumina human660K quad SNP chip, and ALSPAC children were genotyped using Illumina HumanHap550 quad genome-wide SNP genotyping platform. Genotype data for both ALSPAC mothers and children were imputed against HRC v1.1 reference panel, after a similar QC procedure (minor allele frequency (MAF) ≥1%, a call rate ≥95%, in Hardy-Weinberg equilibrium (HWE), correct sex assignment, no evidence of cryptic relatedness, and of European decent). Women of European descent with qualified genotype data and live-born singleton offspring were eligible for inclusion in our MR analyses (N=6826, see Figure S1B).

Maternal age at delivery (years) was derived from date of delivery and date of birth. Maternal height and pre-pregnancy weight were self-reported at 12 weeks of gestation and were used to calculate BMI. Parity was defined as the number of pregnancies resulting in a live or stillbirth by the ALSPAC team [10], which was self-report at 18 weeks of gestation. Mothers were also asked to report their highest educational qualification, and we categorized the answers into three levels ("CSE/Vocational/O level", "A level" and "Degree"). We used self-reported tobacco smoking in the first three months of pregnancy and in the last two weeks at 18 weeks of gestation to construct a binary ever versus never smoked in pregnancy. Never smokers were women who did not smoke in both time periods, while ever smokers were women who smoked in either of them. Mothers reported their frequency of alcohol use in the first three months and in the last two months of pregnancy at six levels, and we also derived a binary ever versus never measure of alcohol consumption. Sex of offspring (ALSPAC children) was recorded. Both maternal and paternal occupational social class were reported by ALSPAC mothers at 32 weeks of gestation. They were defined using the UK Registrar General's occupational coding, and were coded from social class V (unskilled manual) to social class I (professional) [10]. We generated the household occupational social class based on whichever was the higher one.

To calculate the associations of SNPs with pregnancy and perinatal outcomes, we fitted logistic regression (linear regression for BW), adjusting for women's age and top 20 PCs. We further adjusted for fetal genotype in each regression model.

***Born in Bradford (BiB)***

BiB is a birth cohort that recruited 13,776 pregnancies to 12,453 women resident in the Bradford metropolitan district (a city in the North of England), with expected dates of delivery between 2007-2011 [11]. Most pregnancies were recruited during an oral glucose tolerance test (OGTT) undertaken between 24-28 weeks of gestation, and to which all pregnant women booked to give birth in

Bradford Royal Infirmary were invited [11]. BiB reflects the multicultural mix profile of Bradford, with approximately 50% of women being of South Asian descent [11]. The study website (https://borninbradford.nhs.uk/) provides details of all available data. Ethical approval for BiB was obtained from the Bradford Research Ethics Committee.

Genotyping, pre-imputation quality control, and imputation procedures were described elsewhere [12], and we briefly summarize here. Both BiB mothers and BiB children were genotyped using Illumina HumanCoreExome chip. Genotype data for both of them were imputed against UK10K + 1000 Genomes reference panel, after a similar QC procedure (a call rate ≥99.5%, correct sex assignment, no evidence of cryptic relatedness, correct ethnicity assignment). Two subsets of individuals were declared based on a combination of principal component analysis and self-reported ethnicity. To combine the MR estimates with those in other cohorts, only women of European descent with qualified genotype data and live-born singleton offspring were eligible for inclusion in our analyses (N=2940, see Figure S1C).

Information on maternal age (years), height (cm) and the highest educational qualification were collected via a questionnaire at baseline. The BiB research team derived maternal BMI (using a weight measured at pregnancy booking around 12 weeks of gestation), and categorized the highest educational qualification into 7 groups ("<5 GCSE equivalent", "5 GCSE equivalent", "A-level equivalent", "Higher than A-level", "Other", "Don't know" and "Foreign unknown"). In the baseline questionnaire, mothers were asked to report the number of cigarettes they smoked in the first three months of pregnancy and since the fourth month of pregnancy, and a binary ever versus never measure of smoking status in pregnancy was derived. Mothers were also asked to report the frequency of drinking alcohol 3 months before pregnancy, in the first three months of pregnancy and since the fourth month of pregnancy at four levels ("Yes, once a week", "Yes, occasionally", "Yes, not specified" and "No"), from which a binary alcohol consumption was derived. Parity and sex of offspring were extracted from the Eclipse pregnancy record and Eclipse electronic maternity record, respectively.

To calculate the associations of SNPs with pregnancy and perinatal outcomes, we fitted logistic regression (linear regression for BW), adjusting for women's age and top 10 PCs. We further adjusted for fetal genotype in each regression model.

***Norwegian Mother, Father and Child Cohort Study (MoBa)***

MoBa is a population-based pregnancy cohort study conducted by the Norwegian Institute of Public Health. Participants were recruited from all over Norway from 1999-2008 [13]. The women consented to participation in 41% of the pregnancies. The cohort now includes 114,500 children, 95,200 mothers and 75,200 fathers [13]. The current study is based on version 12 of the quality-assured data files released for research on "Prenatal environmental exposures and pregnancy outcomes-Mendelian randomization analysis". The establishment of MoBa and initial data collection was based on a license from the Norwegian Data Protection Agency and approval from The Regional Committees for Medical and Health Research Ethics. The MoBa cohort is now based on regulations related to the Norwegian Health Registry Act. The current study was approved by The Regional Committees for Medical and Health Research Ethics. The Medical Birth Registry of Norway (MBRN), established in 1967, is a national health registry containing information about all births in Norway. MoBa has been linked to the MBRN using unique personal identification numbers [13].

Blood samples were obtained from both parents during pregnancy and from mothers and children (umbilical cord) at birth [14]. Genotyping, pre-imputation quality control, and imputation procedures were described in detail elsewhere [15, 16], and we briefly summarized here. There were five projects contributed to MoBa genetics 1.0 [17], and we had to use an earlier version consisting of two projects – HARVEST and ROTTERDAM1 given their complete QC procedure. In HARVEST, MoBa mothers and children were genotyped using either Illumina HumanCoreExome12v1.1 or Illumina HumanCoreExome24v1.0. In ROTTERDAM1, MoBa mothers and children were genotyped using Illumina GSAMDv1.0. Genotype data from all batches were imputed against HRC v1.1 reference panel, after a similar QC procedure (MAF ≥5%, a call rate ≥99.2%, in HWE, correct sex assignment and no evidence of cryptic relatedness). Women of European descent with qualified genotype data and live-born singleton offspring were eligible for inclusion in our analyses (N=14,584, see Figure S1D).

Maternal age at delivery and parity were recorded in MBRN. Five women were grouped as "less than 17 years", and we recoded them as "NA". Women having more than four previous deliveries were recorded as "4" by the MoBa team. Maternal weight (when they became pregnant), height and the highest education attainment were self-reported at 15 weeks of gestation. We excluded implausibly extreme values (weight less than 30 kg or more than 200 kg, height shorter than 135 cm), and then constructed maternal BMI. Both maternal and paternal education attainments were reported by women at their 15 weeks of gestation. Each of them was grouped into three levels (9-year secondary school, high school, college or university degree). Both maternal and paternal smoking status in pregnancy, and maternal usage to other kinds of nicotine (chewing tobacco/snuff, chewing gum, adhesive patch, and inhaler) in pregnancy were reported by women at both 15 and 30 weeks of gestation. For each time point, we derived a binary ever versus never measure of smoking status by combining "daily" and "sometimes" smoking. Ever smokers (users) in pregnancy were participants who smoked (used other kinds of nicotine) at either 15 or 30 weeks of gestation, while never smokers (users) were those who did not at both time points. Maternal frequencies of alcohol consumption during 0-12, 13-24 and 25-30 weeks of gestation were self-reported at 30 weeks of gestation, with seven levels from "never" to "roughly 6-7 times a week", and we also derived a binary ever versus never measure of alcohol consumption during 0-30 weeks of gestation. Sex of offspring was also recorded in MBRN, and where records showed "not specified" or "uncertain" we recoded them as "NA".

To calculate the associations of SNPs with pregnancy and perinatal outcomes, we fitted logistic regression (linear regression for BW), adjusting for genotyping batch, women's age and top 10 PCs. We further adjusted for fetal genotype in each regression model. We meta-analysed SNP-outcome associations from all three birth cohorts (i.e. ALSPAC, BiB and MoBa) using fixed effects with inverse variance weights.

***FinnGen***

FinnGen is the national wide network of Finnish biobanks, including Auria Biobank, Biobank Borealis of Northern Finland, Biobank of Eastern Finland, Central Finland Biobank, Finnish Red Cross Blood Service Biobank, Finnish Clinical Biobank Tampere, Helsinki Biobank, Terveystalo Biobank, and THL Biobank [18]. Those biobanks were linked to national registries, including Drug purchase and Drug Reimbursement, Digital and Population Data Services Agency, Statistics Finland, Register of primary health care visits: AVOHILMO, Care Register for Health Care: HILMO, and Finnish cancer registry. Clinical endpoints were defined based on ICD-10, and the equivalent in ICD-8 and ICD-9. The Coordinating Ethics Committee of the Helsinki and Uusimaa Hospital District has approved the FinnGen consortium (Nr HUS/990/2017), and the ethical approval of each individual study has been described in detail elsewhere [19].

FinnGen participants were genotyped with Illumina and Affymetrix chip arrays. Genotype data was imputed against SISu v3 reference panel (http://sisuproject.fi), after a QC procedure (minor allele count ≥3, a call rate ≥98%, in HWE no outliers in heterozygosity, correct sex assignment, and of Finnish ancestry).

The FinnGen team released summary-level data of genome-wide association studies of more than 4000 clinical endpoints (https://www.finngen.fi/en/researchers/clinical-endpoints) at R5 wave (N=218,792 men and women). We extracted female-specific associations of the 78 SNPs with miscarriage (https://risteys.finngen.fi/phenocode/O15_ABORT_SPONTAN), gestational diabetes (https://risteys.finngen.fi/phenocode/GEST_DIABETES), hypertensive disorders of pregnancy (https://risteys.finngen.fi/phenocode/O15_GESTAT_HYPERT), and preterm birth (https://risteys.finngen.fi/phenocode/O15_PRETERM). These SNP-outcome associations were generated using SAIGE (mixed-effects logistic regression) [20], adjusting for genotyping batch, top 10 PCs, and women's age.

**Text S2. Technical considerations of one-sample MR to explore potential nonlinear effects**

The largest and most updated GWAS identified 78, 27 and 9 genome-wide significant SNPs for sleep duration (a discrete variable in the unit of 1-hour), short duration (a binary variable, defined as ≤6 hours/day) and long duration (a binary variable, defined as ≥9 hours/day), respectively [21]. We did not propose those 27 and 9 SNPs as IVs because their two-sample MR estimates could be more vulnerable to weak instrument bias [22]. Given short and long duration are binary variables, the original GWAS identified their SNPs via logistic regression, and thus reported odds ratios. However, odds ratios are non-collapsible, and it would be difficult to interpret two-sample MR estimates for binary exposures [23].

In one-sample MR, we followed a previous study [24], to calculate linear MR estimates for sleep duration on pregnancy and perinatal outcomes within each group of residual sleep duration using the ratio of coefficients method [25]. Specifically, linear regression was used to calculate the association of genetic risk score (GRS) with observed sleep duration, and then obtain the regression coefficient $\beta_X$ and its standard error $SE_X$. Logistic regression was used to calculate associations of GRS with binary outcomes, and linear regression was used for BW. We obtained the regression coefficient $lnOR_Y$ ($\beta_Y$ for BW) and their standard error $SE_Y$. Both the exposure and the outcome models were adjusted for genetic array, women's age and top 40 PCs. Within each group, the one-sample MR estimate was calculated as the Wald ratio $\frac{\beta_Y}{\beta_X}$, and its standard error was calculated as $\frac{SE_Y}{\beta_X}$.

**Figure S1. Flow chart of each cohort**

*A. UK Biobank (UKB)*

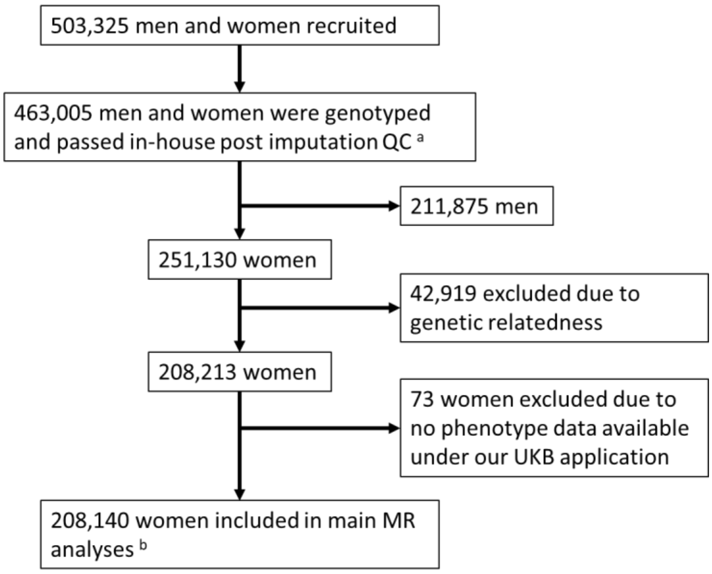


*B. Avon Longitudinal Study of Parents and Children (ALSPAC)*

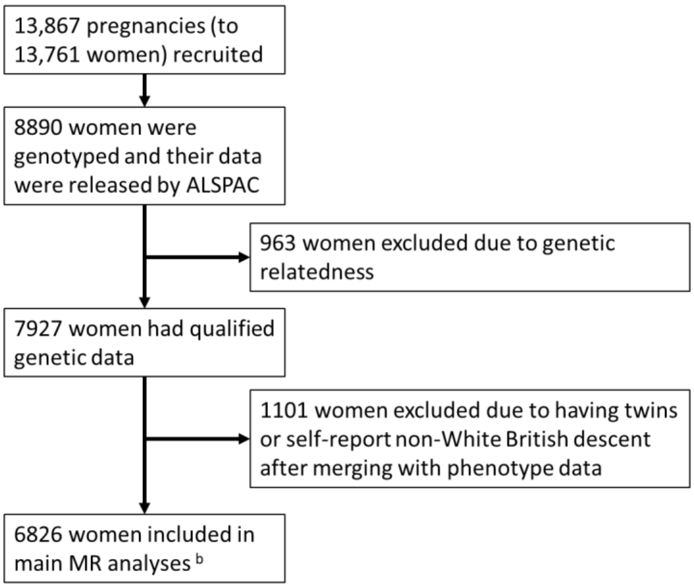

#### C. Born in Bradford (BiB)

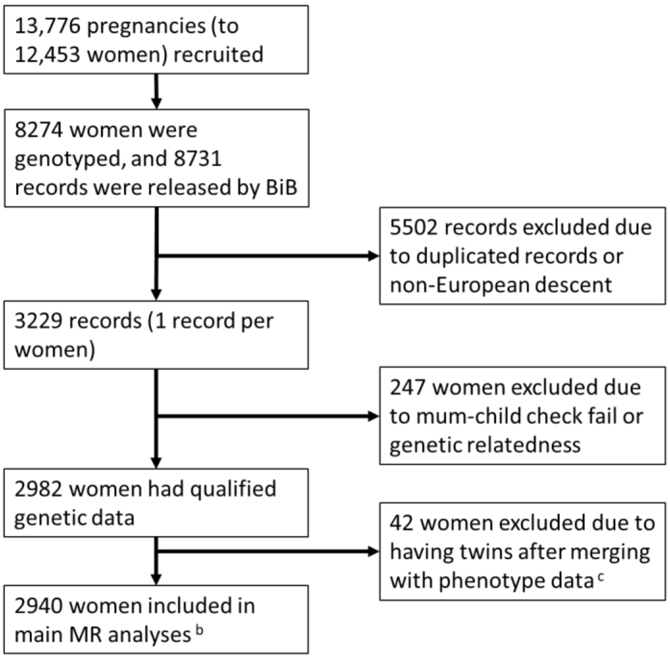


#### D. Norwegian Mother, Father and Child Cohort Study

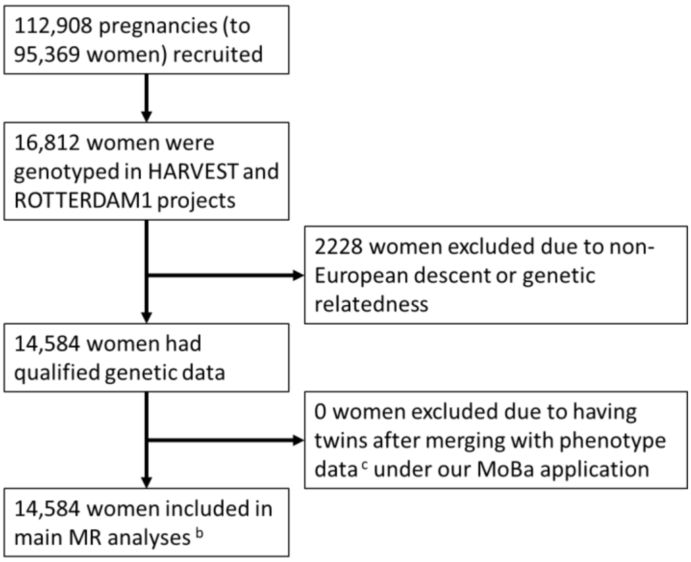


[a] We used the imputed data released by UKB in March 2018, and applied in-house post-imputation QC [4].
[b] The numbers of cases and controls for each pregnancy and perinatal outcomes are listed in Table 1.
[c] We randomly selected one pregnancy per woman in the phenotype data if multiple pregnancies were recorded.
Abbreviation: MR, Mendelian randomization, MR; QC, Quality control.

**Figure S2. Histogram of residual sleep duration in UK Biobank women**

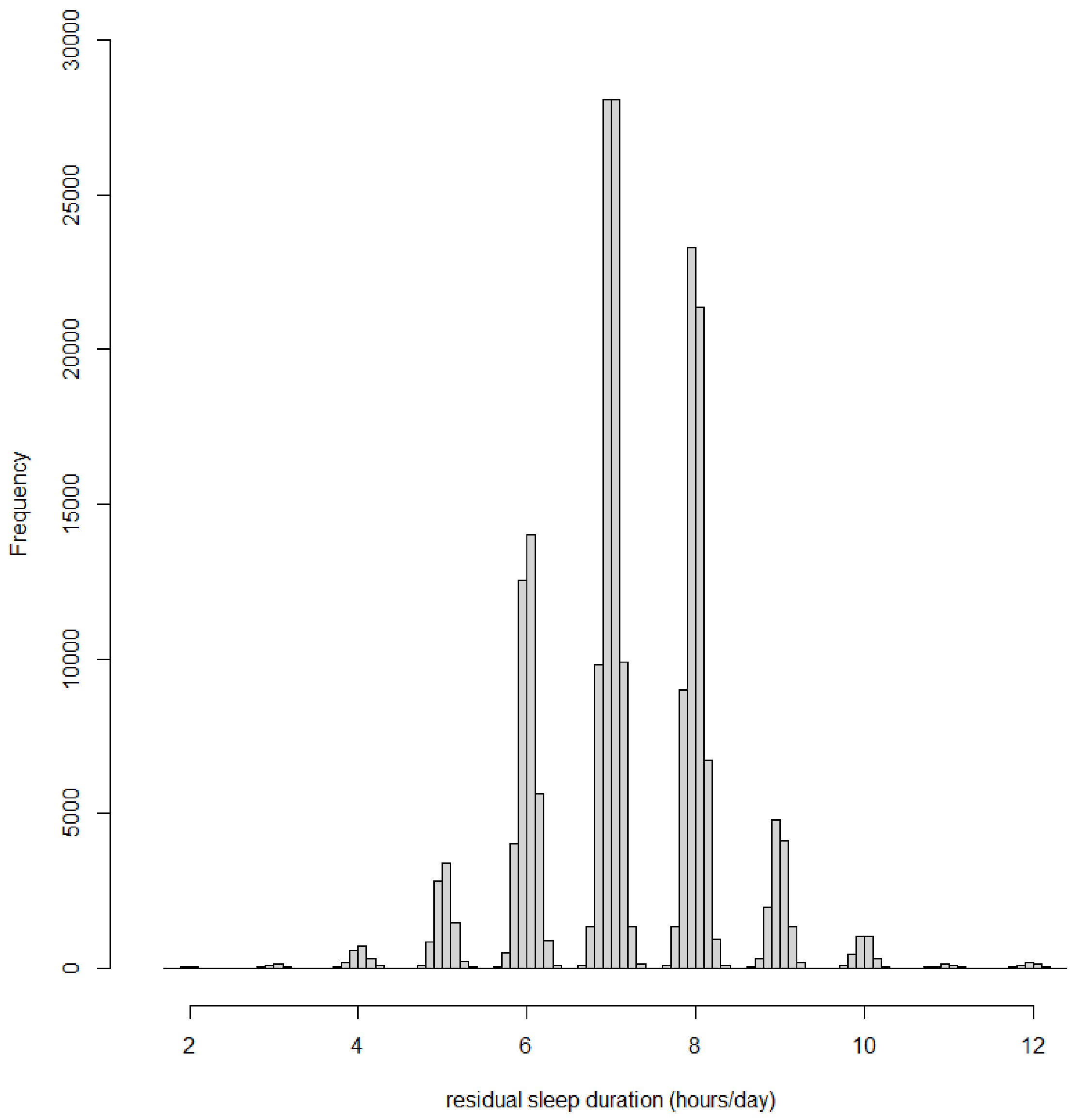


The X-axis is the residual sleep duration, with a mean of 7.183 and a range from 1.765 to 12.330 hours/day, and the Y-axis is the number of women. The residual was calculated using the following formula: residual = observed sleep duration – (genetic contribution to sleep duration from the genetic risk score (GRS) – 7.183), where 7.183 is the mean of genetic contribution to sleep duration from the GRS [24].

**Figure S3. One-sample MR estimates of effects of sleep duration on pregnancy and perinatal outcomes in 206,500 UK Biobank women**

(A) Three groups of different duration lengths based on thresholds from existing literature [26, 27]

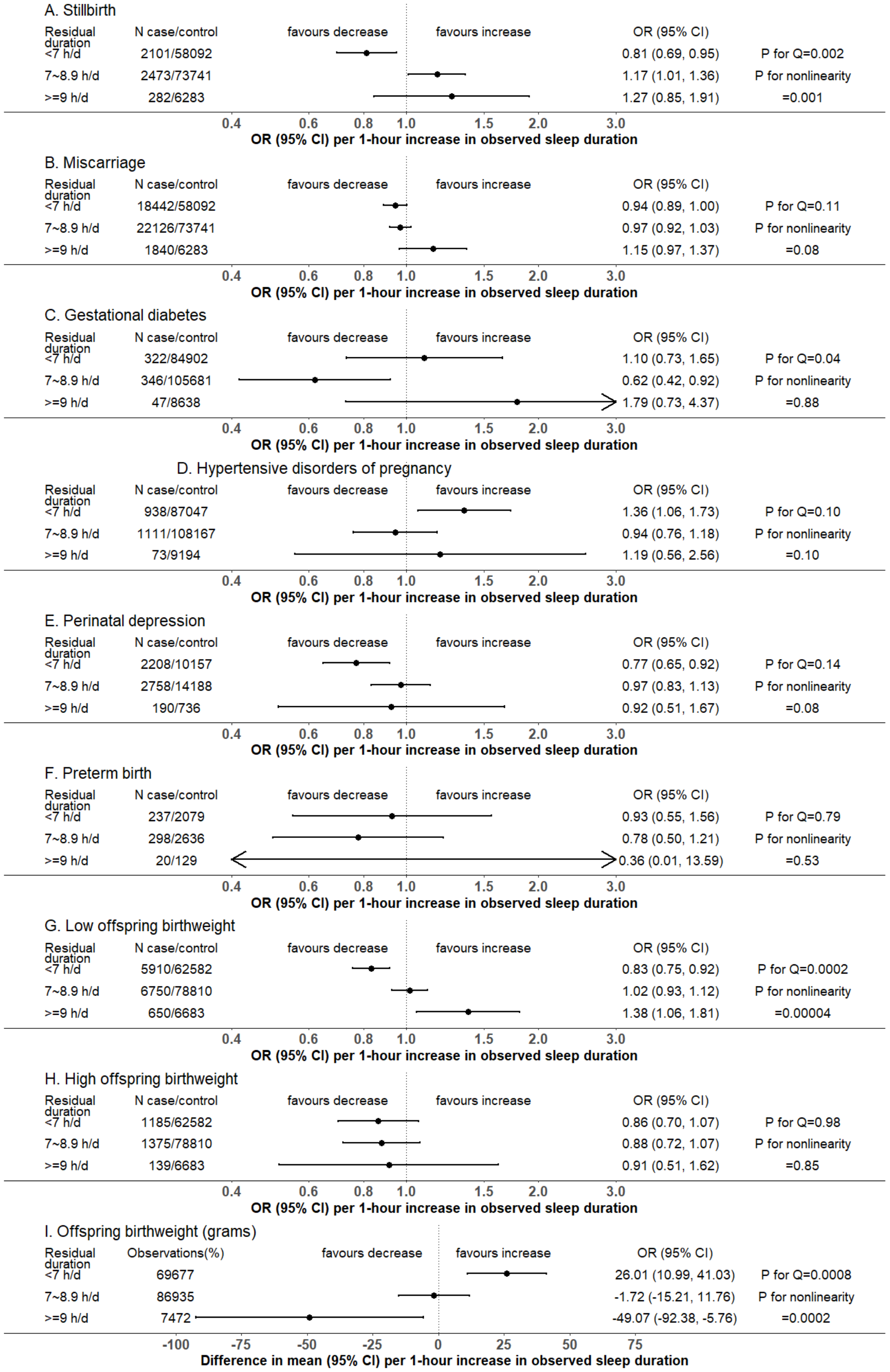

(B) In thirds of residual sleep duration

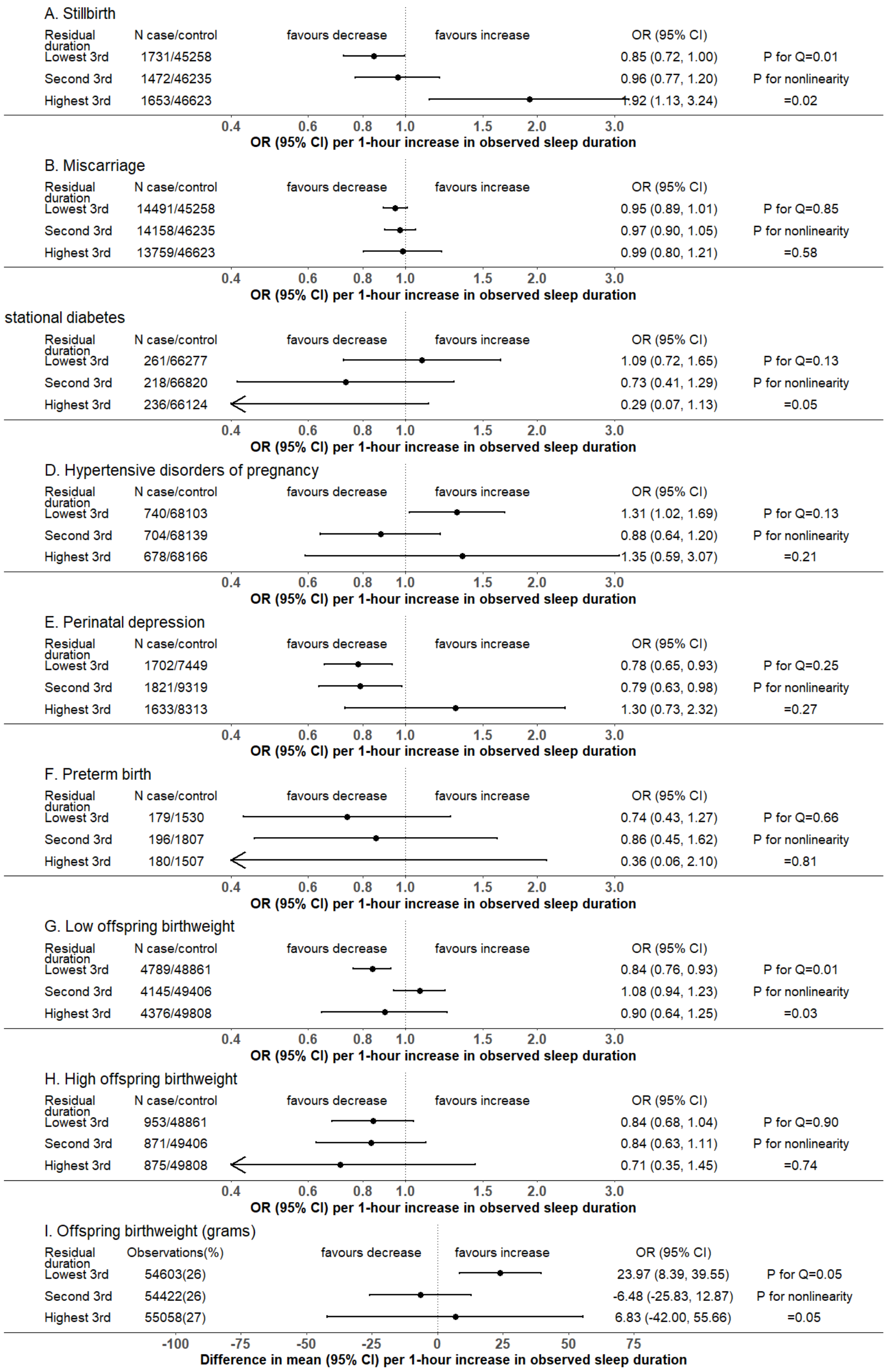


We present MR estimates of a linear effect of increasing duration on the outcome across the length of duration covered in each group. Further details about identifying the pattern of nonlinear effects are illustrated in Table 1.

P-value for Cochran's Q-statistic testing statistical evidence for between-group heterogeneity. P-value for nonlinearity testing statistical evidence whether the MR estimates are changed as the self-reported sleep duration mean increases.
Abbreviations: CI, confidence interval; h/d, hours/day; MR, Mendelian randomization; OR, odds ratio.

**Figure S4. Associations of 78 SNPs with pregnancy and perinatal outcomes with versus without adjustments of fetal genotypes in the combination of Avon Longitudinal Study of Parents and Children, Born in Bradford and Norwegian Mother, Father and Child Cohort Study (N=18,663)**

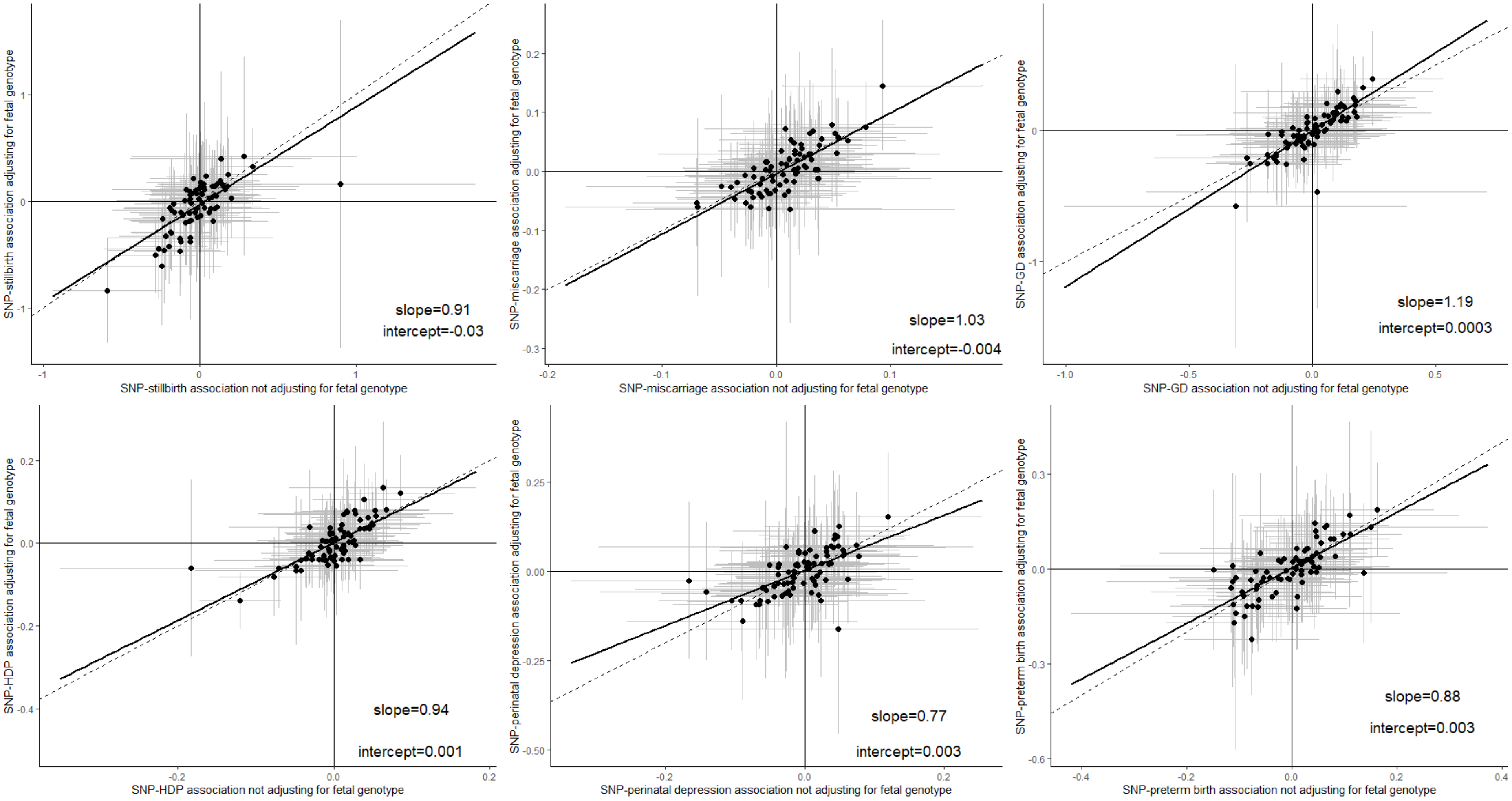

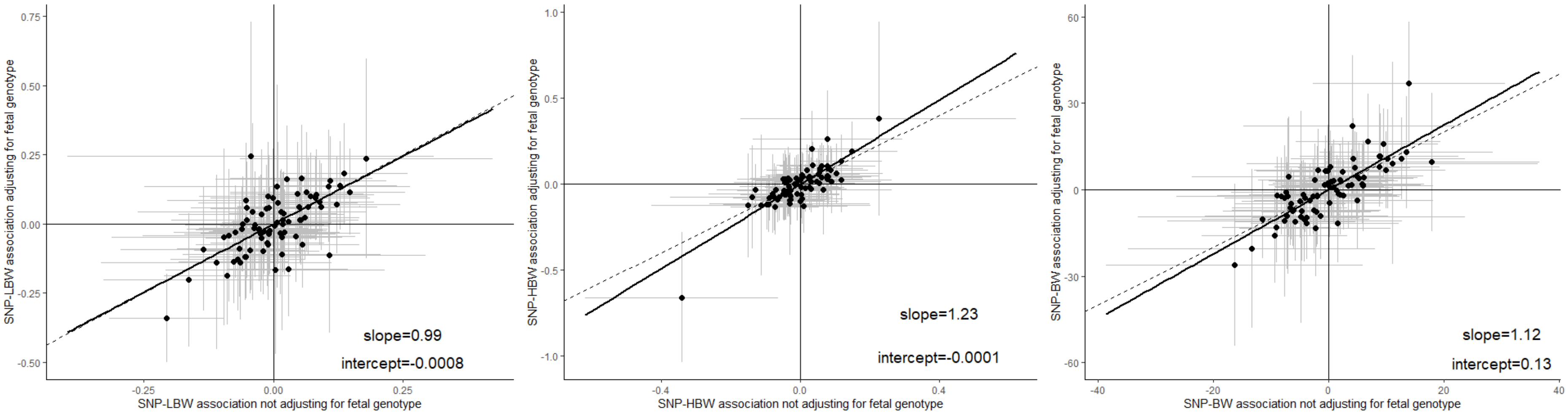


Black dots represent point estimates (i.e. ln[odds ratio]) of the SNP-outcome associations, and grey lines represent their 95% confidence intervals. Solid lines are linear regression lines of those black dots, and dash lines represent y=x.

Abbreviations: GD, gestational diabetes; HBW, high offspring birthweight; HDP, hypertensive disorders of pregnancy; LBW, low offspring birthweight; SNPs, single nucleotide polymorphisms.

**Figure S5. Leave-one-out analyses for sleep duration on pregnancy and perinatal outcomes in UK Biobank (datasets A on B)**

(A) Stillbirth

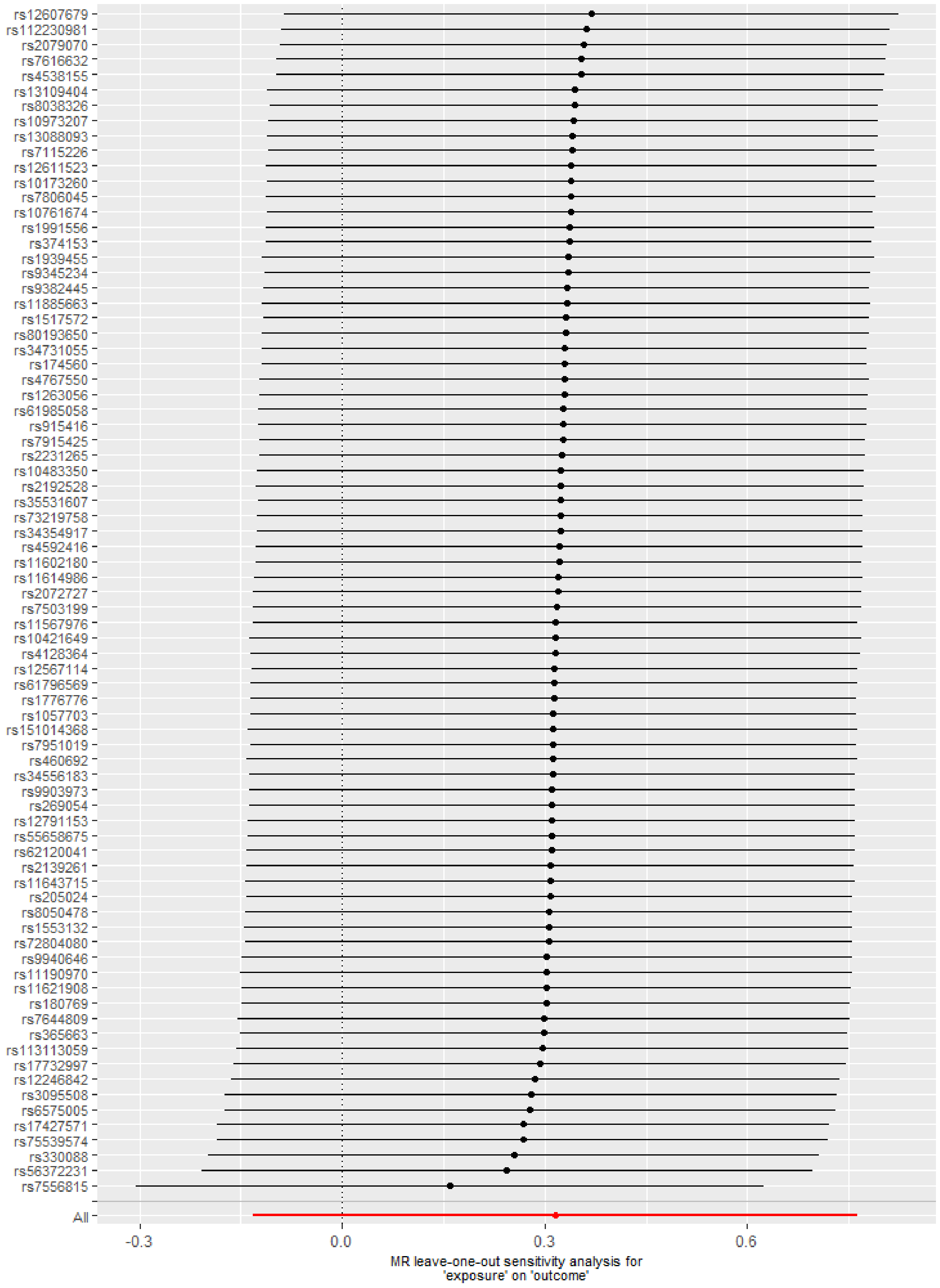

(B) Miscarriage

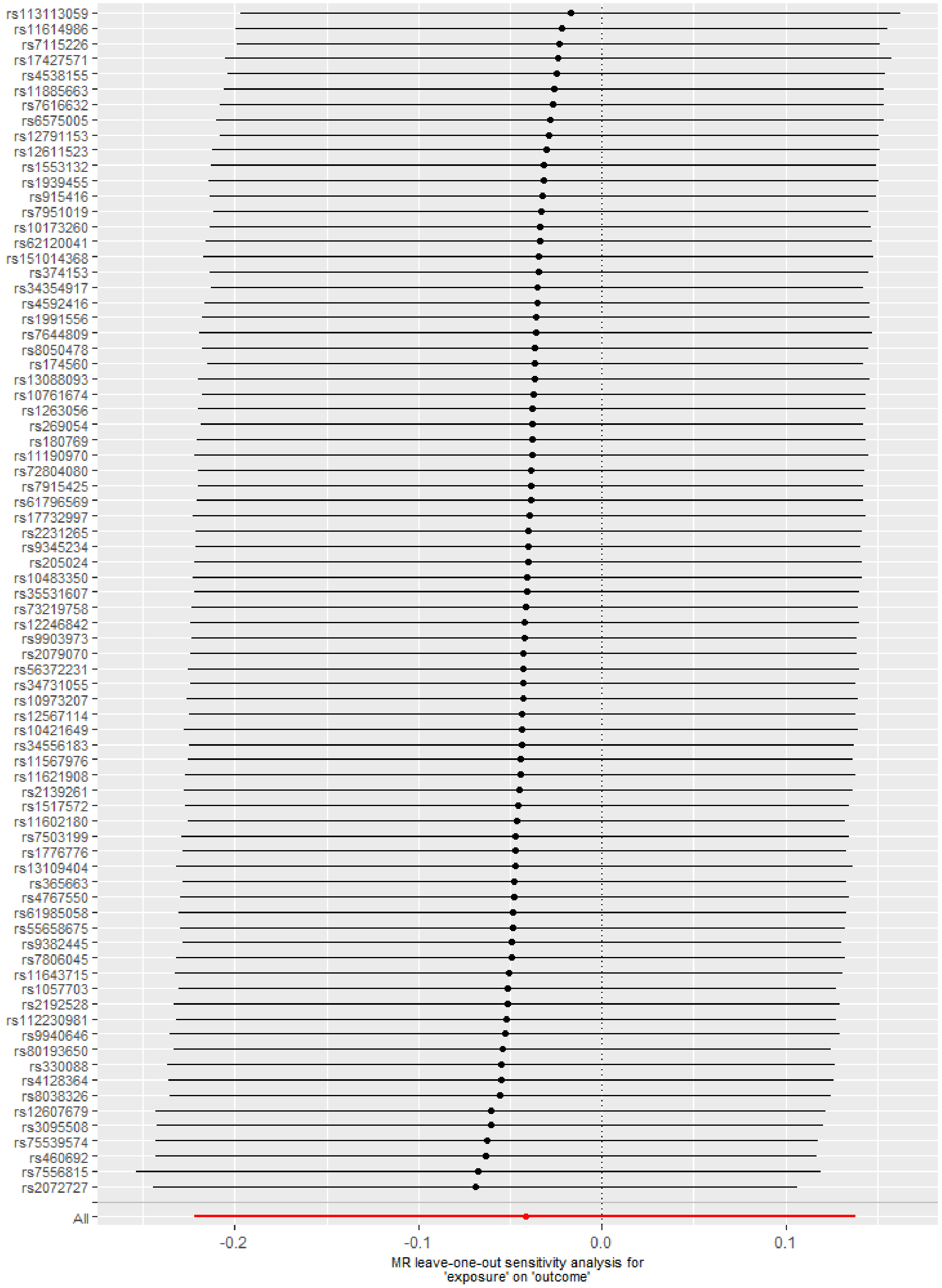

(C) Gestational diabetes

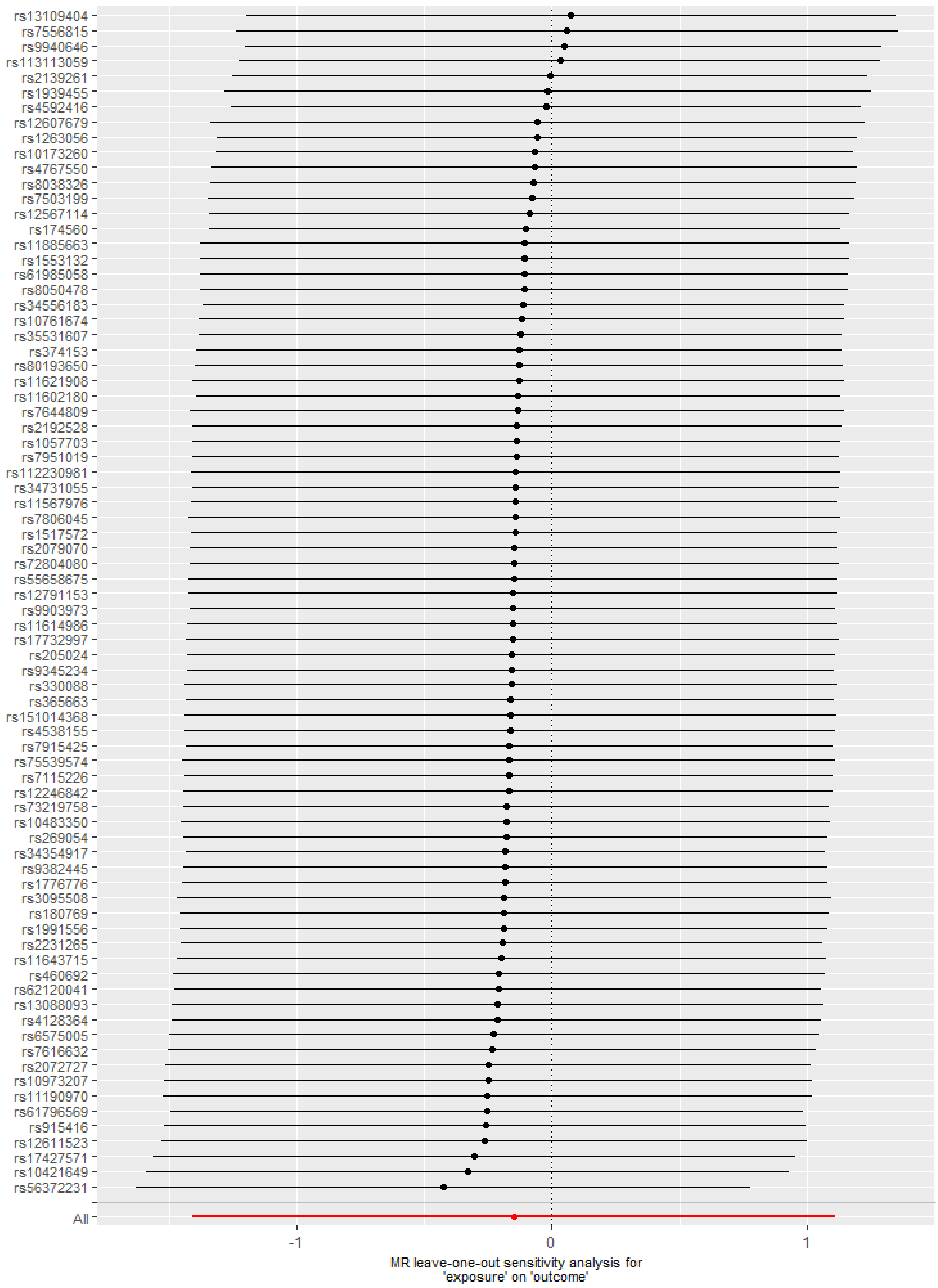

(D) Hypertensive disorders of pregnancy

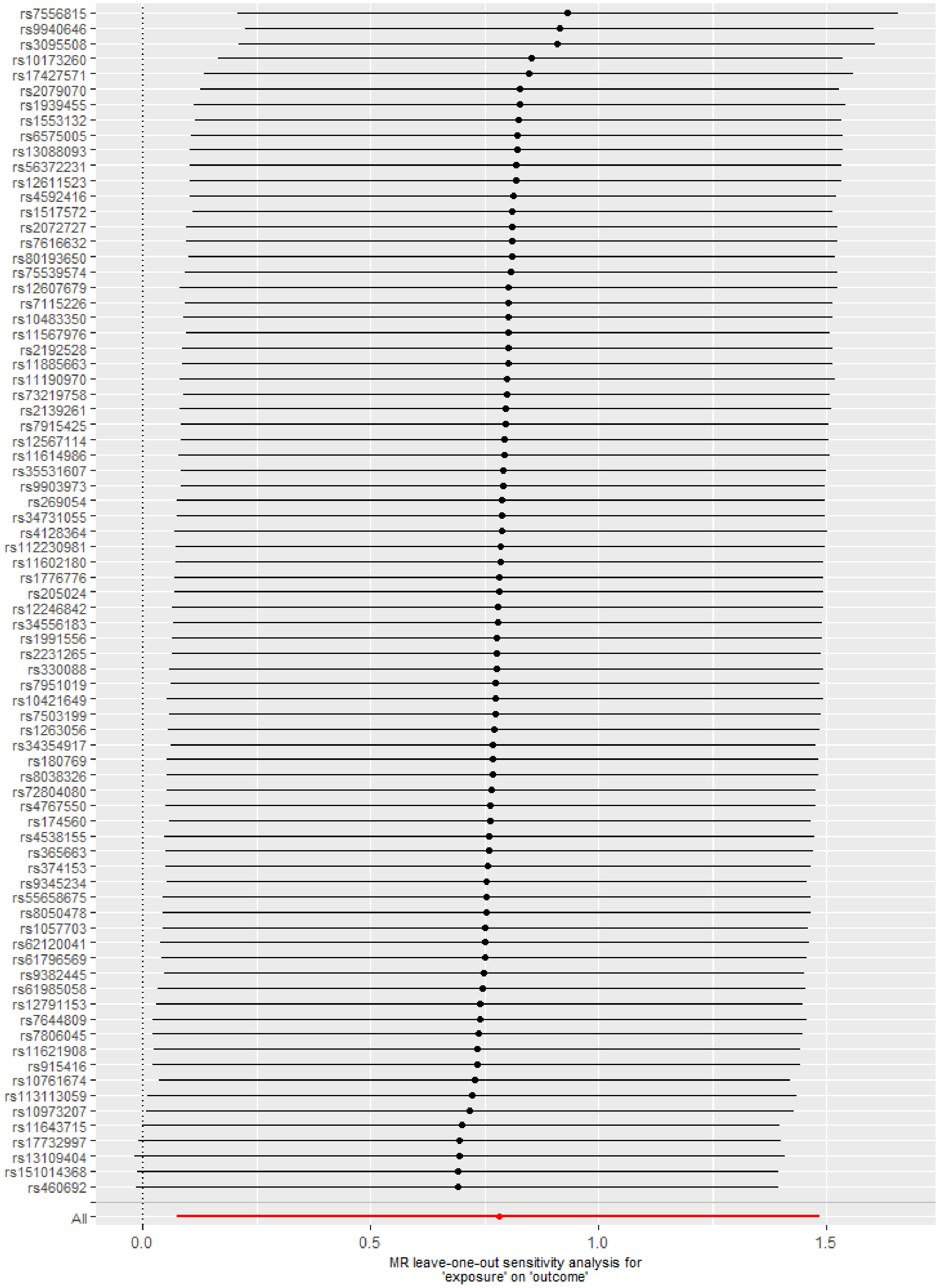

(E) Perinatal depression

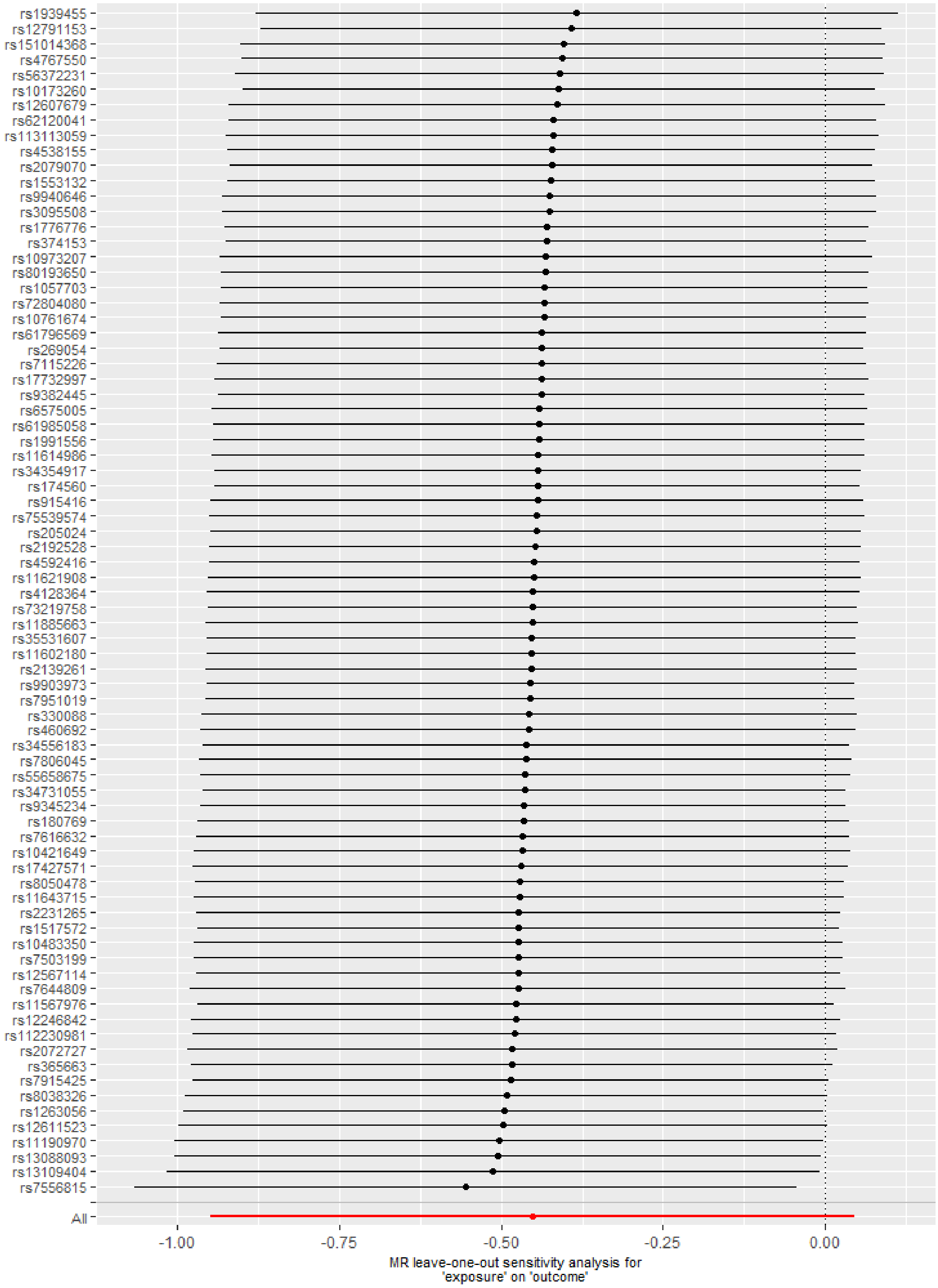

### (F) Preterm birth

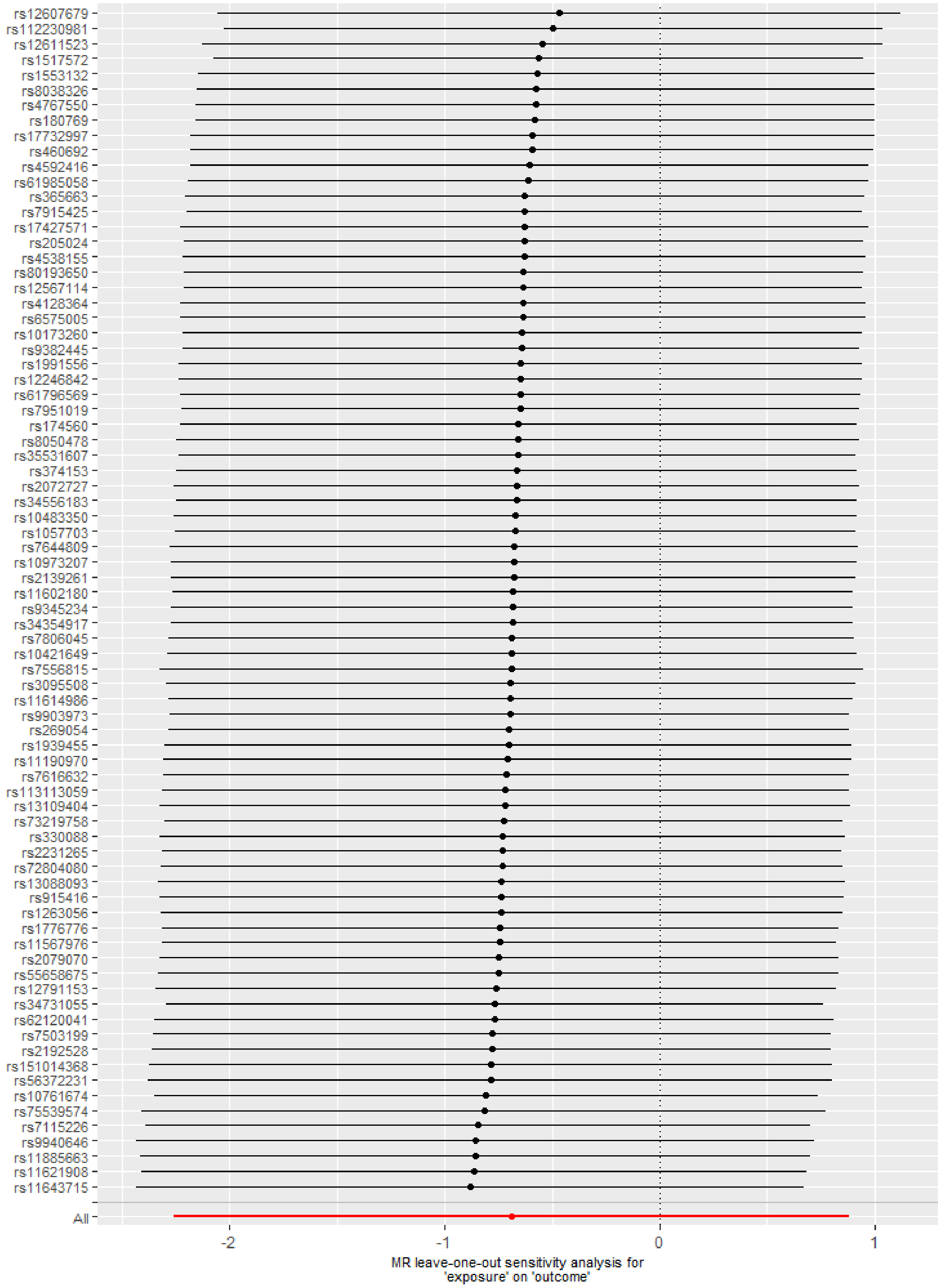

(G) Low offspring birthweight

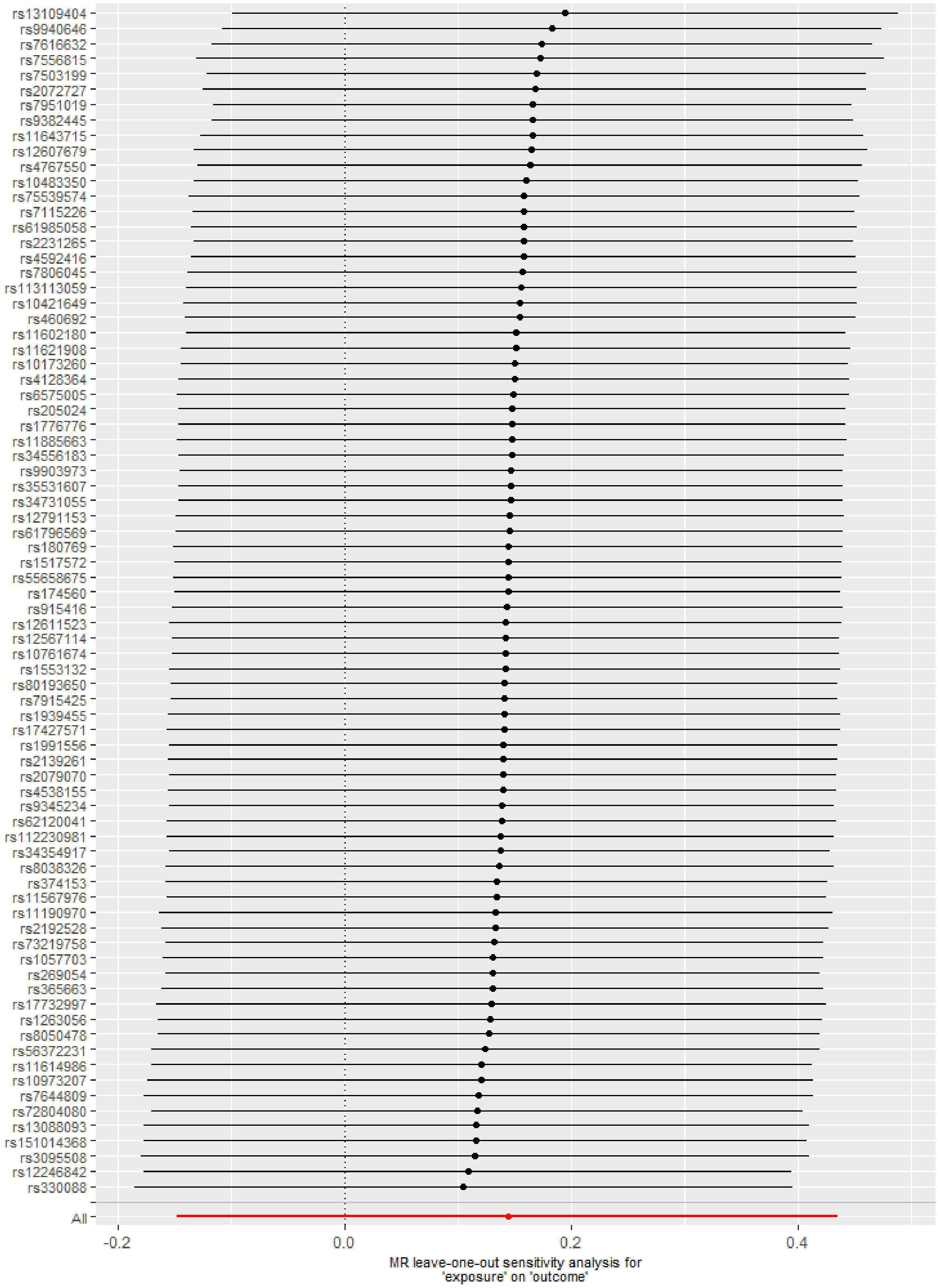

(H) High offspring birthweight

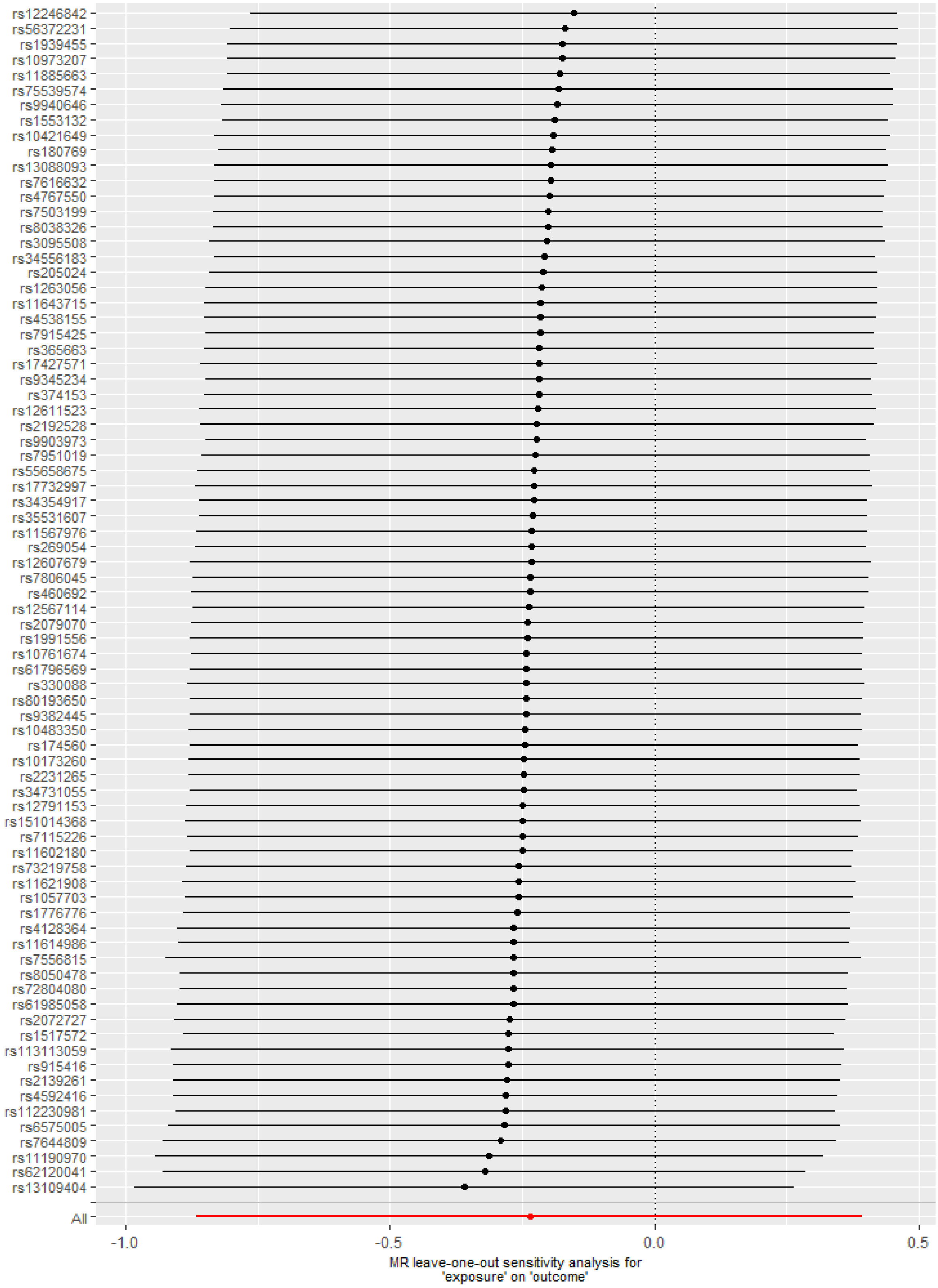

(I) Offspring birthweight (grams)

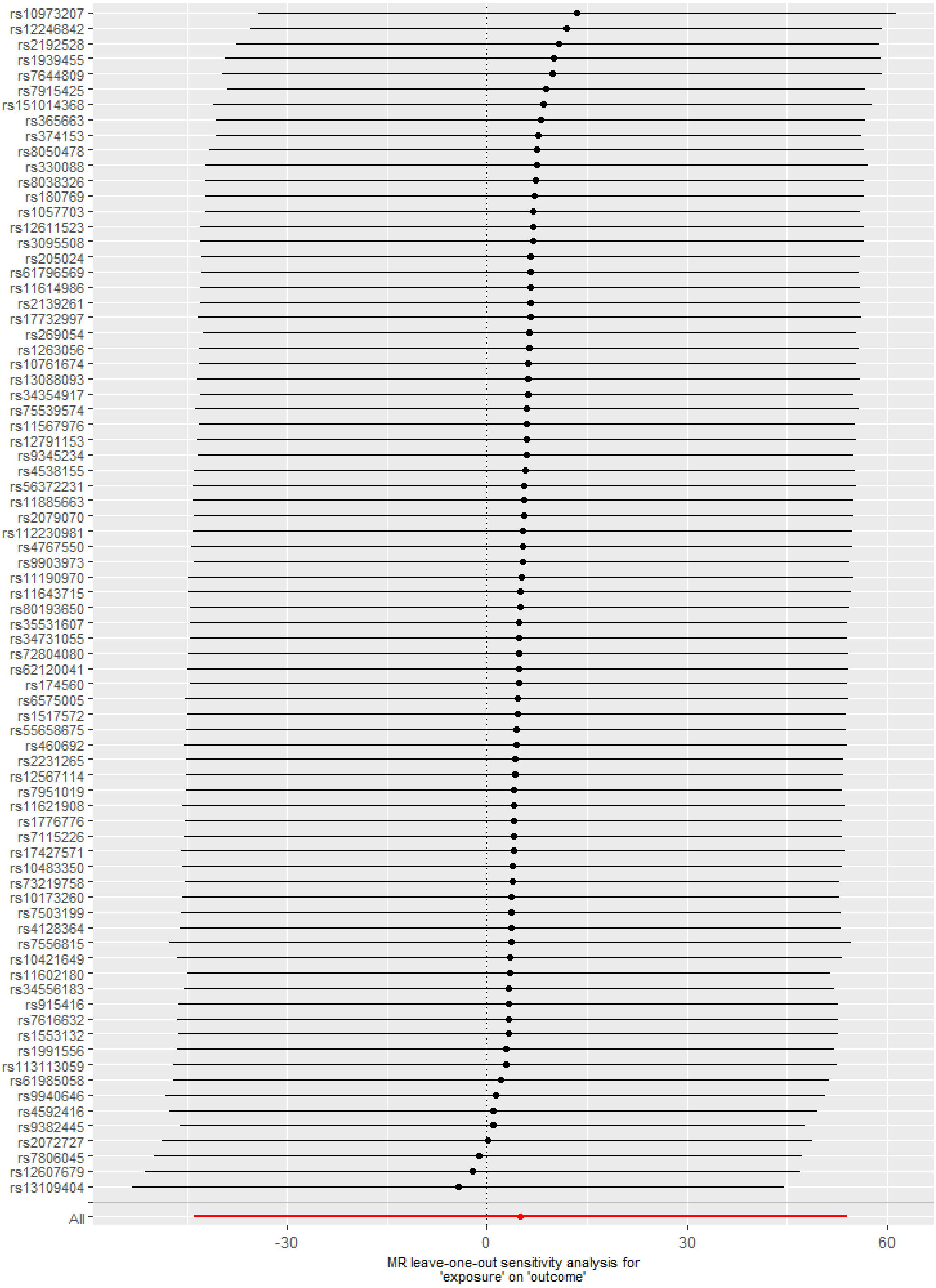

**Figure S6. Leave-one-out analyses for sleep duration on pregnancy and perinatal outcomes in UK Biobank (datasets B on A)**

(A) Stillbirth

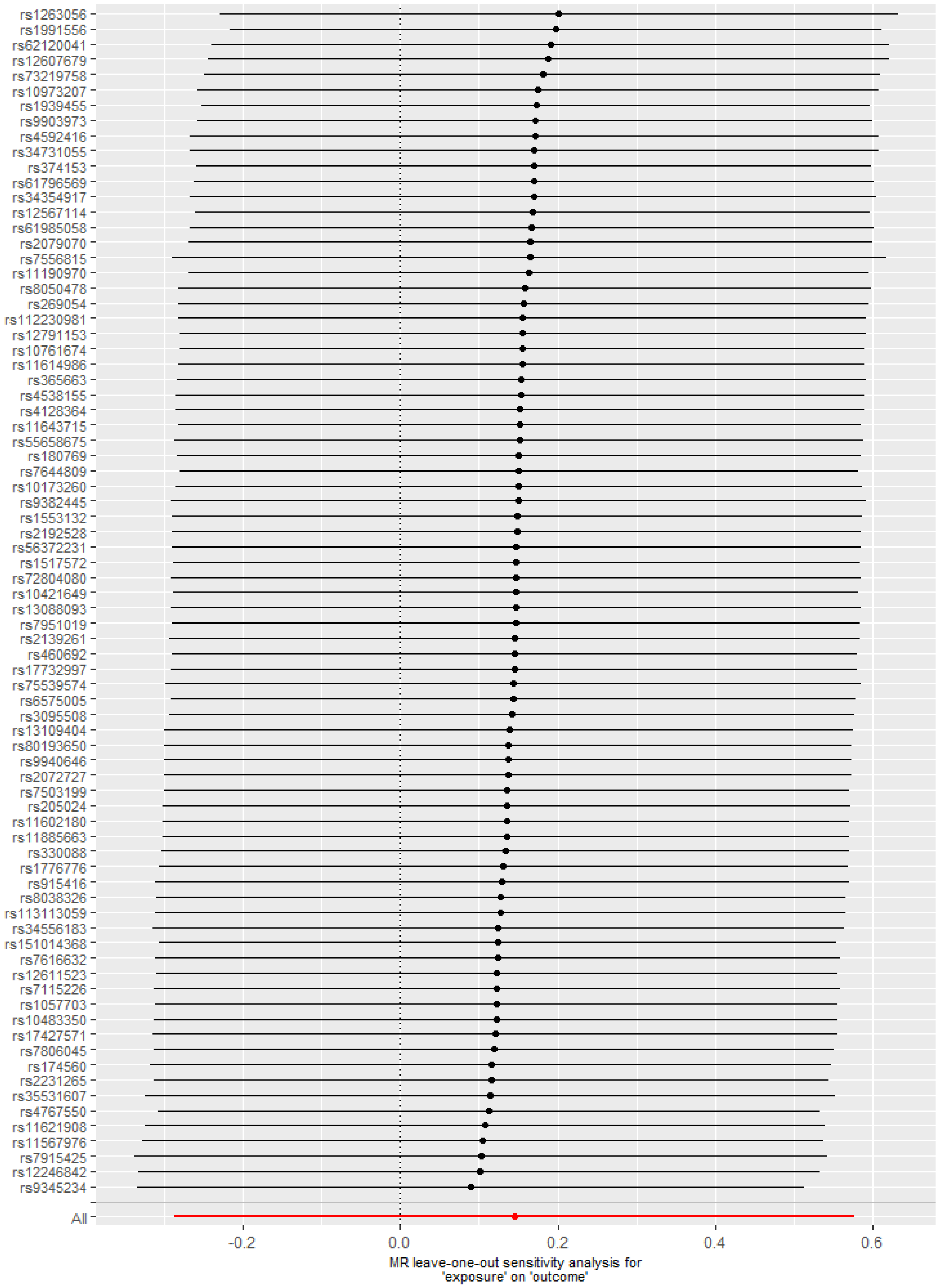

(B) Miscarriage

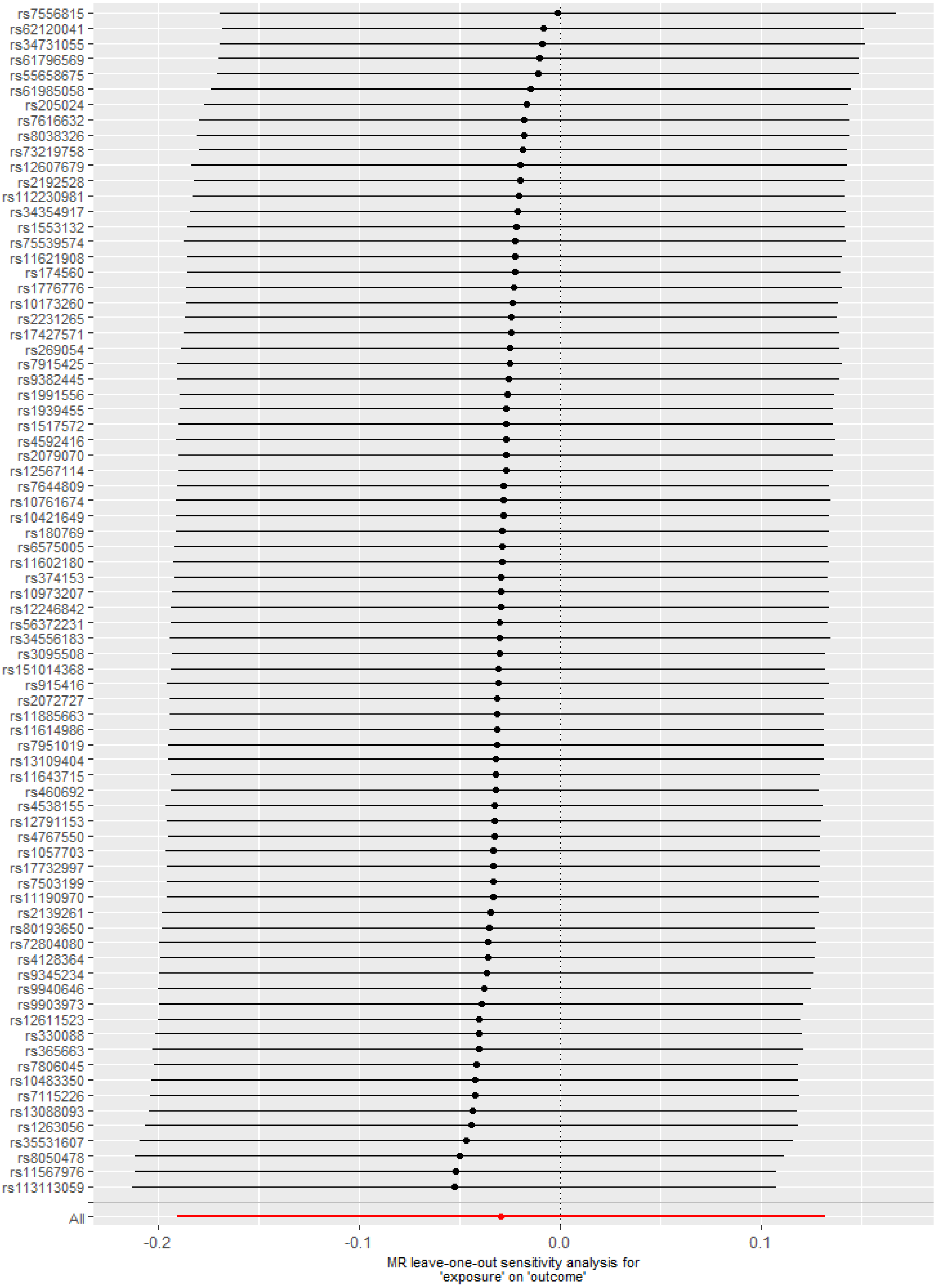

(C) Gestational diabetes

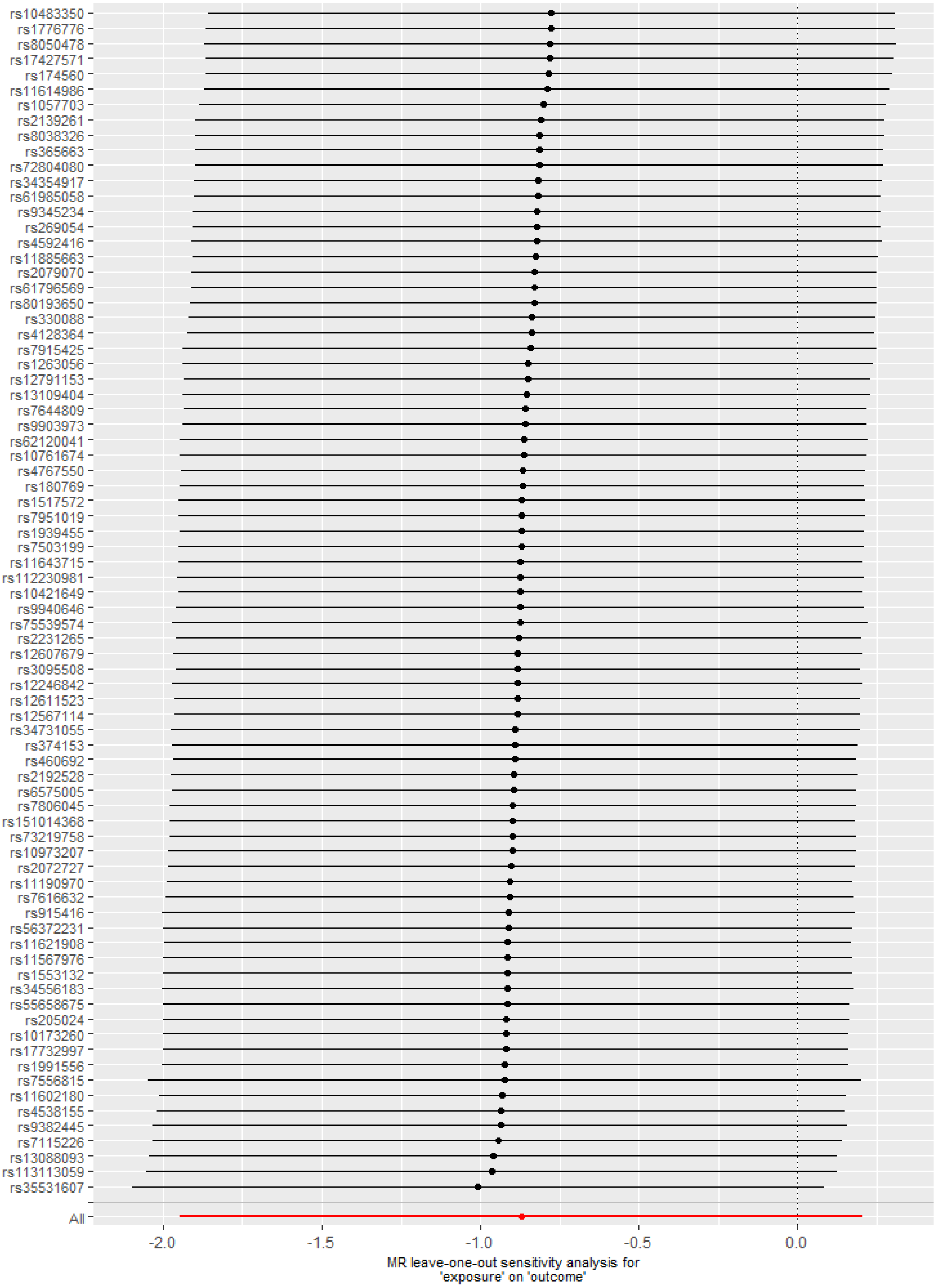

(D) Hypertensive disorders of pregnancy

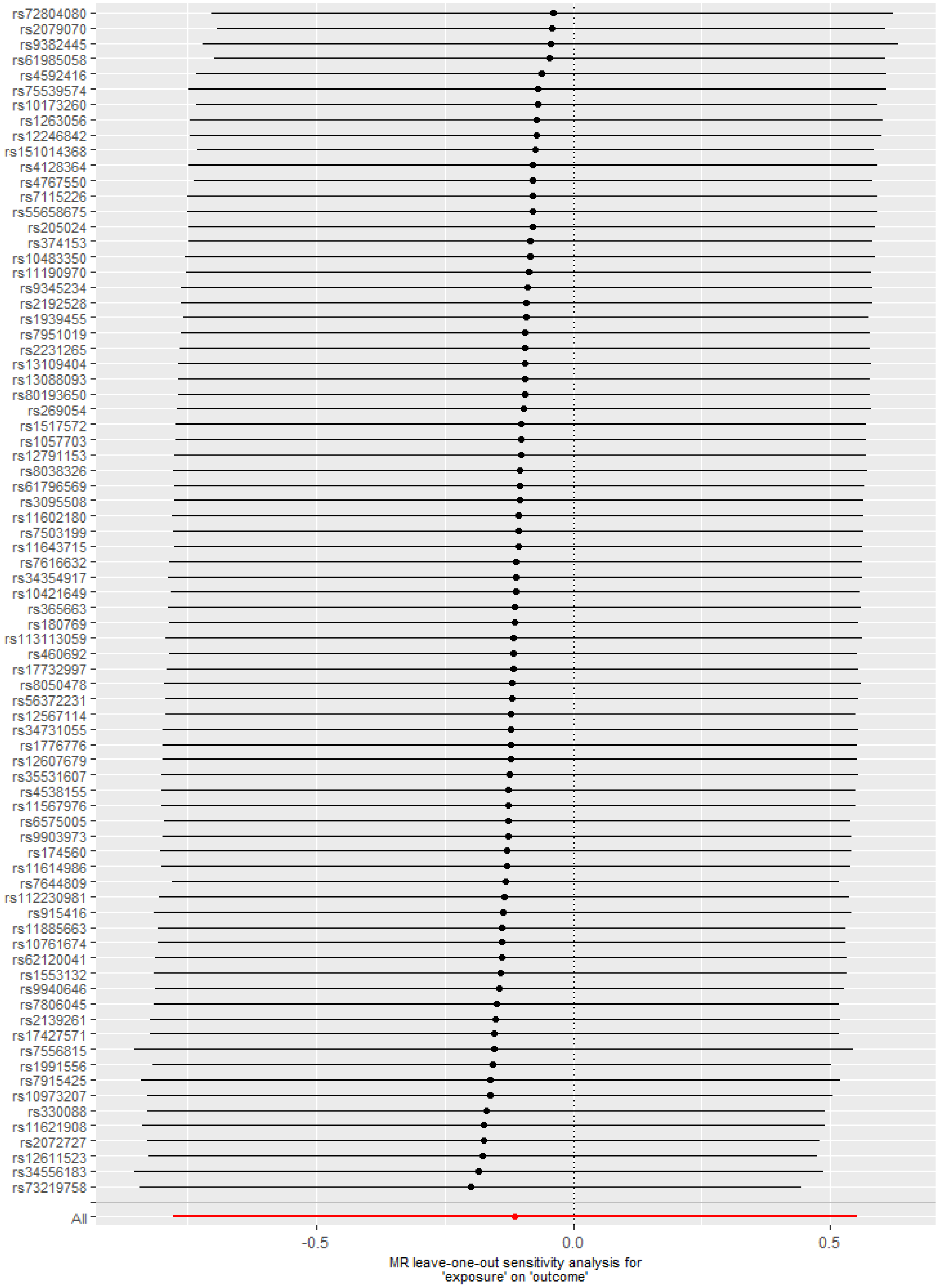

(E) Perinatal depression

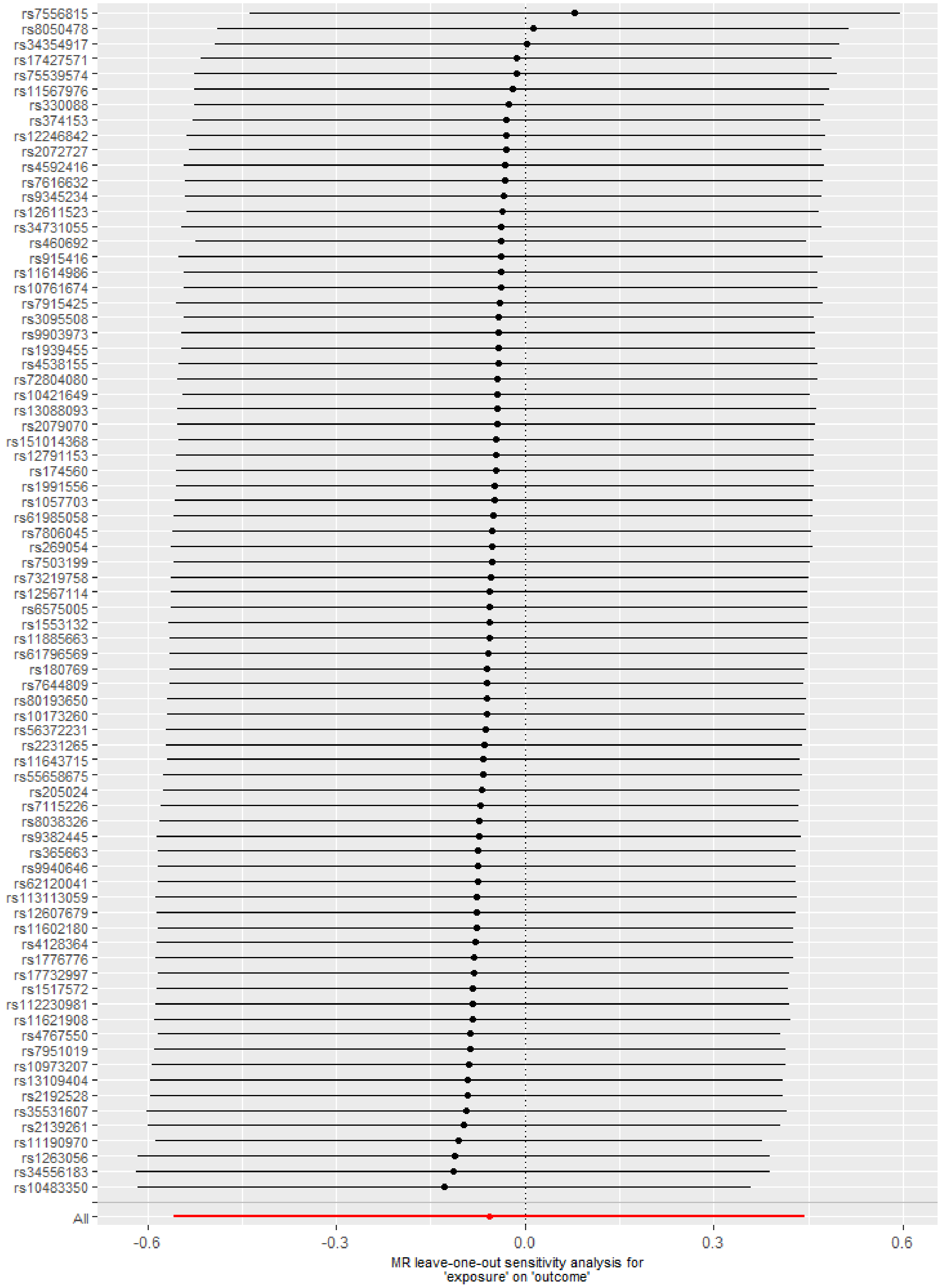

(F) Preterm birth

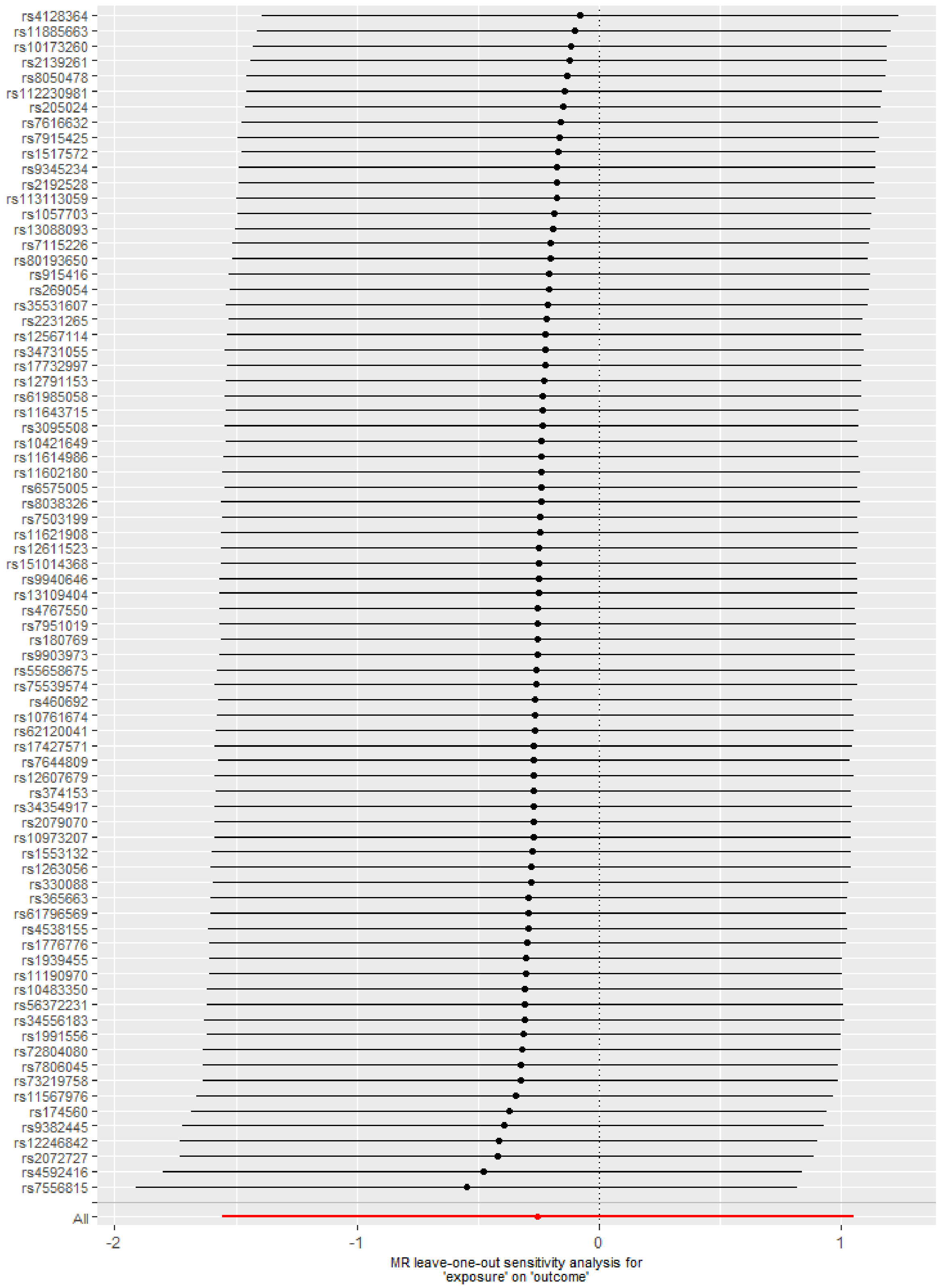

(G) Low offspring birthweight

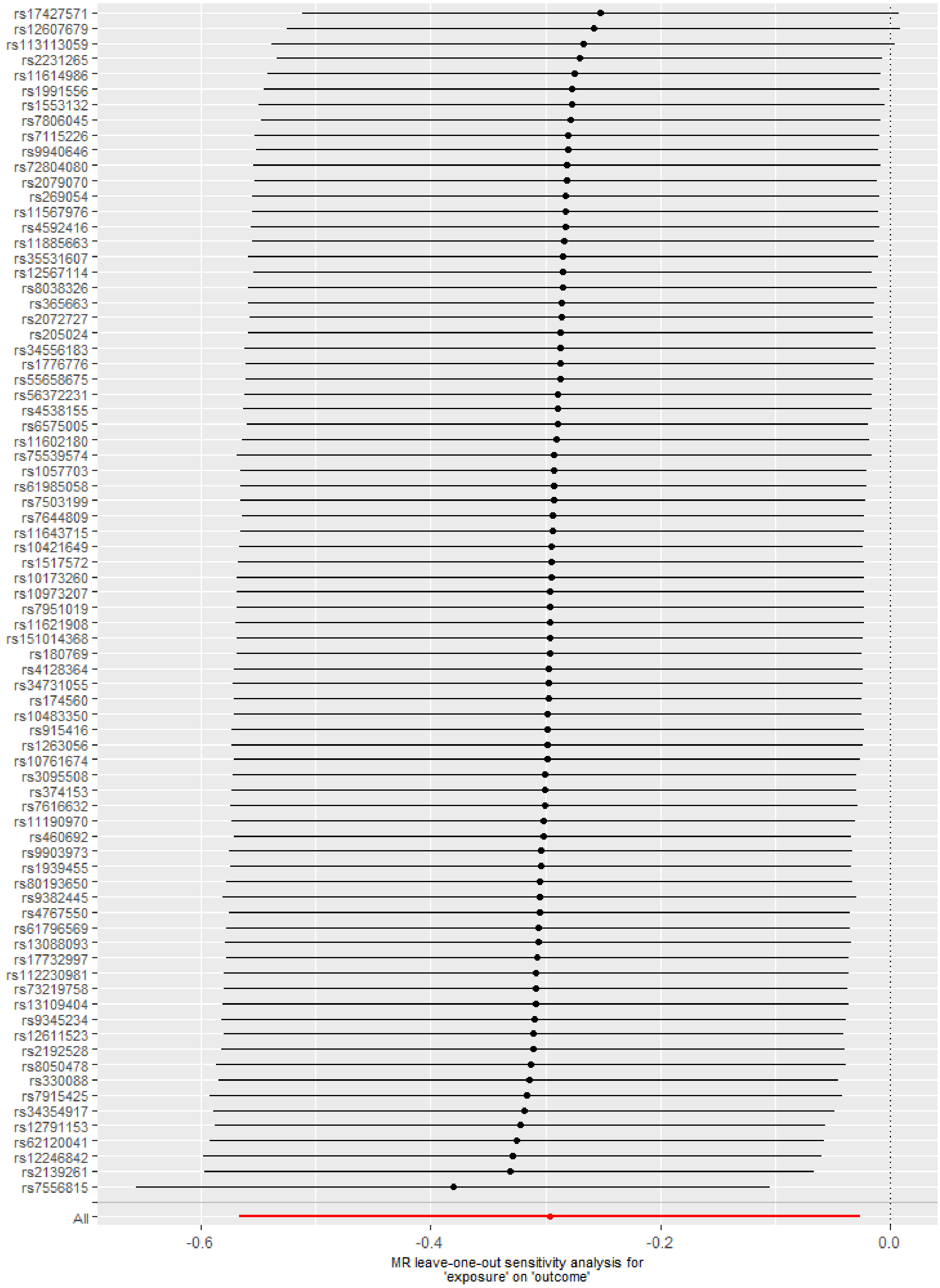

(H) High offspring birthweight

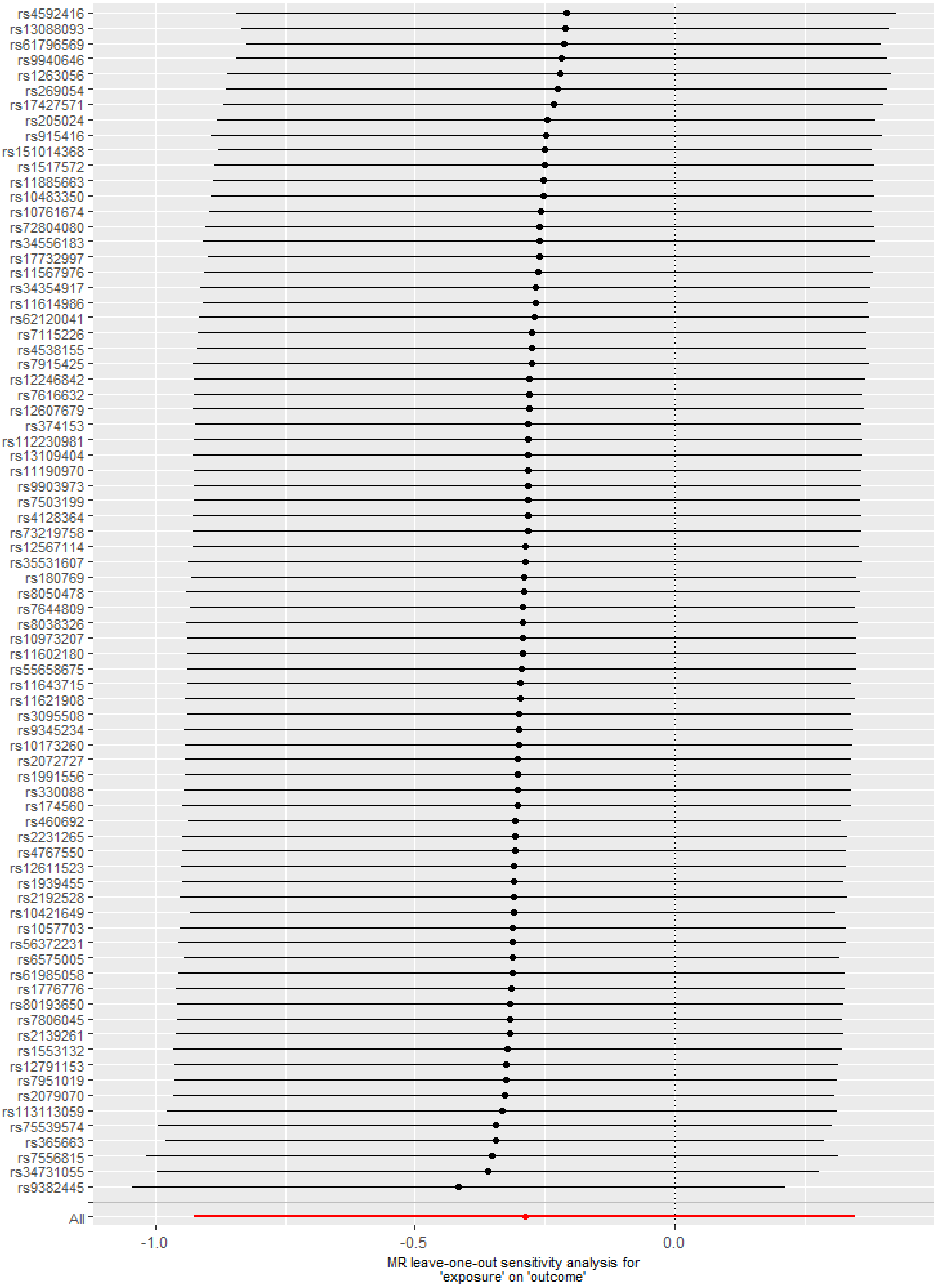

(I) Offspring birthweight (grams)

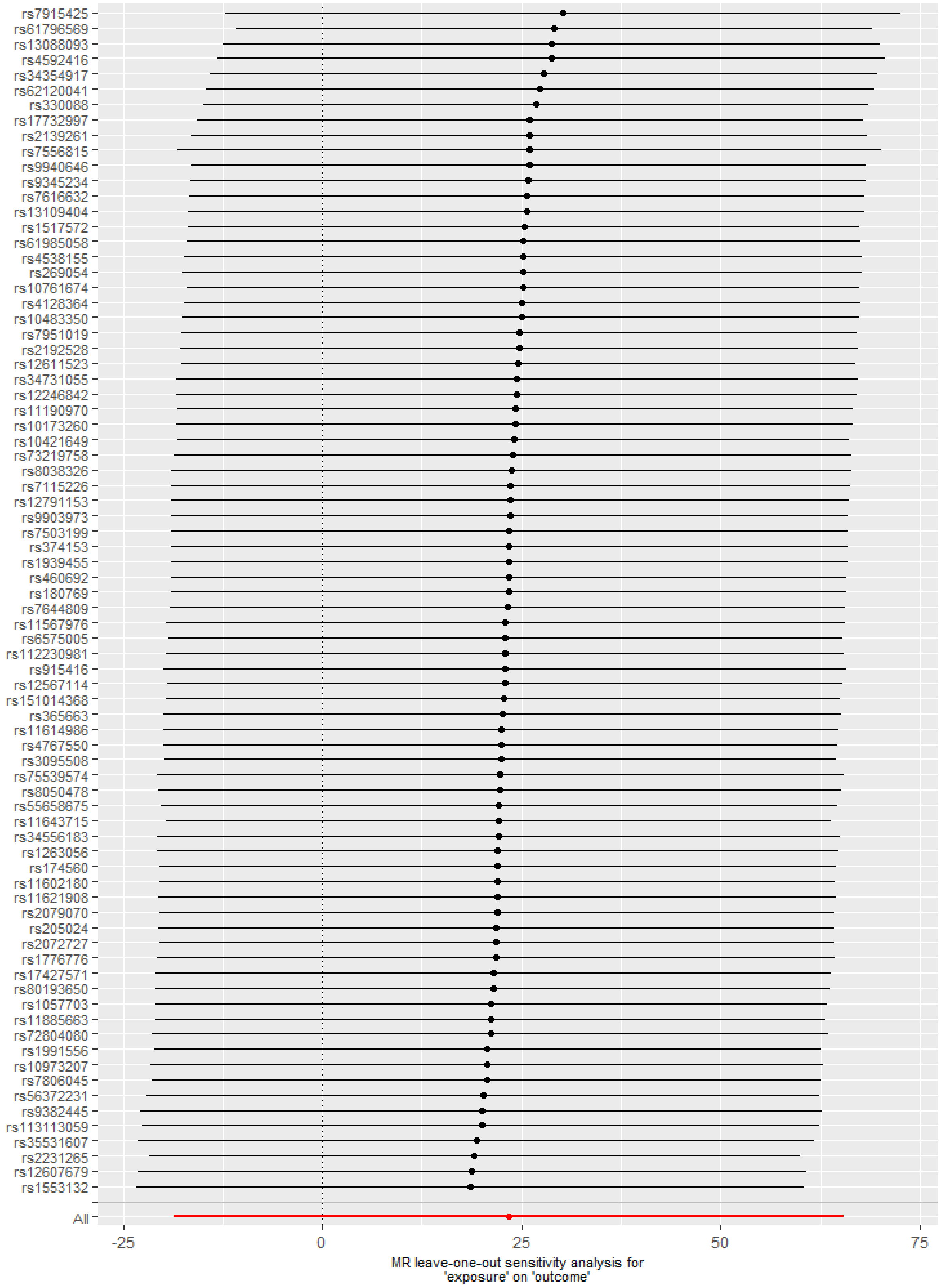

**Figure S7. Leave-one-out analyses for sleep duration on pregnancy and perinatal outcomes in other cohorts**

(A) Stillbirth

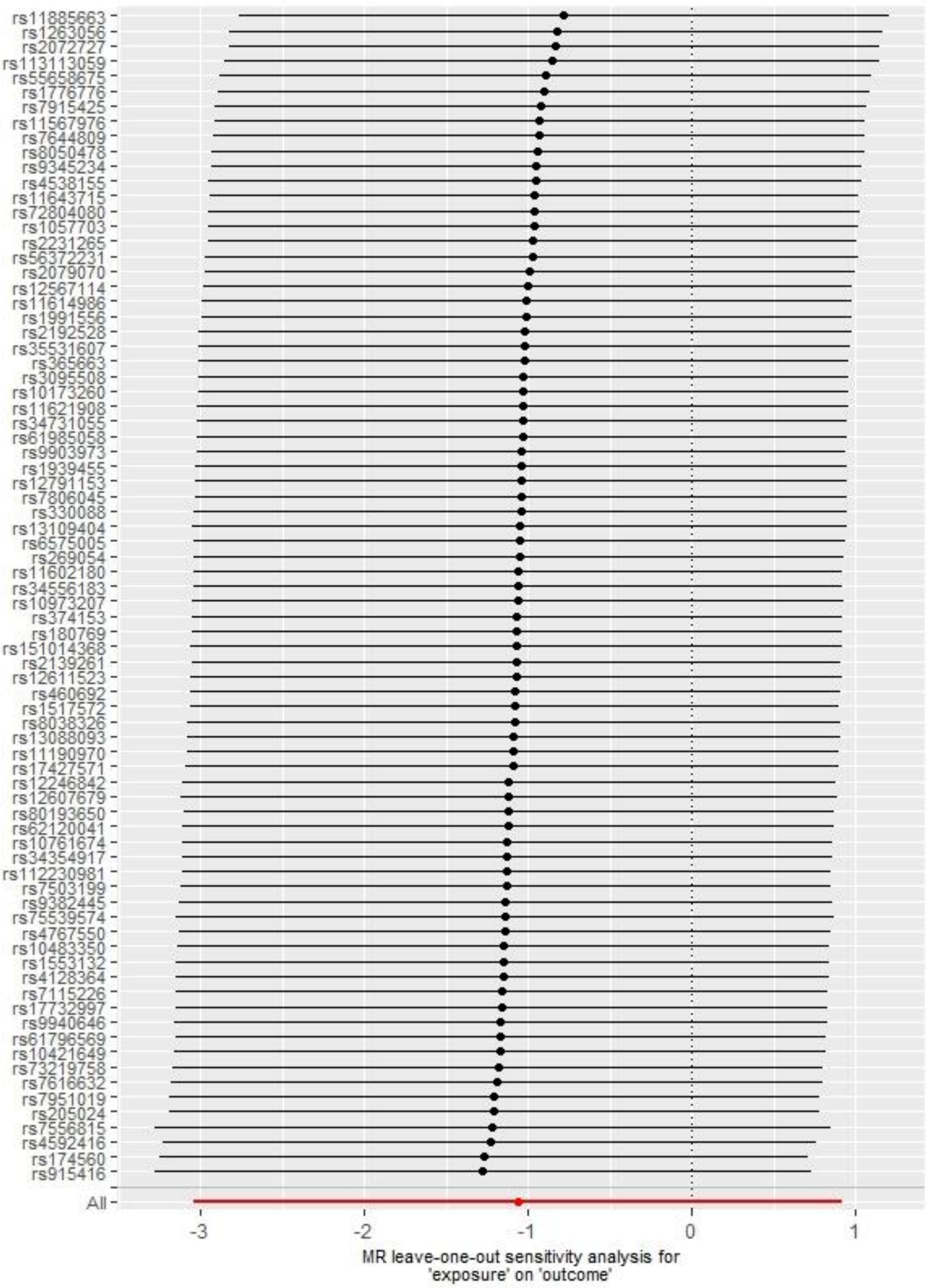

(B) Miscarriage

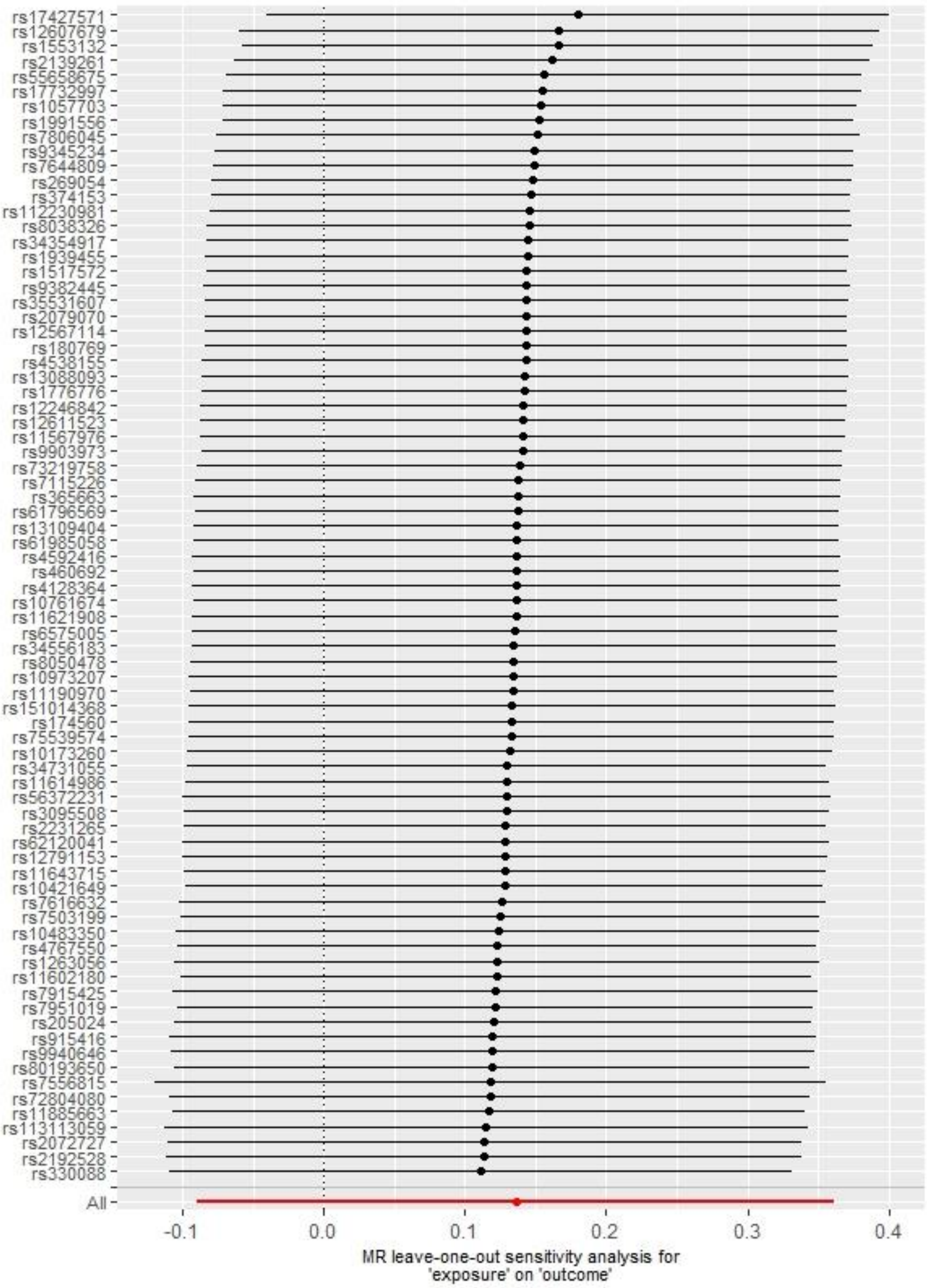

(C) Gestational diabetes

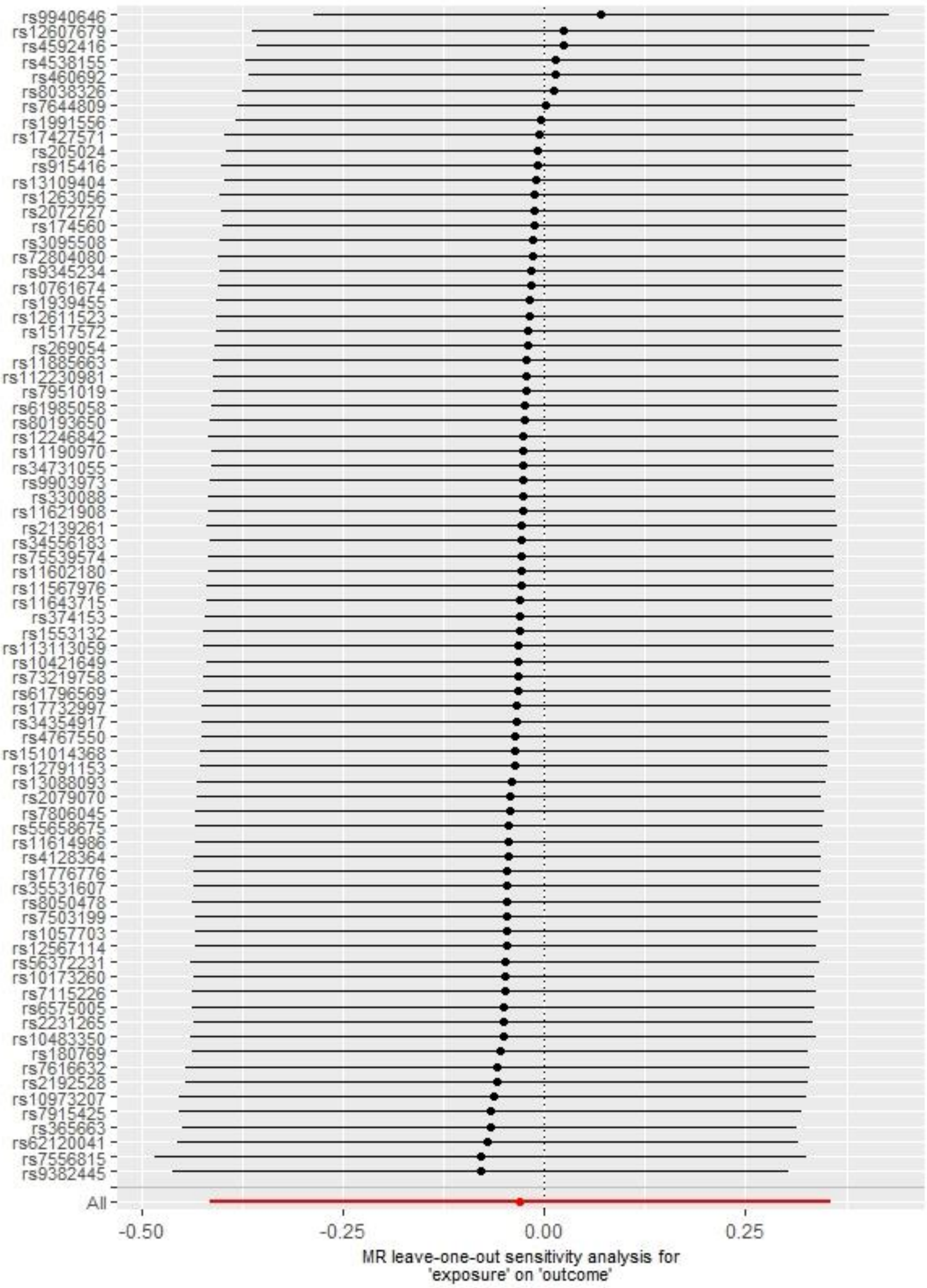

(D) Hypertensive disorders of pregnancy

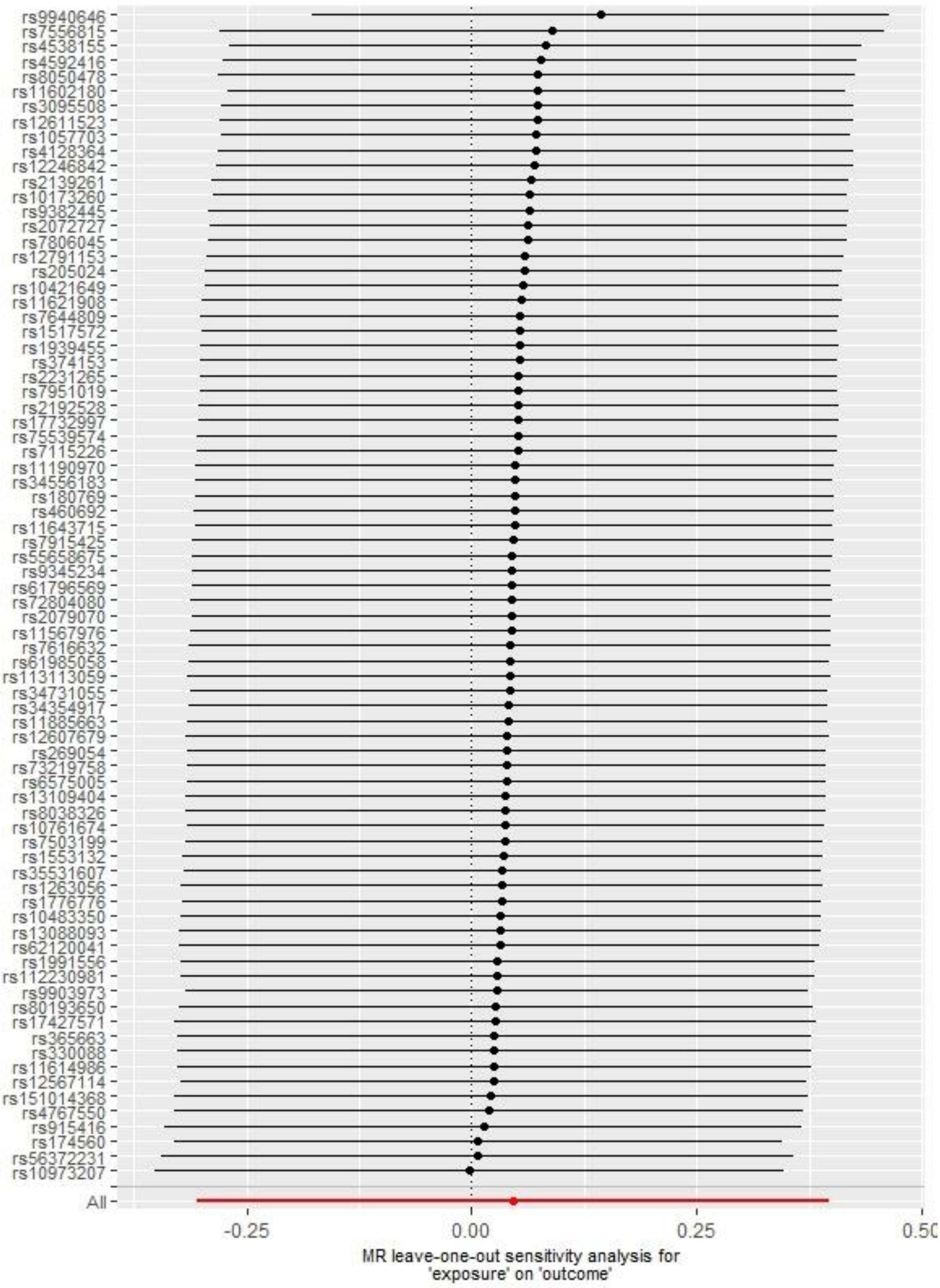

(E) Perinatal depression

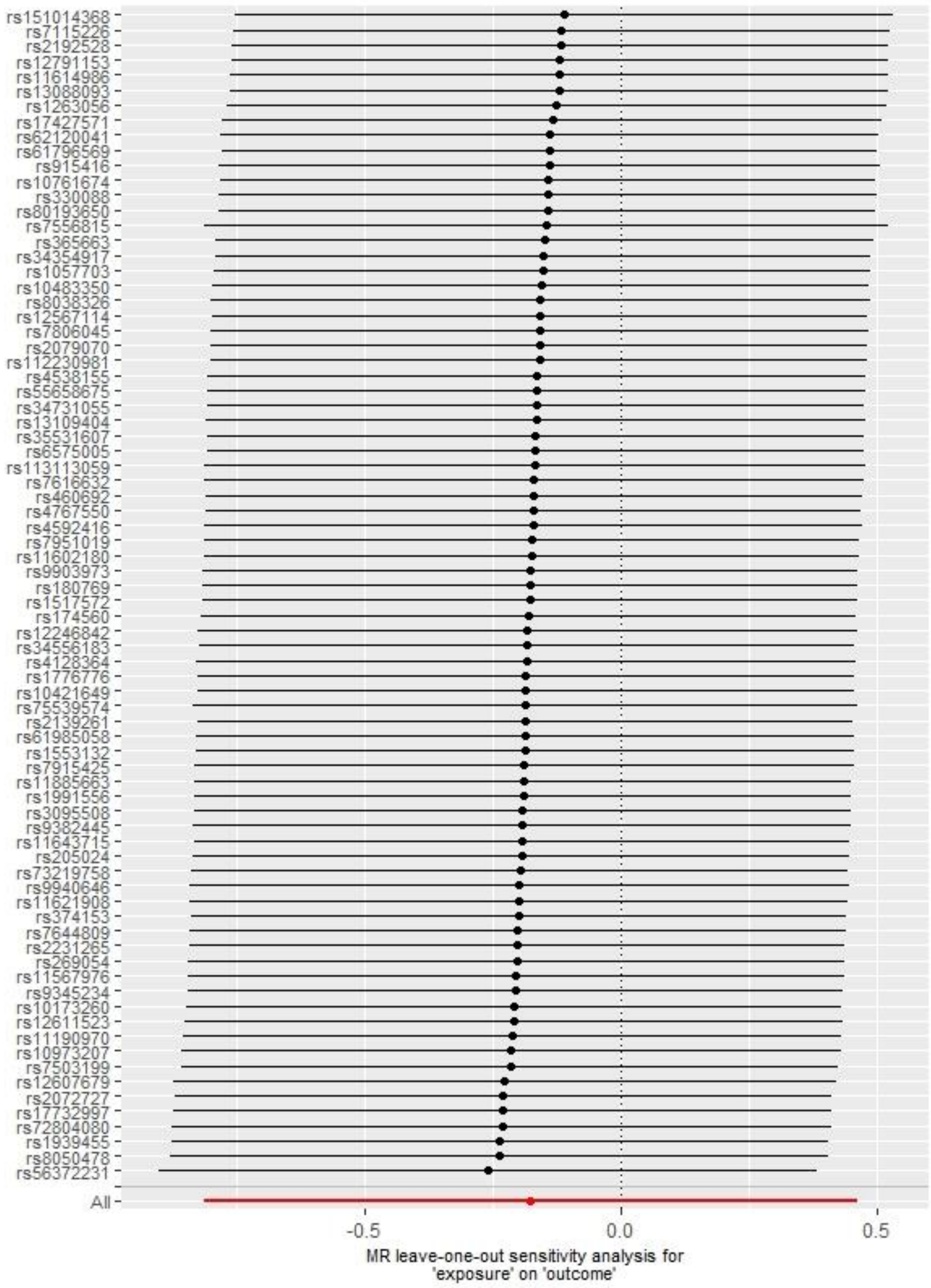

(F) Preterm birth

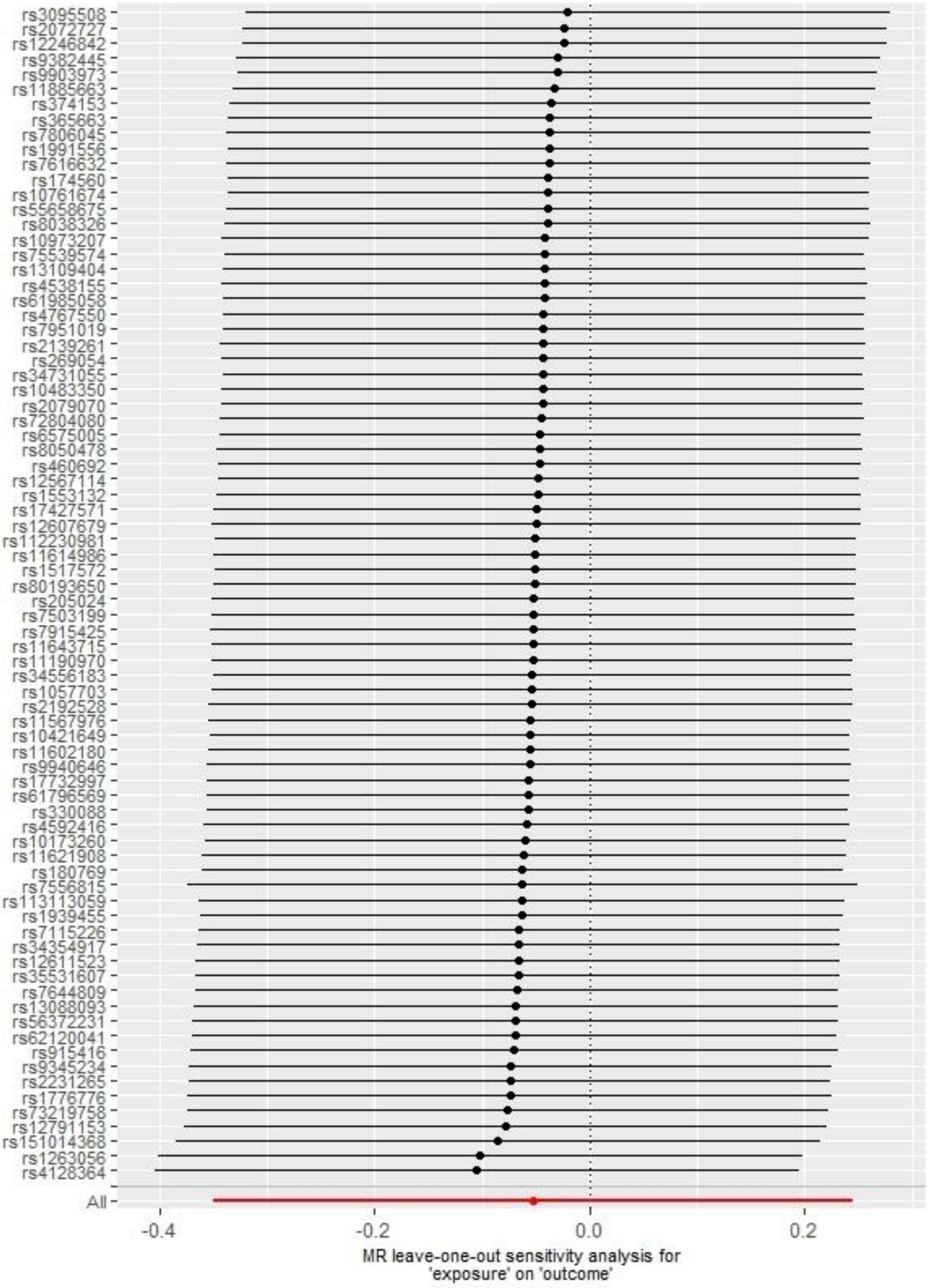

(G) Low offspring birthweight

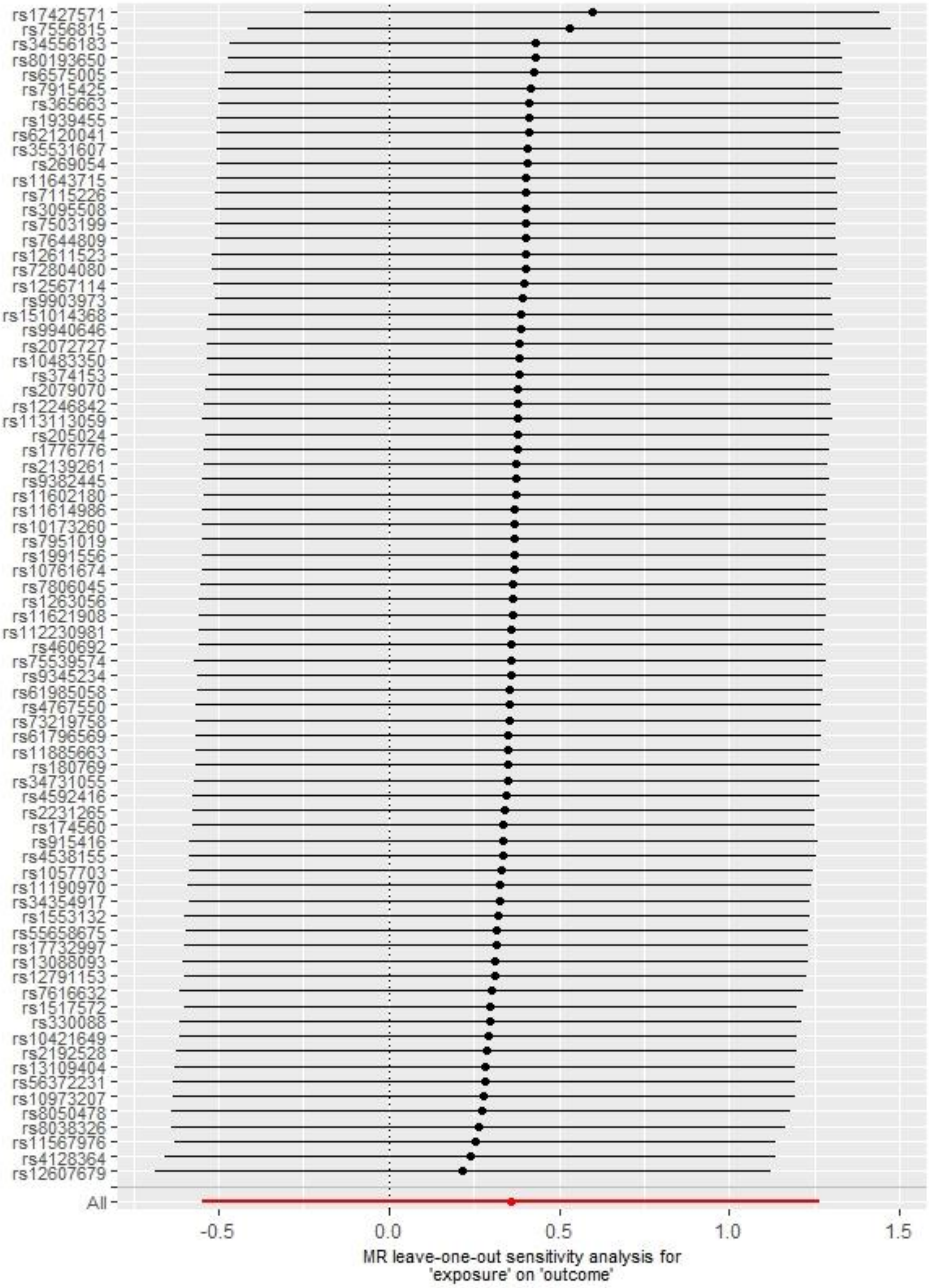

(H) High offspring birthweight

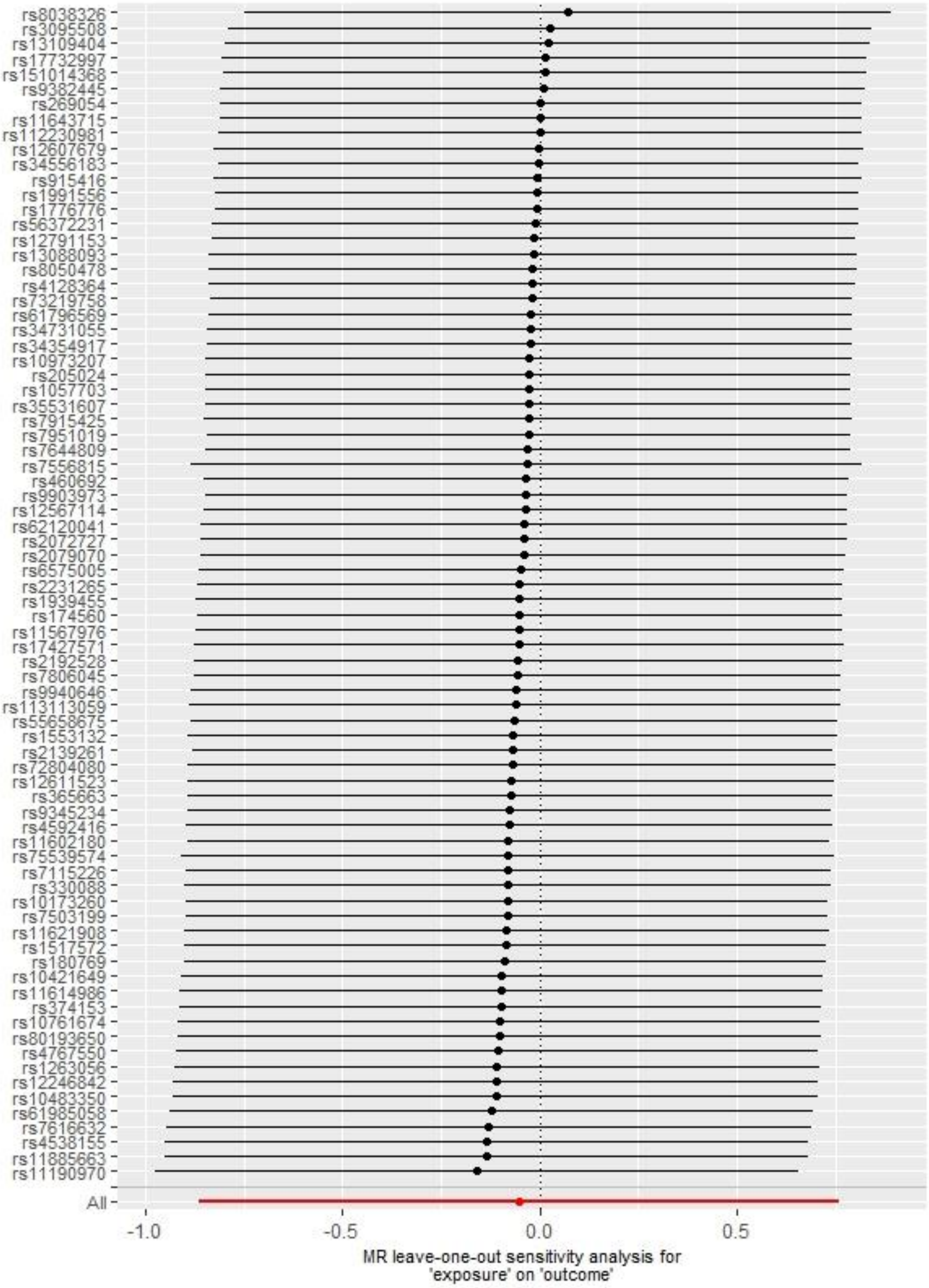

(I) Offspring birthweight (grams)

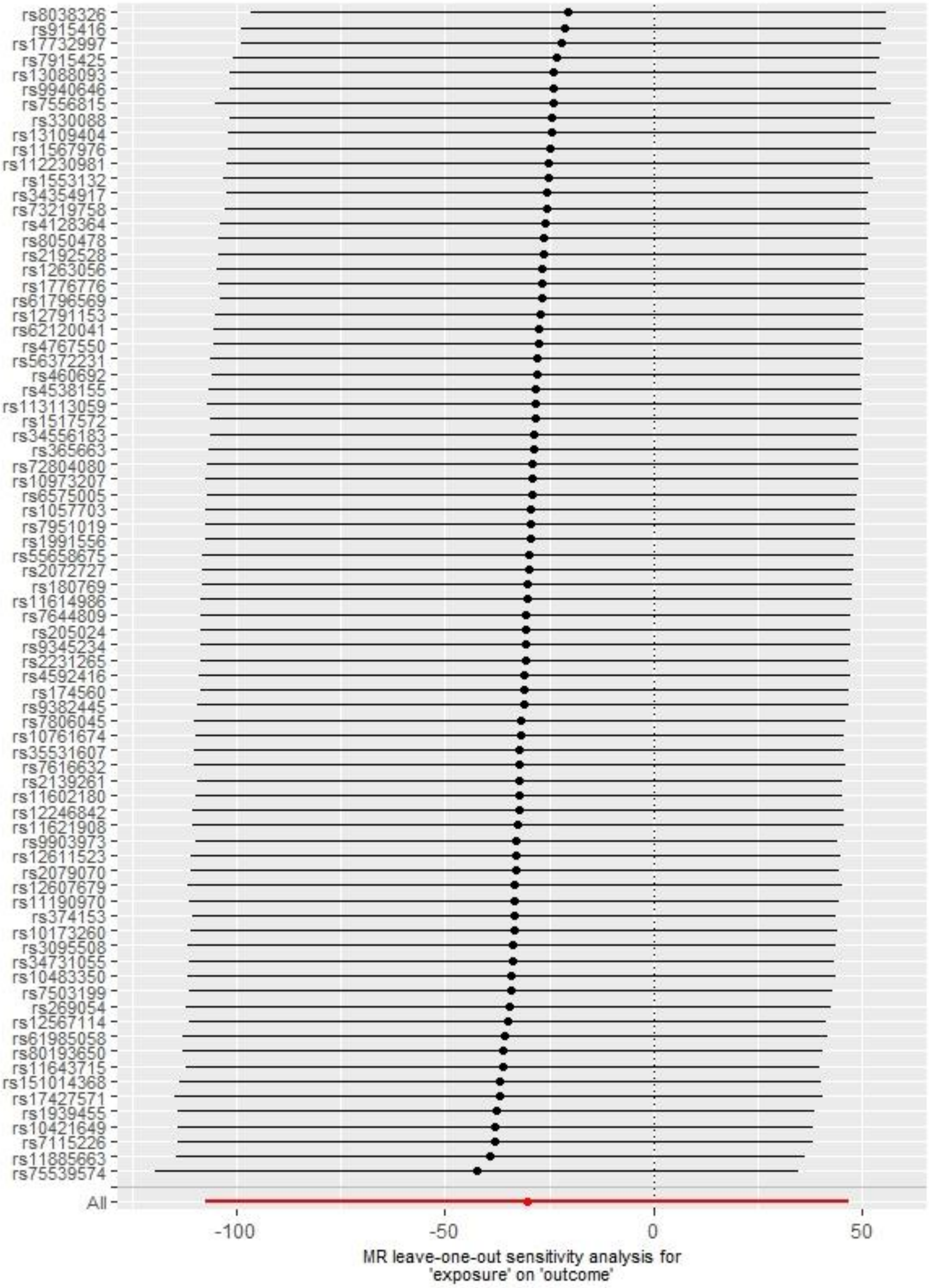

**Figure S8. Two-sample Mendelian randomization associations of sleep duration with pregnancy and perinatal outcomes using inverse variance weighted**

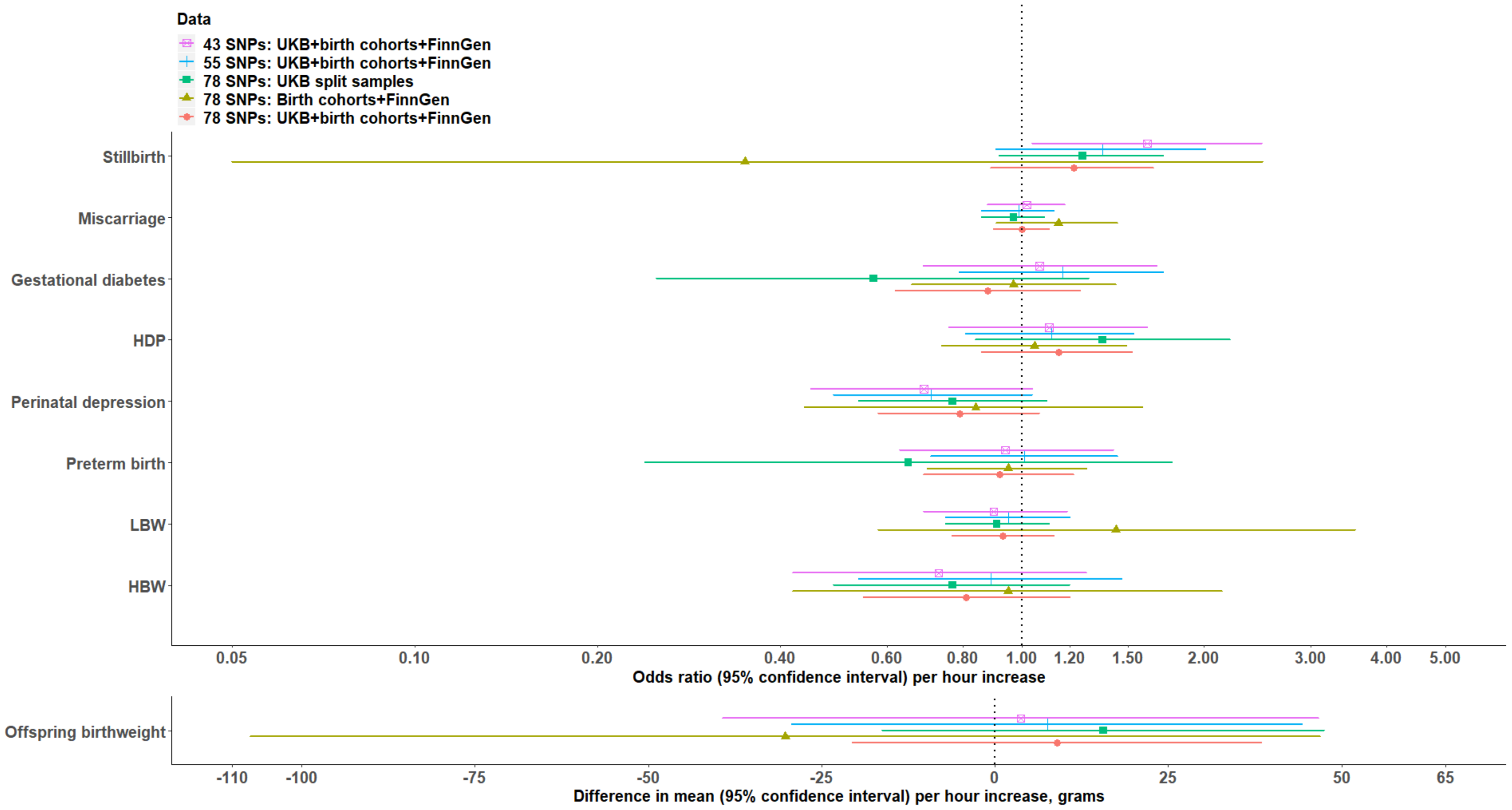


The genome-wide association study of sleep duration identified 78 genome-wide significant SNPs in its discovery cohort [21]. Among the 78 SNPs, 55 SNPS gave estimates in the same directions in the replication cohort. Among 55 SNPs, 43 SNPs achieved genome-wide significance in the meta-analysis of discovery and replication cohorts. FinnGen data were available for miscarriage, gestational diabetes, HDP, and preterm birth. Abbreviations: HBW, High offspring birthweight; HDP, hypertensive disorders of pregnancy; LBW, low offspring birthweight; SNP, single nucleotide polymorphisms; UKB, UK Biobank.

**Figure S9. Comparing multivariable regression associations of self-reported sleep duration categories with odds of LBW and HBW using different outcome comparison groups**

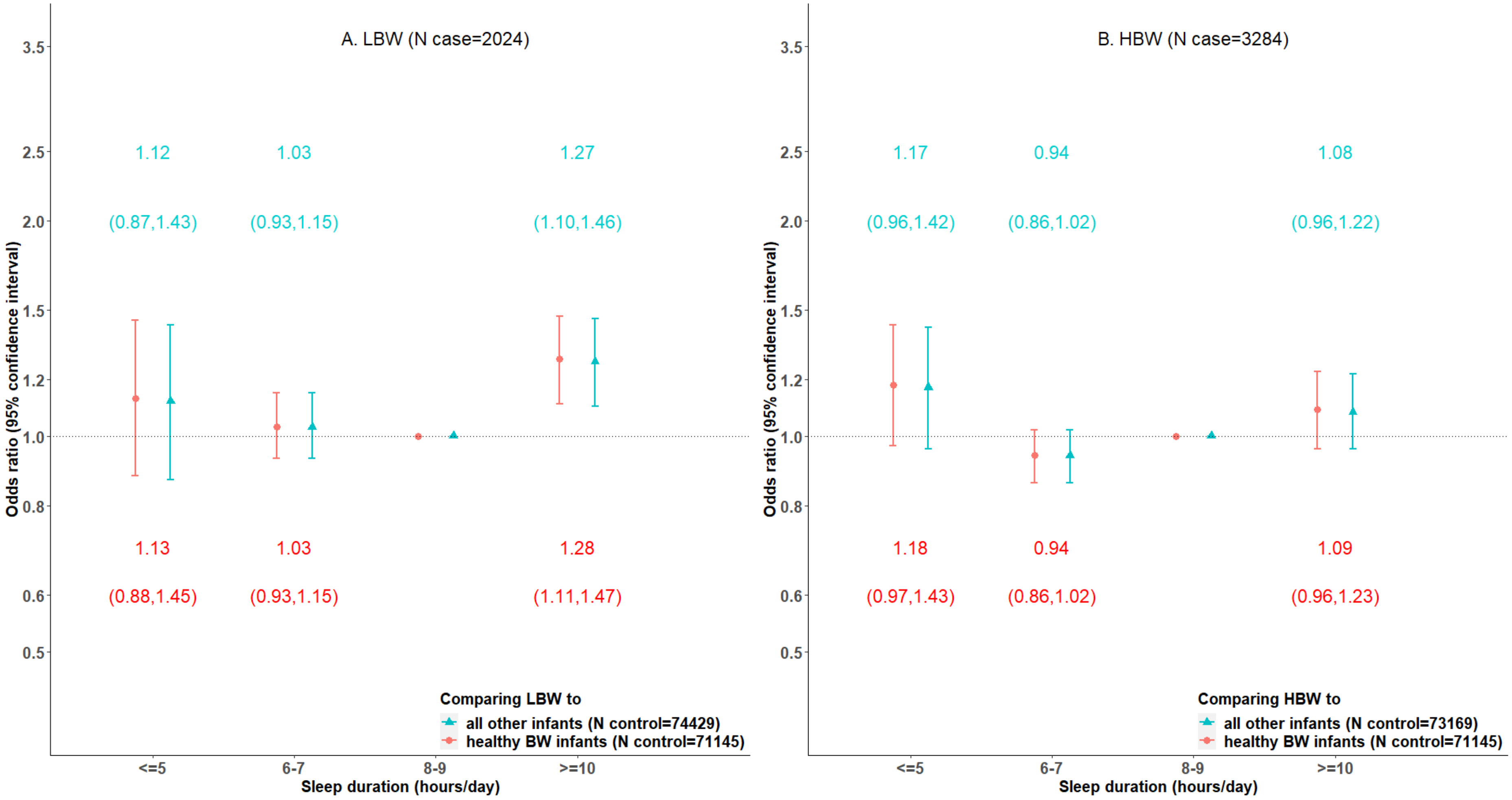


Estimates are from multiple imputation.

Abbreviations: BW, offspring birthweight; HBW, high offspring birthweight; LBW, low offspring birthweight.

**References**


1. Collins R. What makes UK Biobank special? Lancet. 2012; 379(9822):1173-4.
2. UK Biobank Mental health web-based questionnaire Version 1.3. 2017. Available from: https://biobank.ctsu.ox.ac.uk/crystal/crystal/docs/mental_health_online.pdf.
3. Bycroft C, Freeman C, Petkova D, Band G, Elliott LT, Sharp K, et al. The UK Biobank resource with deep phenotyping and genomic data. Nature. 2018; 562(7726):203-9.
4. Mitchell R, Hemani G, Dudding T, Corbin L, Harrison S, Paternoster L. UK Biobank Genetic Data: MRC-IEU Quality Control, version 2. 2019. Available from: https://data.bris.ac.uk/data/dataset/1ovaau5sxunp2cv8rcy88688v.
5. Henry A, Katsoulis M, Masi S, Fatemifar G, Denaxas S, Acosta D, et al. The relationship between sleep duration, cognition and dementia: a Mendelian randomization study. Int J Epidemiol. 2019; 48(3):849-60.
6. Burgess S, Davies NM, Thompson SG. Bias due to participant overlap in two-sample Mendelian randomization. Genet Epidemiol. 2016; 40(7):597-608.
7. Fraser A, Macdonald-Wallis C, Tilling K, Boyd A, Golding J, Davey Smith G, et al. Cohort Profile: the Avon Longitudinal Study of Parents and Children: ALSPAC mothers cohort. Int J Epidemiol. 2013; 42(1):97-110.
8. Boyd A, Golding J, Macleod J, Lawlor DA, Fraser A, Henderson J, et al. Cohort Profile: the 'children of the 90s'--the index offspring of the Avon Longitudinal Study of Parents and Children. Int J Epidemiol. 2013; 42(1):111-27.
9. Richmond RC, Timpson NJ, Felix JF, Palmer T, Gaillard R, McMahon G, et al. Using Genetic Variation to Explore the Causal Effect of Maternal Pregnancy Adiposity on Future Offspring Adiposity: A Mendelian Randomisation Study. PLoS Med. 2017; 14(1):e1002221.
10. Blair PS, Drewett RF, Emmett PM, Ness A, Emond AM. Family, socioeconomic and prenatal factors associated with failure to thrive in the Avon Longitudinal Study of Parents and Children (ALSPAC). Int J Epidemiol. 2004; 33(4):839-47.
11. Wright J, Small N, Raynor P, Tuffnell D, Bhopal R, Cameron N, et al. Cohort Profile: the Born in Bradford multi-ethnic family cohort study. Int J Epidemiol. 2013; 42(4):978-91.
12. Brand JS, Gaillard R, West J, McEachan RRC, Wright J, Voerman E, et al. Associations of maternal quitting, reducing, and continuing smoking during pregnancy with longitudinal fetal growth: Findings from Mendelian randomization and parental negative control studies. PLoS Med. 2019; 16(11):e1002972.
13. Magnus P, Birke C, Vejrup K, Haugan A, Alsaker E, Daltveit AK, et al. Cohort Profile Update: The Norwegian Mother and Child Cohort Study (MoBa). Int J Epidemiol. 2016; 45(2):382-8.
14. Paltiel L, Anita H, Skjerden T, Harbak K, Bækken S, Kristin SN, et al. The biobank of the Norwegian Mother and Child Cohort Study – present status. Nor J Epidemiol. 2014; 24(1-2):29-35.
15. Magnus MC, Miliku K, Bauer A, Engel SM, Felix JF, Jaddoe VWV, et al. Vitamin D and risk of pregnancy related hypertensive disorders: mendelian randomisation study. BMJ. 2018; 361:k2167.
16. MoBaGenetics1.0. 2020. Available from: https://github.com/folkehelseinstituttet/mobagen/wiki/MoBaGenetics1.0.
17. Projects that have contributed to MoBa Genetics. 2020. Available from: https://github.com/folkehelseinstituttet/mobagen/wiki/Projects-that-have-contributed-to-MoBa-Genetics.
18. FinnGen. FinnGen Documentation of R5 release. 2021. Available from: https://finngen.gitbook.io/documentation/.
19. Kiiskinen T, Mars NJ, Palviainen T, Koskela J, Rämö JT, Ripatti P, et al. Genomic prediction of alcohol-related morbidity and mortality. Transl Psychiatry. 2020; 10(1):23.
20. Zhou W, Zhao Z, Nielsen JB, Fritsche LG, LeFaive J, Gagliano Taliun SA, et al. Scalable generalized linear mixed model for region-based association tests in large biobanks and cohorts. Nat Genet. 2020; 52(6):634-9.

21. Dashti HS, Jones SE, Wood AR, Lane JM, van Hees VT, Wang H, et al. Genome-wide association study identifies genetic loci for self-reported habitual sleep duration supported by accelerometer-derived estimates. Nat Commun. 2019; 10(1):1100.
22. Anderson EL, Richmond RC, Jones SE, Hemani G, Wade KH, Dashti HS, et al. Is disrupted sleep a risk factor for Alzheimer's disease? Evidence from a two-sample Mendelian randomization analysis. Int J Epidemiol. 2020; 50(3):817-28.
23. Burgess S, Small DS, Thompson SG. A review of instrumental variable estimators for Mendelian randomization. Stat Methods Med Res. 2017; 26(5):2333-55.
24. Sun YQ, Burgess S, Staley JR, Wood AM, Bell S, Kaptoge SK, et al. Body mass index and all cause mortality in HUNT and UK Biobank studies: linear and non-linear mendelian randomisation analyses. BMJ. 2019; 364:l1042.
25. Palmer TM, Sterne JA, Harbord RM, Lawlor DA, Sheehan NA, Meng S, et al. Instrumental variable estimation of causal risk ratios and causal odds ratios in Mendelian randomization analyses. Am J Epidemiol. 2011; 173(12):1392-403.
26. Hirshkowitz M, Whiton K, Albert SM, Alessi C, Bruni O, DonCarlos L, et al. National Sleep Foundation's updated sleep duration recommendations: final report. Sleep Health. 2015; 1(4):233-43.
27. Lu Q, Zhang X, Wang Y, Li J, Xu Y, Song X, et al. Sleep disturbances during pregnancy and adverse maternal and fetal outcomes: a systematic review and meta-analysis. Sleep Med Rev. 2020; 58:101436.
